# Autoantibodies From Connective Tissue Diseases Penetrate Cells and Exert Functional Properties

**DOI:** 10.64898/2026.06.29.26356321

**Authors:** Aurélien Chepy, Celine Thiesen, Marie-Elise Martel, Maithri Aspari, Solange Vivier, Jakob Hauge Mikkelsen, Emilia Holm, Sören Magnus Morische, Esben Næser, Julien Sottiaux, Marie Duhamel, Klaus Søndergaard, Sarah Barslund Petersen, Marie Mistretta, Maxime Secq, Malene Hvid, Josephine Geertsen Keller, David Abraham, Elif Hatipoglu, Clément Chauvet, Lucile Guilbert, Emilie Grarup Jensen, Søren R. Paludan, Christian Brix Folsted Andersen, Antonino Bongiovanni, Eric Hachulla, Michel Salzet, Joanna Maria Kalucka, Birgitta R. Knudsen, Sylvain Dubucquoi, Stinne Greisen, David Launay, Cinzia Tesauro, Meryem Tardivel, Bent Deleuran, Vincent Sobanski

**Affiliations:** Univ. Lille, Inserm, CHU Lille, U1286 – INFINITE – Institute for Translational Research in Inflammation; F-59000 Lille, France; CHU Lille, Département de Médecine interne et Immunologie Clinique, Centre de Référence des Maladies Auto-immunes Systémiques Rares du Nord et Nord-Ouest, Méditerranée et Guadeloupe (CeRAINOM); F-59000 Lille, France; Department of Biomedicine, Aarhus University; Aarhus, 8000, Denmark; Department of Rheumatology, Aarhus University Hospital; Aarhus 8200, Denmark; Department of Clinical Medicine, Aarhus University; Aarhus 8000, Denmark; Université Lille, Inserm, CHU Lille, U1192 - Protéomique Réponse Inflammatoire Spectrométrie de Masse – PRISM; F-59000 Lille; Department of Molecular Biology and Genetics, Aarhus University; Aarhus 8000, Denmark; UCL Centre for Rheumatology, Department of Inflammation and Rare Diseases, Division of Medicine, University College London; London, United Kingdom; STADIUS Center for Dynamical Systems, Signal Processing and Data Analytics, Department of Electrical Engineering (ESAT), KU Leuven, Leuven, Belgium & Woman and Child, Department of Development and Regeneration, KU Leuven; Leuven, Belgium; CHU Lille, Institut d’Immunologie; Lille, France; Center for Immunology of Viral Infections, Aarhus University; Aarhus, Denmark; Univ. Lille, CNRS, Inserm, CHU Lille, Institut Pasteur de Lille, US 41 - UAR 2014 – PLBS; Lille, 59000, France; Institut Universitaire de France (IUF); Paris, France

## Abstract

Antinuclear antibodies (ANAs) are a hallmark of connective tissue diseases (CTDs) and serve as robust diagnostic biomarkers. Because their cognate antigens are intracellular, ANAs have long been considered non-pathogenic in CTDs. Here, using systemic sclerosis (SSc) associated anti-topoisomerase I antibodies (ATAs) as a model, we provide data challenging this view. We show that ANAs enter living cells, accumulate in nuclei, and engage their intracellular antigen. Nuclear ATAs inhibit topoisomerase I enzymatic activity, induces DNA damage, fibrosis, and, through the STING pathway, activates type I interferon production. We further identify neonatal Fc receptor (FcRn)-dependent intracellular trafficking as a key determinant of ANAs nuclear access and demonstrate that pharmacological FcRn blockade impairs ATAs functionality. These findings reveal a previously unrecognized intracellular effector function of ANAs and establish a mechanistic framework by which ANAs may directly contribute to tissue injury in CTDs.

## Main Text

Connective tissue diseases (CTDs) are characterized by the presence of antinuclear antibodies (ANAs), which serve as robust diagnostic and prognostic biomarkers across disorders such as systemic sclerosis (SSc), systemic lupus erythematosus (SLE) and inflammatory myopathies (*1*). Despite their key clinical relevance, the contribution of ANAs to pathogenesis has remained unresolved. This uncertainty stems, in part, from the intracellular and nuclear localization of their cognate antigens, leading to the longstanding view that ANAs represent non-pathogenic epiphenomena rather than central mediators of disease (*2*, *3*).

SSc is a particularly informative model to revisit this assumption. SSc is a severe CTD characterized by a self-reinforcing interplay among autoantibodies, vasculopathy, and fibrosis, with endothelial cells and fibroblasts playing a central role in tissue damage (*4*). At a molecular level, sustained activation of innate immune pathways, including a type I interferon (IFN) signature, is a hallmark of SSc and other CTDs (*5*, *6*). ANAs are detected in the vast majority of patients with SSc. Notably, the ANA subset anti–topoisomerase I antibodies (ATAs) is associated with more severe disease (*7*, *8*). Recent data has revived interest in the ability of ANAs to enter living cells (*9*, *10*) and exert pathophysiological action. Electroporation of immunoglobulins G (IgG) into the nuclei of myocytes indeed induces molecular profiles similar to those observed in muscle biopsies from patients with myositis of the same serotype (*11*). Nevertheless, the direct pathogenicity of ANAs and the underlying mechanisms are yet to be demonstrated (*10–13*). Here, using SSc and ATAs as a model system, we test the hypothesis that ANAs can penetrate cells, engage intracellular targets, and elicit biologically meaningful responses. By integrating high-resolution imaging, functional assays, and *ex vivo* human tissue explants, we show that ANAs enter living cells, accumulate in nuclei, and engage their intracellular antigen. Nuclear ATAs inhibit topoisomerase I (TOP1) enzymatic activity, induces DNA damage, and, through the Stimulator of Interferon Genes (STING) pathway, activates type I IFN production. Furthermore, we identify neonatal Fc receptor (FcRn)-dependent intracellular trafficking as a key determinant of ANAs cell and nuclear access.

## Results

In order to functionally characterize ANAs, we purified IgG from patients with SSc (SSc-IgG), patients with SLE (SLE-IgG), and healthy controls (HC-IgG). The workflow is shown in **Supplementary Fig. 1**. We evaluated IgG fractions from SSc patients who had autoantibodies to TOP1 (ATA^+^-IgG), centromere (ACA^+^-IgG), RNA polymerase III (ARA^+^-IgG), PM/Scl (anti-PM/Scl^+^-IgG), and SLE patients who had antibodies to DNA (anti-DNA^+^-IgG) and Sm (anti-Sm^+^-IgG). We also used purified ATAs and the corresponding ATA-depleted IgG fractions (ATA-depleted IgG). We prepared fluorescent ATAs (fluo-ATAs) and fluorescent HC-IgG (fluo-HC-IgG).

### Antinuclear Antibodies Access Nuclei and Engage with their Intracellular Antigen

To evaluate whether ANAs can penetrate cells and thus validate our hypothesis, we first examined the presence of IgG in SSc tissues. Confocal microscopy images of skin biopsies from SSc patients revealed clear nuclear IgG staining, whereas no comparable signal was observed in HC skin (**Fig. 1A, Fig. S2, Table S1**). To test whether IgG can enter living cells, fibroblasts and endothelial cells were incubated with SSc-IgG, SLE-IgG and HC-IgG (**outlined in Fig. S3**). Strikingly, IgG nuclear localization differed markedly: pronounced nuclear localization occurred only with SSc-IgG and SLE-IgG, whereas HC-IgG showed minimal to no nuclear signal, with only rare intracytoplasmic puncta (**Fig. 1B**, **Fig. S4A-E**). Both autologous (i.e. fibroblasts and IgG from the same patient) and heterologous (i.e. fibroblasts and IgG from different patients) SSc-IgG entered SSc-FB and reached the nucleus (**Fig. S4F**). We also observed similar results with ATAs (**Fig. 1B, Fig. S4C-D**). Live-cell imaging confirmed immediate internalization of fluo-ATAs by fibroblasts, with rapid nuclear localization, as demonstrated by 3D high-resolution live microscopy (**Movies S1-S5)**. Similar patterns of nuclear localization were seen in skin explants (NativeSkin®) used as an *ex vivo* model and exposed to ATAs, compared to those treated with HC-IgG (**Fig. 1C**). These findings support that IgG can enter living cells in both cellular and skin explant models. We observed ANAs to efficiently reach the nucleus; we then investigated the intranuclear ANAs and corresponding nuclear antigens colocalization (**Fig. S5A-C**). Three-dimensional analyses further confirmed intranuclear IgG localization (**Fig. 1D, Fig. S4G, Movie S6**). To confirm that the nuclear entry of ATAs is antigen-specific, we mixed ATAs with TOP1 prior to cell incubation. We observed a marked reduction in nuclear spots in the ATA-TOP1 complex condition compared with ATAs alone (**Fig. 1D**). Importantly, internalized ANAs interacted with their respective targets in cells and explants exposed to ATAs, as proximity ligation assays (PLA) revealed contact occurring within 40 nm (**Fig. 1E, Fig. S5D**). High-resolution nanoscopy identified intimate contacts between SSc-IgG and nuclear antigens (**Fig. 1F, Fig. S5E-F**). Together, these results show that ANAs not only enter living cells and reach the nucleus but also physically engage their cognate nuclear antigens.

**Figure 1.**
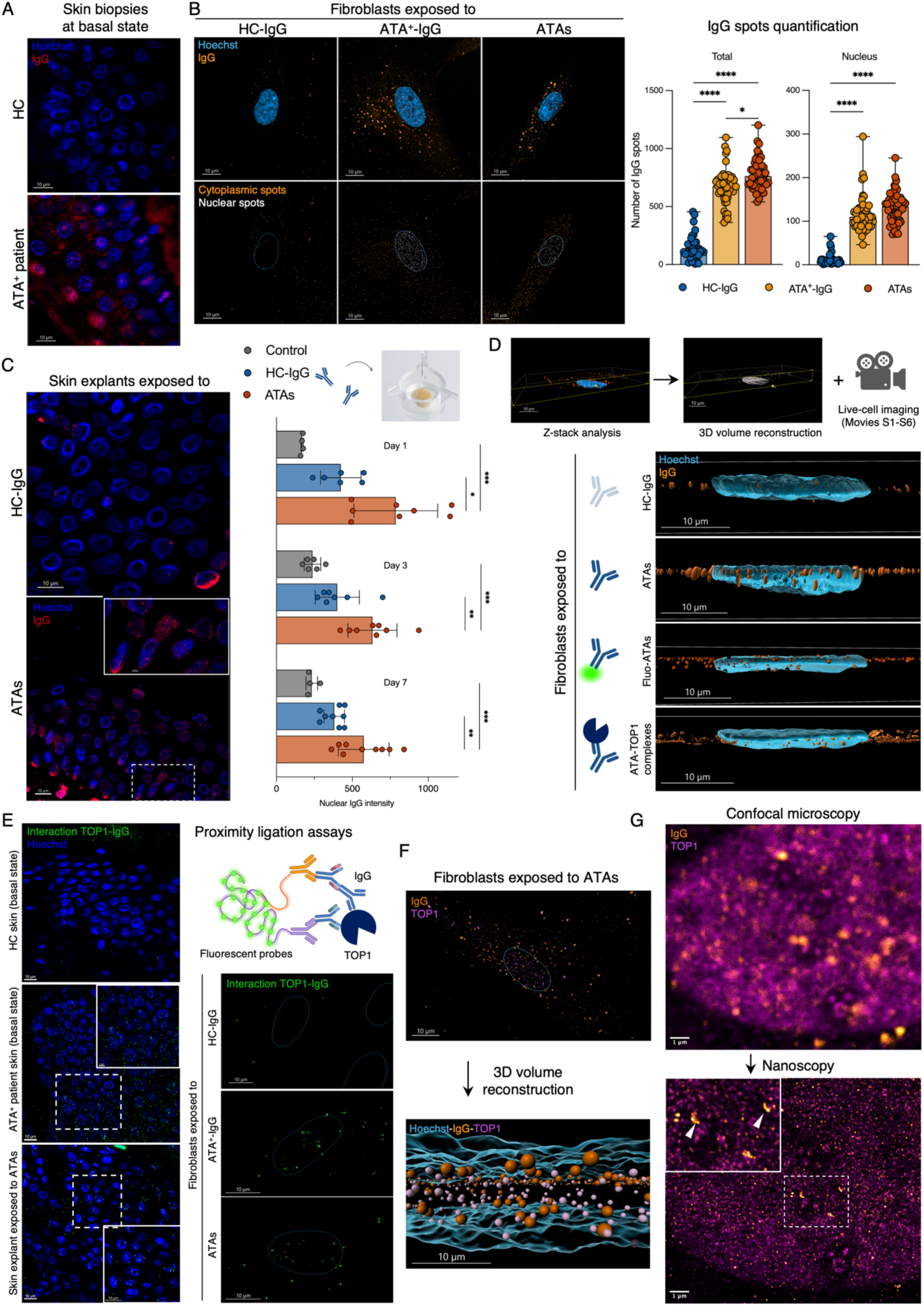
Antinuclear Antibodies Access Nuclei and Engage their Intracellular Antigen. **(A)** Confocal micrographs of IgG staining in HC and ATA^+^ SSc skin biopsies. **(B)** Micrographs (left) and quantification (right) of intracellular and nuclear IgG in fibroblasts after 1-hour incubation with indicated IgG preparations. Nuclear and cytoplasmic spots pseudo-colored orange and white following Hoechst-based segmentation (IMARIS). *n* = 10 subjects for HC-IgG, ATA^+^-IgG, *n* = 3 for ATAs; for each condition ≥ 300 analyzed cells **(C)** Micrographs (left) and quantification (right) of intranuclear IgG in NativeSkin® explants treated with medium alone (control, *n* = 3), HC-IgG (*n* = 3), or ATAs (*n* = 3) at days 1, 3, 7. *n* = 2 coverslips analyzed per explant. **(D)** Z-stack and 3D reconstruction (IMARIS) of fibroblast nuclei after 1-hour exposure to HC-IgG, ATAs, fluo-ATAs, or TOP1-ATA complexes. **(E)** TOP1-IgG PLA in SSc and HC skin biopsies, NativeSkin® explants (left), and fibroblasts (right); individual fluorescent spots indicate TOP1-IgG interactions occurring within 40 nm. **(F)** IgG/TOP1 co-staining and 3D reconstruction (IMARIS) in ATAs-exposed fibroblasts (1 hour). **(G)** Confocal (top) and STED (bottom) of ATAs-exposed fibroblasts. IgG concentrations: 100 µg/mL for total IgG and 25 µg/mL for ATAs (B, D, E, F); 450 µg/explant for HC-IgG, 300 µg/explant for ATAs (C, E). Box plots show median and full range. *P < 0.05, **P < 0.01, ***P < 0.001, ****P < 0.0001; Kruskal-Wallis with Dunn’s post-hoc test (B); one-way ANOVA with Tukey’s post-hoc test (C). Airyscan confocal (∼140 nm lateral resolution) and STED (∼41 nm lateral resolution) for illustration; spinning disk for *ex vivo* quantification (G).

### Internalized Anti-Topoisomerase Antibodies Inhibit Topoisomerase I and Induce DNA Damage

We assessed the functionality of ANA internalization in fibroblasts, focusing on ATAs, using compartment-specific proteomics in order to separate cytoplasmic and nuclear profiles after exposure to SSc-IgG or HC-IgG. Partial Least Squares Discriminant Analyses (PLS-DA) showed a clear segregation by ANAs serotypes, with ATA^+^-IgG inducing a homogeneous cytoplasmic profile distinct from ACA^+^-IgG and HC-IgG. ATA^+^-IgG *vs.* HC-IgG comparison identified 177 differentially expressed proteins, including increased type I collagen and α–smooth muscle actin, with an enrichment in type I IFN signaling and eukaryotic elongation factor pathways (**Fig. 2A, Fig. S6A, C-D**). Nuclear PLS-DA was also clustered by ANAs serotypes. In ATA^+^-IgG *vs.* HC, 1,282 proteins were differentially expressed, including reduced Poly [ADP-ribose] polymerase 1 (PARP-1) and high-mobility group box 1, with enrichment in DNA/RNA metabolism related pathways (**Fig. 2A, Fig. S6B, E-F**), which aligned with the profile produced by intranuclear antibody-antigen interactions.

**Figure 2.**
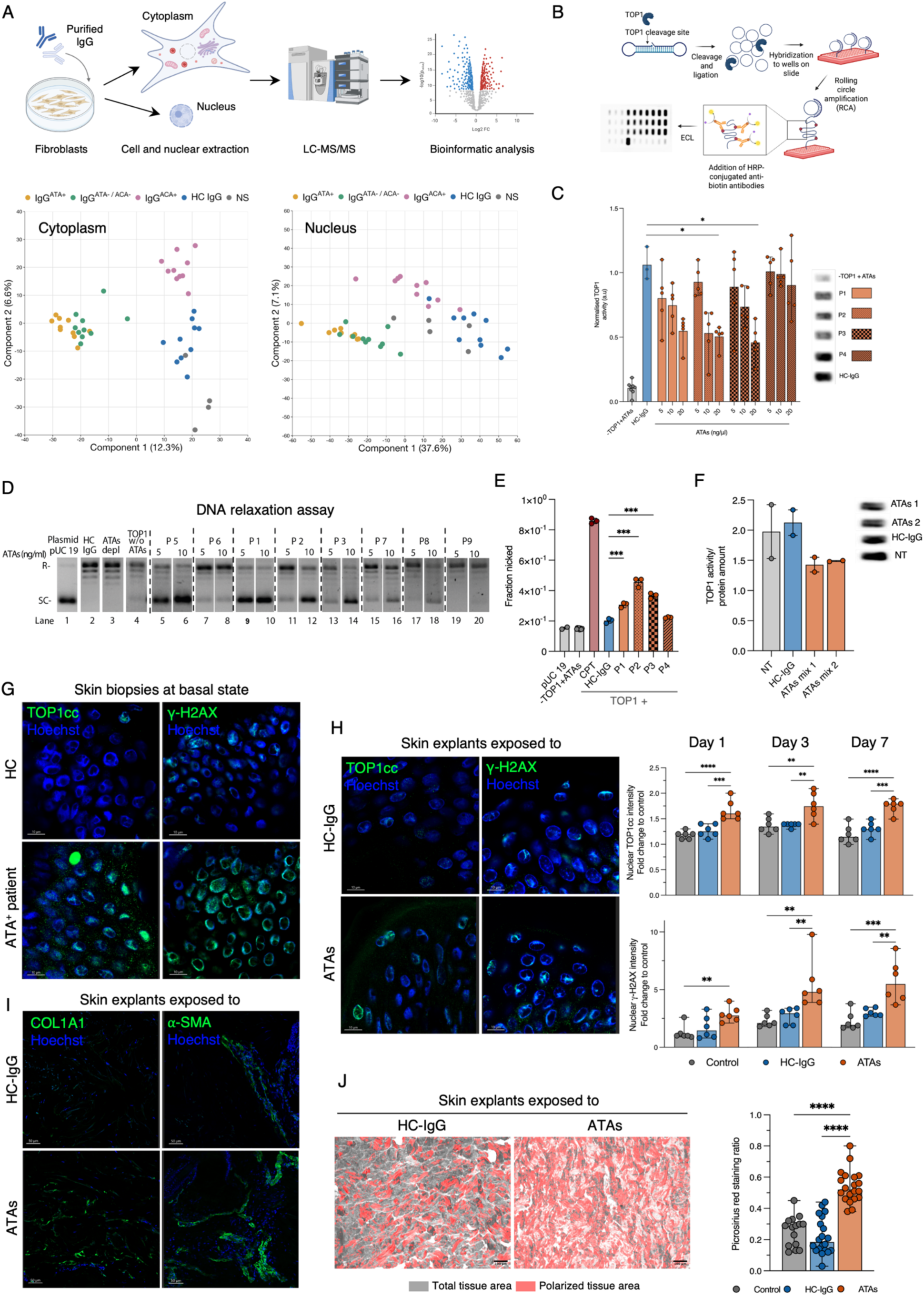
Internalized Antinuclear Antibodies Inhibit Topoisomerase I and Induce DNA Damage. **(A)** Compartmental proteomics. Fibroblasts cultured for 3 days with HC-IgG or SSc-IgG were fractionated for liquid chromatography-mass spectrometry. *n* = 10 per serotype (ATA⁺, ACA⁺, ATA⁻/ACA⁻); *n* = 9 for ATA⁺ nuclear fraction. **(B)** REEAD assay for TOP1 activity. **(C)** REEAD assessing TOP1 inhibition by HC-IgG or ATAs from four individual SSc patients (P1-4). *n* = 5 per condition. **(D)** DNA relaxation assay with HC-IgG, ATAs-depleted IgG, or ATAs from eight SSc patients. **(E)** Nicked DNA fraction with pUC 19 plasmid vector, ATAs without TOP1 (negative controls), TOP1 with Campthotecin (CPT, 50 µM), HC-IgG, or ATAs from P1**-**4). **(F)** TOP1 activity by REEAD in endothelial cells nuclear fractions exposed to HC-IgG or ATAs (16 hours), normalized to protein concentration. NT: non-treated. *n* = 2 per condition. **(G)** TOP1cc and γ-H2AX in HC and ATA^+^ SSc skin biopsies (*n* = 3 per condition) and **(H)** in NativeSkin® explants after 7-day exposure (left); quantification at days 1, 3, 7 normalized to control (right). *n* = 3 per condition; *n* = 2 coverslips per condition. **(I)** COL1A1, α-SMA, and **(J)** picrosirius red collagen in NativeSkin® explants after 7-day exposure; collagen expressed as polarized positive/total area ratio. *n* = 3 (ATAs, HC-IgG), *n* = 2 (control), *n* = 6 regions per explant. IgG concentrations: 100 µg/mL (A); HC-IgG: 20 µg/mL, ATAs: 5-20 µg/mL (C); HC-IgG: 10 µg/mL, SSc-IgG: 5-10 µg/mL (D, F); ATAs 40 µg/mL (E); HC-IgG: 450 µg/explant, ATAs: 300 µg/explant (H-J). Box plots show median and full range. Ns: not significant; *P < 0.05, **P < 0.01, ***P < 0.001, ****P < 0.0001; Kruskal-Wallis with Dunn’s post-hoc test (C, E, J); one-way ANOVA with Tukey’s post-hoc test (H). Airyscan confocal (∼140 nm lateral resolution) for illustration; spinning disk for *ex vivo* quantification.

To directly probe intranuclear antibody-antigen interaction hypothesis, we progressed to examine whether ATAs could inhibit TOP1 enzymatic activity. The biological function of TOP1 includes DNA cleavage and ligation as well as a strand rotation step during which the DNA strand cleaved by TOP1 rotates around the intact strand to remove supercoils in the genome. We previously reported that the Rolling Circle–Enhanced Enzyme Activity Detection (REEAD) assay measure TOP1 cleavage and ligation activity (**Fig. 2B**) (*14*). ATAs inhibited the cleavage-ligation activity of TOP1 when measured *in vitro* by REEAD (**Fig. 2C**). We confirmed ATAs inhibition of TOP1 to be dose-dependent, while no inhibition was observed in the presence of ATAs-depleted IgG or HC-IgG (**Fig. S7A-D**). We then continued to examine the functional interaction between ATAs and TOP1 in a relaxation experiment, which measure the overall catalytic cycle of TOP1 (consisting of binding to the DNA, cleavage, strand rotation, ligation, and dissociation from the DNA), showing that ATA inhibited the TOP1-dependent conversion of supercoiled plasmid to relaxed plasmid (**Fig. 2D**). TOP1 functions by introducing transient single-strand breaks in the DNA through the formation of a short-lived TOP1-DNA cleavage complex (TOP1cc). We showed that ATAs stabilized the TOP1cc in a nicking experiment (**Fig. 2E, Fig. S7E**) consistent with the possible transient inhibition of the TOP1-mediated DNA ligation. However, the TOP1 functionality assays also show a patient-dependent variation.

After confirming that ATAs inhibit TOP1 activity *in vitro*, we investigated whether the nuclear fraction of ATA-exposed cells exhibited reduced TOP1 activity. Endothelial cells were cultured in the presence or absence of ATAs. Nuclear extracts were then prepared, and TOP1 activity in the extracted nuclear fraction was measured using the REEAD assay. This showed that nuclear TOP1 activity decreased when the cells were exposed to ATAs but not to HC-IgG. Moreover, using the RADAR (rapid approach to DNA adduct recovery) assay, we detected TOP1cc in DNA extracted from ATA-treated fibroblasts (**Fig. 2F, Fig. S7F-G**). This reduction was not due to diminished TOP1 protein levels, as western blot analysis showed no change in TOP1 abundance (**Fig. S7H**). Accordingly, we examined TOP1cc accumulation, DNA damage (γ-H2A histone family member X, γ-H2AX), and DNA repair responses (PARP-1) in cellular models. Treatment with either Topotecan (TPT), ATA^+^-IgG, and ATAs increased TOP1cc, γ-H2AX, and PARP-1 expression in cells compared to HC-IgG. Anti-DNA^+^-IgG also increased γ-H2AX expression in cells. Whole cellular PARP1 accumulation at early timepoints, followed by nuclear depletion at 72h as shown in proteomic, reflects a temporal shift from damage-induced recruitment under sustained TOP1cc-driven genotoxic stress (*15*) (**Fig. S8A-D**). In fibroblasts cultured from SSc patients, basal TOP1cc and PARP-1 levels were elevated and further increased upon ATAs exposure (**Fig. S8E**). We further investigated the TOP1cc and γ-H2AX presence in skin samples at basal state and their accumulation in skin explant models. In skin samples from ATA^+^ patients, we detected strong basal TOP1cc and γ-H2AX staining, which were similarly reproduced in cellular and skin explant models exposed to ATAs (**Fig. 2G-H**). As fibrosis is a key feature of SSc skin lesions, we examined whether ATAs could induce fibrosis in our skin explant model. In alignment with ATAs being biologically active, we observed increased α–smooth muscle actin staining (α-SMA) and type I collagen deposition in skin explants exposed to ATAs (**Fig. 2I-J, Fig. S9**). Collectively, these results support that ATAs alter TOP1 functions in the nucleus resulting in DNA damage and collagen deposition.

### Internalized Anti-Topoisomerase Antibodies Activate Innate Interferon Signaling

The IFN signature in CTDs is characteristic (*16*). Given the established link between DNA damage and TOP1cc formation (*17*, *18*), we progressed to examine if ATAs could lead to STING activation and hereof type I IFN (*19*). IFN-α pretreated endothelial cells transcriptomic analysis revealed a distinct clustering pattern after exposure to ATAs compared to HC-IgG (**Fig. 3A and B**). Treatment with ATAs resulted in 122 differentially expressed genes (DEGs) (p < 0.05) (**Fig. 3C**). When comparing HC-IgG to our none-treated control, only two genes were differentially expressed, supporting the idea that only ATAs are bioactive. The unique activation of cells by ATAs was also evident in the hierarchical clustering and pathway analysis, showing robust induction of type I IFN-related genes and enrichment of pathways mainly associated with interferon signaling. We further observed induction of IFN-related genes (*IFI44, IFIT5, XAF1, MX1*), nucleic acid sensing and regulation related genes (*OAS1, OAS2, OAS3, RIG, EIF2AK2*), along with tripartite motif (TRIM) family genes (*TRIM21*) (**Fig. 3D-E, Fig. S10A-D**). We confirmed that ATAs induce STING phosphorylation in a time-dependent manner, with maximal phosphorylation after 12h (**Fig. 3F**). Cells exposed to ATAs resulted in secretion of C-X-C motif chemokine ligand 10 (CXCL10), and production of bioactive type I IFN (**Fig. 3G-H, Fig. S10E**). The induction of CXCL10 was reduced in cyclic GMP-AMP synthase (cGAS)-deficient and completely inhibited in STING-deficient THP-1 cells (**Fig. 3I**). Lung explant models exposed to ATAs similarly exhibited IFNβ and CXCL10 synthesis (**Fig. 3J, Fig. S10F**). These findings show that nuclear ATA-TOP1 interactions activate the type I IFN pathway via cGAS-STING DNA sensing, connecting ATAs intranuclear activity to the observed type I IFN signature in SSc.

**Figure 3.**
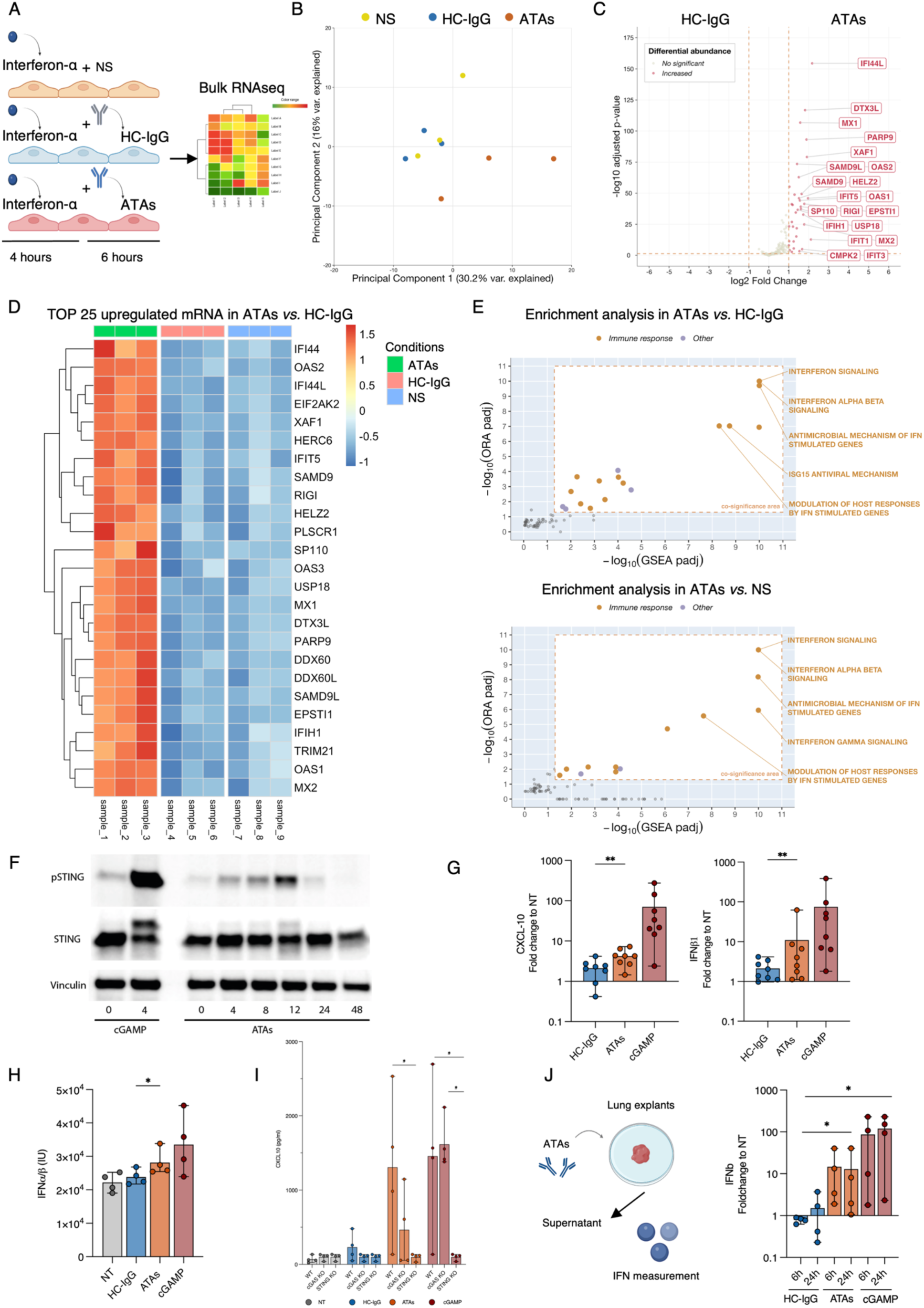
Internalized Antinuclear Antibodies Activate Innate Interferon Signaling. **(A)** Transcriptomic workflow. Endothelial cells pre-treated with IFN-α (4 hours) were exposed to ATAs or HC-IgG for 6 hours followed by bulk RNA sequencing. **(B)** PCA of mRNA profiles in ATAs- or HC-IgG-treated endothelial cells. *n* = 3 per condition. **(C)** Volcano plot of DEGs in ATAs versus HC-IgG (adjusted P < 0.05; |log₂ FC| > 1). **(D)** Heatmap of top 25 dysregulated mRNAs across ATAs-, HC-IgG-, and NS-treated cells (adjusted P < 0.05; |log₂ FC| > 1). **(E)** Pathway enrichment of mRNAs upregulated in ATAs versus HC-IgG; ORA (y-axis) and GSEA (x-axis, Reactome); five most significant labeled. **(F)** Western blot of p-STING in endothelial cells exposed to ATAs for 0-48 hours; cGAMP as positive control. **(G)** IFN-β (left) and CXCL10 (right) mRNA in endothelial cells exposed to HC-IgG or ATAs. *n* = 8 per condition. **(H)** Bioactive type I IFN in HEK-Blue IFN-α/β reporter cell supernatants treated with cGAMP, HC-IgG, or ATAs. *n* = 4 per condition. **(I)** CXCL10 protein in wild-type, cGAS-KO, and STING-KO THP-1 supernatants exposed to cGAMP, HC-IgG, or ATAs; attenuated in STING-KO cells. *n* = 4 per condition. **(J)** IFN-β mRNA in human lung explants exposed to HC-IgG, ATAs, or cGAMP for 6 or 24 hours. *n* = 4 per condition. NS: non-stimulated. Concentrations: 10 µg/mL HC-IgG and ATAs (all panels); cGAMP 50 µg/mL (F), 16 µg/mL (H-J). Box plots show median and full range. *P < 0.05, **P < 0.01; Kruskal-Wallis with Dunn’s post-hoc test (G, H, I, J).

### Endocytic Uptake and Intracellular Trafficking of Anti-Topoisomerase Antibodies

Building on evidence that ANAs engage their cognate nuclear antigens within target cells, we next investigated the intracellular trafficking steps that enable ANAs to reach the nucleus, beginning with endocytosis. Cold blockage of endocytosis showed a marked reduction in IgG fluorescent spots, indicating an active endocytic mechanism (**Fig. S11A**). Using pharmacological inhibitors and siRNA-mediated knockdown, we established that clathrin-mediated endocytosis was the dominant entry route for antibody internalization, whereas caveolin-mediated uptake played a minimal role (**Fig. 4A and Fig. S11B**).

**Figure 4.**
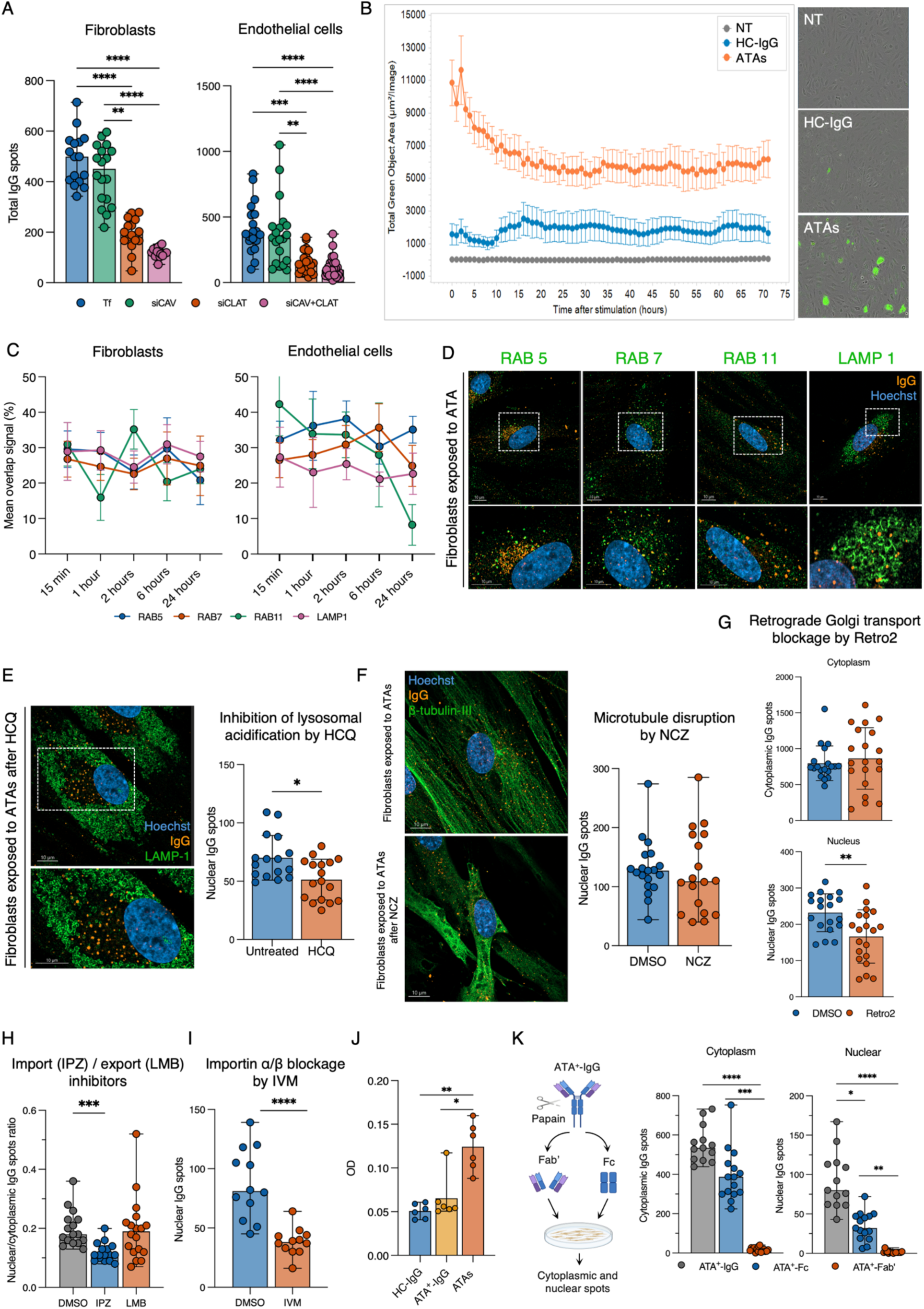
Endocytic Uptake and Intracellular Trafficking of Antinuclear Antibodies. **(A)** Intracellular IgG in fibroblasts (left) and endothelial cells (right) after caveolin (CAV), clathrin (CLAT), or CAV+CLAT siRNA knockdown; Tf: transfectant. **(B)** Incucyte time-lapse of fluo-HC-IgG and fluo-ATAs in endothelial cells after 24-hour pre-exposure; signal (green object area, µm²/image) monitored hourly for 72 hours. **(C)** ATAs colocalization with endosomal compartments in fibroblasts and endothelial cells at 15 minutes and 1, 2, 6, 24 hours; y-axis: proportion of overlapping spots. **(D)** ATAs in fibroblast endosomal compartments at 1 hour: Rab5 (early), Rab7 (late), LAMP-1 (lysosome), Rab11 (recycling). **(E)** Intranuclear IgG quantification and representative micrographs in fibroblasts exposed to ATAs after hydroxychloroquine (HCQ, 20 µM, 20 hours; lysosome staining: LAMP-1), and (**F**) nocodazole (NCZ, 10 µM, 1 hour; β-tubulin-III staining). **(G)** Cytoplasmic and nuclear IgG quantification in fibroblasts exposed to ATAs after Golgi apparatus retrograde transport blockage by Retro-2 (20 µM, 1 hour). **(H)** Nuclear-to-cytoplasmic ATA ratio after importazole (IPZ 20 µM) or leptomycin B (LMB 20 nM, 2 hours), and **(I)** intranuclear IgG after ivermectin (IVM). **(J)** Arginine content in HC-IgG, ATA⁺-IgG, and ATAs (OD; *n* = 6/condition). **(K)** Fc/Fab staining after papain cleavage of ATA⁺-IgG. *n* ≥ 100 cells per condition, *n* = 3 experiments for A, C, D, E, F, G, H, I, K. ATAs at 25 µg/mL (A, C-H, K) and 10 µg/mL (I). Box plots show median and full range. ns, not significant; *P < 0.05, **P < 0.01, ***P < 0.001, ****P < 0.0001; Mann-Whitney U test (E, F, G, I, K); Kruskal-Wallis with Dunn’s post-hoc test (A, H, J). Airyscan confocal (∼140 nm lateral resolution).

Time-lapse imaging revealed rapid intracellular trafficking. IgG reached intracytoplasmic and intranuclear maximum accumulation at 1h in endothelial cells and at 2h in fibroblasts (**Fig. S12A-B**). We confirmed that HC-IgG, SSc-IgG, and ATAs are recycled by cells, with continuous intracellular trafficking consistent with the endosomal route (**Fig. 4B and Fig. S12C-D**). To characterize this pathway, we assessed the colocalization of ATAs and endosomal compartments on confocal micrographs staining early endosomes (Rab5), late endosomes (Rab7), lysosomes (LAMP-1), and recycling endosomes (Rab11). In fibroblasts, co-distribution with early/late endosomal and lysosomal compartments remained stable over time, whereas recycling endosomes displayed a pronounced peak at 2 hours. In endothelial cells, early/late endosomal and lysosomal co-distribution remained globally stable, while recycling endosomes showed a maximum value at 15 min followed by a decline (**Fig. 4C-D**). Inhibition of lysosomal acidification altered partially intranuclear ATAs spots, suggesting that a proportion of ATAs can evade lysosomal degradation (**Fig. 4E, Fig. S13A).** We next examined the contribution of tubulin-dependent transport, hallmark of endosomal route. ATAs showed cytoplasmic colocalization with β-tubulin-III and proximity in nanoscopy, but tubulin polymerization blockade did not reduce nuclear ATAs accumulation (**Fig. 4F, Fig. S13B-C**). No colocalization was detected between ATA and the mitochondria or endoplasmic reticulum. Blocking retrograde Golgi transport partially reduced intranuclear ATAs staining (**Fig. 4G, Fig. S13D-E**). Galectin-9 staining, performed to detect endosomal membrane damage, indicated low-level endosomal escape (**Fig. S13F**). Together, these findings reveal that while ATAs primarily follow the canonical endosomal route to the nucleus, a subset escapes endosomal confinement and traffic independently without detectable endosomal damage.

Because nuclear import is an active process requiring importins, nuclear pore complexes, and import machinery (*20*, *21*), we tested the effect of nuclear transport inhibitors. Blocking importin function significantly reduced intranuclear fluorescent spots following ATAs exposure, whereas blocking nuclear export did not, confirming active import-mediated nuclear trafficking in cells (**Fig. 4H-I, Fig. S14A-D**). This aligned with the ATAs, showing high arginine levels essential for importin-mediated transport (*20*, *22*) (**Fig. 4J).** Finally, to determine structural requirements for ANA nuclear delivery, we monitored uptake of isolated Fc and Fab’ fragments from ATA^+^-IgG. Compared to full ATA^+^-IgG, Fc fragments displayed reduced nuclear localization but preserved cytoplasmic uptake, whereas Fab’ fragments showed reduced cytoplasmic uptake and no nuclear localization (**Fig. 4K, Fig. S14E**). These findings indicate that ATAs trafficking to the nucleus requires the Fc region, while nuclear entry additionally depends on importin transfer.

### Neonatal Fc Receptor is Required for Intracellular Trafficking and Nuclear Access of Anti-Topoisomerase Antibodies

FcRn enables recycling of endocytosed IgG, explaining its long half-life. FcRn resides predominantly in the endosomal compartment and is broadly expressed across both immune and non-immune cells, including endothelial and stromal cells (*23*, *24*). We investigated its role in ANA trafficking in fibroblasts and endothelial cells. FcRn was expressed in SSc and HC skin samples (**Fig. S15A**). FcRn silencing by siRNA markedly reduced intracellular ANA accumulation, implicating FcRn as the critical transporter (**Fig. 5A, Fig. S15B-C**). The time-course analysis showed that ATAs and FcRn were already co-distributed at 15 min, with values of approximately 30-50% that remained broadly stable over 24h (**Fig. S15D**). PLA revealed both cytoplasmic and nuclear ATA-FcRn interaction in cells and skin explants exposed to ATAs (**Fig. 5B-C**). Structural interaction was explored by nanoscopy which clearly revealed ATA attachment to FcRn (**Fig. 5D**). To confirm the central role of FcRn in the nuclear uptake of ANAs and its downstream consequences, we employed two clinically approved anti-FcRn agents (efgartigimod and nipocalimab) to our models. In cells treated with anti-FcRn agents prior to ATA exposure, intranuclear IgG spots, TOP1cc, and γ-H2AX staining were all markedly reduced compared to untreated cells (**Fig. 5E-G, Fig. S16**). Consistently, efgartigimod treatment in skin explants exposed to ATAs diminished intranuclear IgG signal intensity, TOP1cc and γ-H2AX as well as α-smooth muscle and type 1 collagen staining and collagen deposition (**Fig. 5I-N, Fig. S17)**.

**Figure 5.**
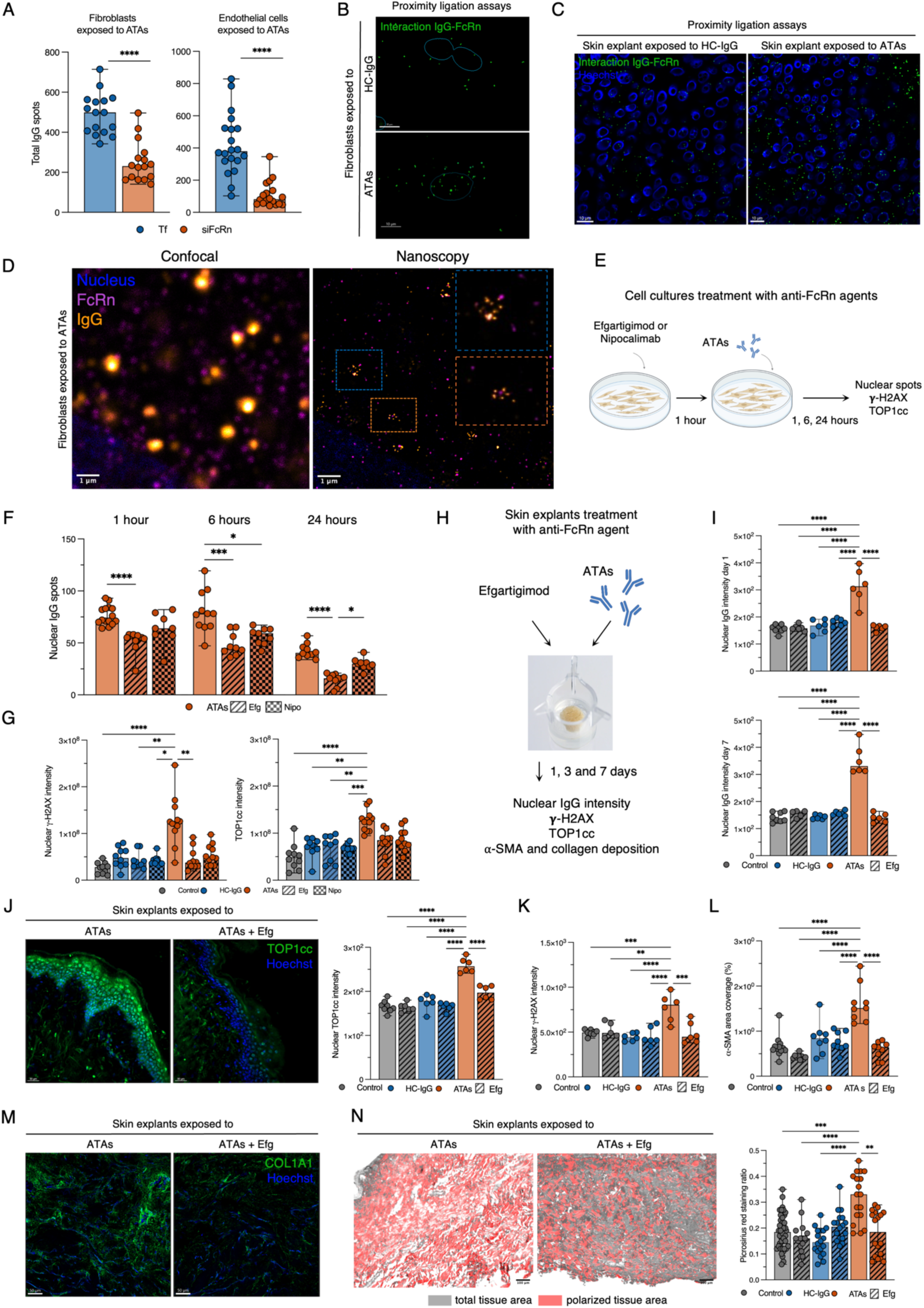
FcRn Governs Intracellular Trafficking and Nuclear Access of ANA. **(A)** Intracellular IgG in fibroblasts and endothelial cells after FcRn siRNA knockdown (*n* ≥ 100 cells; *n* = 3 experiments). **(B)** FcRn-IgG PLA in fibroblasts exposed to HC-IgG or ATAs (1 hour) and **(C)** in NativeSkin® explants (ATAs, 300 µg/explant, 24 hours). **(D)** STED of IgG/FcRn in fibroblasts exposed to ATAs (1 hour). **(E)** Experimental design of cell cultures pretreated with Efgartigimod (Efg, 25 µg/mL) or Nipocalimab (Nipo, 10 µg/mL) for 1 hour before HC-IgG (100 µg/mL) or ATAs (25 µg/mL) for 24 hours. **(F)** Intranuclear IgG quantified at 1, 6, 24 hours (*n* = 3 experiments, *n* = 100 cells/experiment) and **(G)** γ-H2AX/TOP1cc after 24 hours (*n* ≥ 100 cells, *n* = 3 experiments). **(H)** NativeSkin® explants treated with culture medium (control) or HC-IgG (450 µg/explant) or ATAs (300 µg/explant) ± Efg (10 µg/explant) for 1, 3, 7 days. **(I)** Intranuclear IgG at days 1 and 7, **(J)** TOP1cc (with micrographs), **(K)** γ-H2AX, and **(L)** α-SMA coverage. (**M**) Type 1 collagen staining (COL1A1) at day 7 in NativeSkin® explants (*n* = 3 samples/condition; *n* = 2 coverslips/sample). **(N)** Picrosirius red staining (left) and collagen quantification (right) in NativeSkin® explants (HC-IgG or ATAs, 7 days); polarized positive/total area ratio (*n* = 3 samples/condition; *n* = 6 regions/sample). Box plots show median and full range. ns, not significant; *P < 0.05, **P < 0.01, ***P < 0.001, ****P < 0.0001; Kruskal–Wallis with Dunn’s post-hoc test (A, F, G and M); one-way ANOVA with Tukey’s post-hoc test (I-L). Airyscan confocal (∼140 nm lateral resolution) for cells; spinning disk for explants.

## Discussion

The longstanding view that ANAs are non-pathogenic has been rooted in a simple premise: antibodies cannot exert biological effects on antigens that are within the cell. Our findings directly overturn this assumption. We demonstrate that ANAs can access living cells, accumulate in nuclei, and engage their cognate antigens in a biologically meaningful manner. In doing so, this work reframes ANAs not as passive biomarkers of immune dysregulation but as active intracellular effectors capable of directly shaping cellular responses and tissue pathology (**Fig. S18**).

Using SSc and ATAs as a model, we uncover a direct mechanistic link between autoantibody specificity and intracellular dysfunction. Nuclear ATAs inhibit TOP1 enzymatic activity, induce DNA damage, and elicit robust activation of type I IFN signature, using the cGAS-STING pathway. These effects provide a unifying explanation for two central features of CTDs: the tight association between specific ANAs and distinct clinical phenotypes, and the pervasive activation of a type I IFN-driven transcriptional program. In this framework, type I IFN production is not merely a downstream consequence of inflammation but an intrinsic response to intracellular autoantibody-autoantigen engagement.

A central implication of this work is that intracellular antigen localization does not represent an immunological safe harbor. Instead, we show that antibody access to nuclear compartments is enabled by regulated intracellular immunoglobulin trafficking. Identification of FcRn as a key determinant of ANAs nuclear routing reveals a novel intracellular role for a receptor classically viewed as a mediator of IgG recycling and homeostasis (*24*). This finding expands the conceptual scope of FcRn biology and suggests that cellular entry alone is insufficient for pathogenicity, as downstream effects require productive nuclear antigen engagement.

Beyond SSc, these findings have broader implications for CTDs. Disease-specific autoantibodies have long been associated with distinct patterns of organ involvement and prognosis. Recent data suggested that ANAs could define molecularly homogeneous patient subsets (*25–27*). Our results propose a mechanistic basis for this association: intracellular engagement of specific nuclear autoantigens by ANAs may directly program cell-type–specific responses and immune activation associated with nucleotide function. In this context, autoantibody specificity becomes a central determinant of intracellular signaling pathways rather than just a disease marker. A key finding is that ATAs purified from different patients varied in their capacity to induce cell penetration, TOP1 inhibition, TOP1cc stabilization and IFN production, reflecting the substantial heterogeneity in clinical outcomes observed in SSc.

More broadly, this work challenges the traditional compartmentalization of antibody function as exclusively extracellular. By demonstrating that circulating autoantibodies can access intracellular targets and exert direct nuclear effects, we reveal a previously unrecognized dimension of antibody-mediated immunity. This intracellular effector function offers a new perspective on how humoral autoimmunity leads to tissue damage and disease variability in CTDs. Furthermore, it prompts consideration of intracellular humoral immunity in more general terms, explaining the importance of this response against intracellular pathogens (*28*). These findings support a revised model of humoral immunity in which antibodies can function across both extracellular and intracellular compartments.

## Data Availability

All data produced in the present study are available upon reasonable request to the authors. The code and raw data used to generate omics analysis and visualizations are available at https://github.com/j-sottiaux/chepy_2026_fig2_fig3

## Acknowledgments

We thank Solène Audry for her assistance with patient skin sectioning, and Sarah Gabut and Yassine Jarmoni for their assistance with cellular imaging, including virtual slide scanning and spinning disk microscopy mosaic acquisition. We thank the OrganOmics Core Facility of the University of Lille, which is accredited by the GIS IBiSA and is a member of the French Proteomics Infrastructure (ProFI) UAR 2048. We are grateful to Prof. Per Glud Ovesen for technical support and Dr. Amélie Bonnefond for her critical review and constructive suggestions on this manuscript.

## Funding

Inserm and the French Ministry of Health in the context of MESSIDORE 2023 call operated by IReSP (AAP-2023-MSDR-341423) (VS)

CPER ResIsT-omics (n◦22-C-0128) (DL)

FHU PRECISE (Univ. Lille, CHU Lille, Inserm) (DL)

Association des Sclérodermiques de France (Grant 2021) (VS)

Société Nationale Française de Médecine interne (Grant 2022) (AC)

CSL Behring (Research grant 2024) (VS)

Groupe Francophone de Recherche sur la Sclérodermie (Grant 2024) (AC)

French government through the Programme Investissement d’Avenir (I-SITE ULNE / ANR-16-IDEX-0004 ULNE) managed by the Agence Nationale de la Recherche (n°I-KUL-22-005-ARCHIE-INFINITE) (VS)

Foundation for Research in Rheumatology (MA)

The Danish Rheumatoid Association (MA, CT, BD, SRG)

Aarhus University (MA, CT)

Karen Elise Jensens Mindefond (SRG)

AUFF NOVA AUFF-E-2025-9-11 (SRG)

LEO Foundation LF-FE-24-700026 (SRG)

The Aarhus University Research Foundation: AUFF-E-2016-9-27 (BD)

Gilead Nordic Fellowship Grants (BD)

Grosserer Valdemar Foersom og hustru Thyra Foersons fond (BRK)

Grosserer L.F. Foghts Fond (BRK)

Independent Research Fund, Denmark DFF-4285 (BD)

Danish National Research Foundation DNRF164 (SRP)

## Author contributions

Conceptualization: AC, CT, MEM, MA, SD, SG, DL, MT, BD, VS

Methodology: AC, CT, MEM, MA, SV, JHM, EmH, EN, JS, MD, KS, MM, MS, MH, JGK, CC, LG, EGJ, CBFA, AB, MS, BRK, SD, SG, DL, CT, MT, BD, VS

Investigation: AC, CT, MEM, MA, SV, JHM, EmH, JS, MD, SBP, MM, MS, JGKS, LG, EGJ, AB, BRK, SD, SG, DL, MT, BD, VS.

Visualization: AC, CT, MEM, MA, SV, JS, MD, CC, SG, MT, BD, VS

Funding acquisition: AC, ErH, SG, BRK, DL, BD, VS Project administration: AC, SG, BRK, MT, BD, VS

Supervision: AB, BRK, CT, JHM, DA, CBFA, SG, MT, BD, VS

Writing – original draft: AC, CT, MEM, JHM, CBFA, BRK, MA, SG, MT, BD, VS

Writing – review & editing: AC, CT, MEM, MA, SV, JHM, EmH, SMM, EN, JS, MD, KS, SBP, MM, MS, MH, JGK, DA, ElH, CC, LG, EGJ, SRP, CBFA, AB, ErH, MS, JMK, BRK, SD, SG, DL, CT, MT, BD, VS

## Data and materials availability

The code and raw data used to generate omics analysis and visualizations are available at https://github.com/j-sottiaux/chepy_2026_fig2_fig3.

## Supplementary Materials

## Materials and Methods

### Patients, sera and skin

For Lille cohort, sera from patients (SSc and SLE) were obtained from the FHU PRECISE biobank of the Department of Internal Medicine, Lille University Hospital (NCT04334031, ethical approval: CPP N°2019-A01083-54). Sera from healthy controls (HC) were obtained from the Etablissement Français du Sang. For Aarhus cohort, blood samples were obtained from patients enrolled at the Department of Rheumatology, Aarhus University Hospital (approval by the Regional Ethics Committee (Reference number 700779) and Region Midt’s Data Protection Agency (1-16-02-542-20)). Serum was collected from SSc patients (*n* = 16) and age- and gender-matched HC obtained from the Danish blood bank, Denmark (HC, *n* = 8).

### Skin biopsies

Skin samples from SSc patients were obtained by forearm biopsy from treatment-naive SSc patients who were ATA^+^ or ARA^+^. HC skin samples were obtained from reconstructive surgery after bariatric surgery (CPP N°2021-A03137-34). SSc and HC skin samples were embedded in optimal temperature compounds and stored at -80°C.

### Immunoglobulins G and antibodies

#### Total IgG purification

Total IgG were purified from patients or HC sera by affinity chromatography on an ÄKTA-start FPLC system using HiTrap Protein G columns (Cytiva, Marlborough, MA, USA). Total IgG-level measurement and serotype assay were performed by Immunoturbidimetry (Optilite® Analyzer, Binding Site, Thermo Fisher Scientific, Waltham, Massachusetts, USA) and BioPlex ANA screen (BioPlex 2200 ANA Screen, Bio-Rad Laboratories, Hercules, CA, USA), according to the manufacturer’s instructions to ensure the validity of the purification step.

#### Construction of protein fragments

Guided by X-ray structural data deposited in the Protein Data Bank, we designed three constructs for recombinant expression in E. coli (**Fig. S19A**). TopI_aa. 213-429, aa. 434-765Δ633-714 and aa. 639-765 were all synthesized using Genscript and inserted into the MCS of p.ET15b. These contained 13 mutations in the DNA-binding residues that allow expression in the bacterial cytoplasm. The following twelve residues were mutated to alanine: p.R364, p.K374, p.T411, p.K425, p.K443, p.N491, p.K493, p.T501, p.K532, p.D533, p.N574, and p.K587. In addition, the enzymatically important Y723 residue was mutated to phenylalanine. The resulting three expression vectors were transformed into E. coli BL21 Rosetta and plated on LB agar. All LB media contained 100 µg/ml ampicillin and 34 µg/ml chloramphenicol. Overnight cultures were prepared by inoculation with the LB medium. One millilitre of each overnight culture was inoculated into 500 ml fresh LB medium and grown at 37°C in a shaking incubator. Expression was induced with 0.2 mM Isopropyl β-d-1-thiogalactopyranoside (IPTG) when the cells reached an OD600 of 0.6-0.9. After overnight expression at 20°C, cells were harvested by centrifugation at 4,000 × g for 20 min. Pellets were resuspended in 20 mM imidazole, 20 mM Tris-HCl (pH 7.5), and 500 mM NaCl and lysed by sonication (Branson Sonifier 250). Lysates were cleared by centrifugation for 20 min at 25,000 g. The supernatants were subjected to nickel-affinity chromatography on HisTrap FF Crude columns (GE Healthcare), and bound proteins were eluted with a linear gradient of 20-500 mM imidazole in 20 mM Tris-HCl (pH 7.5) and 500 mM NaCl. Size-exclusion chromatography on a Superdex 200 Increase 10/300 GL column (GE Healthcare) equilibrated in 10 mM HEPES, pH 7.5, and 150 mM NaCl was used as the final step of purification. Protein purity was examined by SDS-PAGE on 4-15% TGX stain-free gels (Bio-Rad). Relevant fractions were pooled and either stored at -20 °C or dialyzed against 0.1 M NaHCO_3_ pH 8.3 and 500 mM for the generation of affinity columns for purification of ATAs from total IgG.

#### Immunoglobulin, anti-topoisomerase I antibodies and depleted fraction purification

Protein concentration was measured by absorption at 280 nm using a NanoDrop ND-1000 spectrophotometer (NanoDrop Technologies, Wilmington, DE, USA) and an extinction coefficient of 13.5 (1% IgG solution).

Topoisomerase fragments were coupled to CNBR-activated Sepharose 4 B medium, according to the manufacturer’s instructions (Cytiva) at a density of 1 mg/ml of swelled medium and packed into small columns for gravity flow purification. Total IgG from the protein G affinity purification described above was then applied to this column, and the flow-through was collected. Following thorough washing, bound IgG was eluted with a 0.1 M glycine pH 2.5 buffer and immediately neutralized with 1 M Tris pH 9.0. One millilitre of IgG and ATAs were dialyzed against 3x500 ml of PBS, pH 7.4, to remove the glycine. IgG were tested for endotoxin levels by Lumulus Amebocyte Lysate endotoxin quantification to be < 0.01 EU/ml (Pierce, UK). Fractions collected during the purification process were analyzed after separation on SDS-PAGE (**Fig. S19B**) and Western blotting (**Fig. S19C**).

#### Affinity testing of ATA by surface plasmon resonance

Surface plasmon resonance (SPR) experiments were carried out on a Biacore 3000 using 10 mM HEPES (pH 7.5), 150 mM NaCl, 2 mM CaCl_2_, and 0.05% Tween 20 as the running buffer. A surface for IgG capture was prepared by amine coupling of recombinant protein G (08062, Merck) on a CM5 chip, according to the manufacturer’s instructions (Cytiva). In short, surfaces were activated by a 7 min injection of a 1:1 mixture of 0.1 M NHS (N-hydroxysuccinimide) and 0.4 M EDC (3-(N,N-dimethylamino) propyl-N-ethylcarbodiimide). Protein G was immobilized at 30 µg/ml in 10 mM sodium acetate (pH 4.5). The residual reactive groups were blocked by a 7 min injection of 1 M ethanolamine at pH 8.5. Binding analysis was performed at 25 °C and data were collected at a rate of 1 Hz. Using BIAevaluation 4.1.1 software (Cytiva), the recorded signals from the active flow cell were double-referenced, that is, the signal from the in-line reference flow cell was subtracted as was the signal from a blank run (0 nM analyte). For analysis of purified ATAs, IgG was captured on the protein G surface at a density of approximately 1,500 RU, followed by an injection of 200 nM of purified recombinant topoisomerase fragments. Surfaces were regenerated at the end of each cycle by a 30 s injection of 10 mM glycine (pH 1.5) (**Fig. S19D**).

#### Functionality assays of commercial polyclonal ATAs

In addition, we used commercially sourced and pooled polyclonal ATAs (ATAs, TopoGEN, Buena Vista, Colorado, United States) to enable large-scale experiments and to ensure that the purification method did not influence our results. To verify that the polyclonal ATAs preparation was representative of patient-derived ATAs, TOP1 inhibitory activity of polyclonal ATAs was further confirmed by DNA relaxation assay (**Fig. S19E**). ATA reactivity was verified by two orthogonal approaches: indirect immunofluorescence on HEp-2 cells (**Fig. S19F**), using the ICAP-defined “DNA topoisomerase I (topo I)-like” (AC-29) pattern as a reference (*29*), and antigen-specific detection by multiplexed bead-based immunoassay (BioPlex).

#### Immune complex formation assay

Immune complexes were prepared by incubating ATAs with 5 µg/mL recombinant human TOPI protein (MBS8122098, MyBioSource, San Diego, CA, USA) in culture medium for 30 minutes at room temperature. The mixture was then filtered using 300 kDa filter unit (Vivaspin® 300 KD concentrateur, Sartorius, Göttingen, Germany) to filter unbound antibodies. Cells were subsequently incubated with the resulting unfiltrated fraction for 1h at 37°C, followed by processing according to the standard immunofluorescence protocol described below in the immunofluorescence section.

#### Arginine content in immunoglobulin (Sakaguchi test)

Purified HC-IgG, SSc-IgG and ATAs were diluted in 40% NaOH 1% (3:1). 1-naphthol (Sigma-Aldrich, 90-15-3) diluted in alcohol was added to the solution. Hereafter, 10 µl of 10% sodium hypobromite solution (Sigma-Aldrich, 71329) was added, and the color change was immediately examined at optical density of 510 nm.

#### Fab’ and Fc fragmentation

ATA^+^-IgG were fragmented into Fab’ and Fc components using immobilized papain (Pierce™ Fab Micro Preparation Kit, Thermo Fisher Scientific, Waltham, Massachusetts, USA), following the manufacturer’s instructions. Briefly, IgG were buffer exchanged into digestion buffer, incubated with papain resin at 37°C for 6 h with continuous mixing and centrifuged to recover the digest. Fab’ fragments were isolated by negative selection using Protein A spin columns; Fc and undigested IgG were recovered by low-pH elution and neutralized. Protein concentrations were determined by A280 using an extinction coefficient of 1.4. For each subject, 250 µg of purified IgG were fragmented, and resulting Fab’ and Fc fractions were used at 100 µg/mL for immunofluorescence assay.

#### IgG fluorescent clicking (Fluo-ATAs & Fluo-HC-IgG)

ATAs and HC-IgG were site-specifically labeled using the SiteClick™ Antibody Azido Modification Kit (Thermo Fisher Scientific, Waltham, Massachusetts, USA) followed by copper-free click conjugation to Alexa Fluor™ dyes via SiteClick™ sDIBO Alkyne reagents, according to the manufacturer instructions. Briefly, 1 mg of ATAs or HC-IgG was first concentrated and buffer-exchanged using centrifugal filtration. The Fc region carbohydrate domains were enzymatically modified by sequential incubation with β-galactosidase and β-1,4-galactosyltransferase in the presence of UDP-GalNAz, introducing azide moieties at terminal GlcNAc residues without affecting antigen binding. The azide-modified antibody was then reacted overnight at 25°C with an excess of Alexa Fluor™ 488 sDIBO Alkyne (Thermo Fisher Scientific, Waltham, Massachusetts, USA) to achieve site-specific labeling via strain-promoted azide–alkyne cycloaddition. Excess dye was removed by centrifugal purification, and the labeled antibodies were stored at 4°C in the dark until use.

### Cell culture

#### Endothelial cells and fibroblasts culture

Human umbilical vein endothelial cells (HUVECs, CC-2519, Lonza, Basel, Switzerland) were cultured in M199 culture medium (Thermo Fisher Scientific, Waltham, Massachusetts, USA) supplemented with 1.8% antibiotics (penicillin and streptomycin, Invitrogen, Thermo Fisher Scientific, Waltham, Massachusetts, USA), 0.9% GlutaMAX™ 100X (Invitrogen, Thermo Fisher Scientific, Waltham, Massachusetts, USA), 20% fetal calf serum (FCS, Lot RH20220001, Cytiva, Marlborough, Massachusetts, USA) and 10% EBM-2 medium supplemented with EGM-2 SingleQuots kit (endothelial growth media, Lonza, Basel, Switzerland) and used between 2th and 9th passage. Primary dermal human fibroblasts (ATCC® Number: PCS-201-012™, Manassas, Virginia, USA) between the 5th and 11th passage were cultured with Dulbecco’s Modified Eagle medium (Invitrogen, Thermo Fisher Scientific, Waltham, Massachusetts, USA) combined with antibiotics at 1% concentration (penicillin and streptomycin, Invitrogen, Thermo Fisher Scientific, Waltham, Massachusetts, USA) and 10% FCS. The STING ligand 2′3′-cGAMP (Invivogen) was utilized at a concentration of 16 µg/ml as a positive control. For PCR studies, cells were lysed, and RNA was extracted after 8 hours of stimulation. For ELISAs, supernatants were harvested after 24 hours of stimulation with autoantibodies and controls. To measure TOP1 activity in nuclear extracts, the cells were first washed twice and subsequently trypsinised. These were once again washed three times to remove extracellular ATAs. Cell pellets were resuspended in lysis buffer (0.1% NP-40, 10 mM Tris pH 7.9, 10 mM MgCl_2,_ 15 mM NaCl, 250 µg/mL β-glycerophosphate, 1x IP (Roche, 05892970001), 1 µM phenylmethylsulfonyl fluoride (PMSF), 1 µM DTT) at 4°C for 10 min. Pellets were created by centrifugation at 4°C at 2000 RPM for 5 min, followed by the removal of supernatant. The pellets were not disturbed when 100 µL extraction buffer (0.5 M NaCl, 20 mM HEPES pH 7.9, 20% glycerol, 250 µg/mL β-glycerophosphate, 1x IP, 1 µM DTT) was added. The pellets were rotated at 4°C for 1h, with the addition of 1 µM PMSF every 15 min. The samples were centrifuged at 15,000 RPM for 15 minutes at 4°C. The supernatant was used for Rolling circle Enhanced Enzyme Activity Detection (REEAD) assay. For proteomic analysis by LC-MS/MS, to avoid interference due to the presence of FCS, cells were FCS-deprived for 24 h after seeding. Fibroblasts were then cultured for 72 h in the presence of HC-IgG and SSc-IgG (100 µg/mL, *n* = 10 for HC and SSc serotypes). Cytoplasmic and nuclear fractions were separated using the manufacturer’s instructions (NE-PER™ Kit, Thermo Fisher Scientific).

#### SSc patients’ fibroblasts

Dermal fibroblasts from ATA^+^ and ACA^+^ SSc patients (*n* = 2 individual SSc patients) were isolated from forearm skin biopsies and cultured in Dulbecco’s Modified Eagle medium combined with antibiotics at 1% concentration (penicillin and streptomycin) and 10% FCS. Cultured cells were then used between the 5th and 11th passages and processed according to our standard immunofluorescence assay.

#### CaCo2 cells cultured

Caco2 cells were obtained from the American Type Culture Collection (ATCC) and cultured in Minimal Essential Medium (MEM) supplemented with 20% fetal bovine serum (FBS), 1% non-essential amino acids (NEAA), 100 units/mL penicillin, and 100 mg/mL streptomycin (Sigma-Aldrich ApS, Søborg, Denmark).

#### THP-1 cell lines

THP-1 WT, *STING* knockout (KO) (Cat. (thpd-kostg)) and *cGAS* KO (Cat. (thpd-kocgas) cells were obtained from InvivoGen. Cells were cultured in RPMI culture medium containing 10% FBS+ 1% penicillin/streptomycin and 70% RPMI. 200,000 cells were seeded onto 48 well plates and differentiated with 20 ng/ml PMA into macrophage-like cells. These cells were rested for 24 hours before stimulating them with ATAs, HC-IgG (10 µg/mL) or cGAMP (16 µg/mL).

### Tissue explant models

#### Lung explants

Lung tissue was acquired from patients undergoing lobectomy due to lung cancer (Request number 106/2025 from The Central Denmark Region Committees on Health Research Ethics). Biopsies of lung tissue of 20-30 mg were cultured in RPMI culture medium containing 10% FBS+ 1% penicillin/streptomycin at 37°C in a humidified atmosphere of 5% CO_2_. The lung tissue was stimulated with either HC-IgG, ATAs (10 µg/mL), or cGAMP (16 <u>µ</u>g/ml) in triplicats for 6h and 24 h.

#### Human skin explants

NativeSkin® human skin explants (11 mm diameter) were obtained from Genoskin (Toulouse, France). The explants originated from two healthy female donors (age range: 36-40 years) and were maintained *ex vivo* according to the manufacturer’s recommendations.

Upon receipt (day 0), explants were intradermally injected with either 300 µg of ATAs, 450 µg of HC-IgG, both diluted in culture medium for a total volume of 10 µL, or 10 µL of culture medium only as controls. Following injection, explants were returned to culture for 1 day, 3 days, or 7 days under standard conditions, with culture medium replaced daily. At each predefined time point, explants were cut, embedded in OCT compound and snap frozen in ice-cold isopentane, before storing at -80°C.

### Endocytosis inhibition assays

#### Cold blockage

Cells were pre-cooled at 4°C for 15 min, then incubated with ATAs for 1h at 4°C. Following incubation, samples were rinsed twice with ice-cold PBS and fixed with paraformaldehyde 4% (PFA) before proceeding to the standard immunofluorescence protocol.

#### Pharmacological inhibition

To assess the role of endocytosis in antibody uptake, endothelial cells and fibroblasts were pre-treated with pharmacological inhibitors targeting distinct endocytic pathways. Cells were incubated for 30 minutes at 37°C with chlorpromazine (10 µM; clathrin-mediated endocytosis inhibitor; MilliporeSigma, Burlington, Massachusetts, USA), genistein (50 µg/mL; caveolin-dependent endocytosis inhibitor, MilliporeSigma, Burlington, Massachusetts, USA), or dynasore (40 µM; dynamin-dependent endocytosis inhibitor, MilliporeSigma, Burlington, Massachusetts, USA). Following pre-treatment, ATAs were added to culture medium for 30 minutes and cells were processed according to our standard immunofluorescence protocol. As internal controls for inhibitor efficacy, AF488-conjugated transferrin (25 µg/mL; Thermo Fisher Scientific, Waltham, Massachusetts, USA) and AF488-conjugated cholera toxin subunit B (CTB; 5 µg/mL; Thermo Fisher Scientific, Waltham, Massachusetts, USA) were added to parallel samples during the inhibitor incubation period. All inhibitors and fluorescent probes were freshly prepared, and appropriate DMSO controls were included.

### Rapid Approach to DNA Adduct Recovery (RADAR) assay

RADAR assay was performed as detailed by Meroni *et al.* (*30*). Cells (20-40 × 10⁵ per well from a 6 well-plate, all 6 wells pooled per condition) were treated with topotecan (2 µM, Bio-techne, Minneapolis, Minnesota, USA), ATAs (25 µg/mL), or unstimulated, in FCS containing medium for 24 hours. Following treatment, media were aspirated, and cells were lysed with DNAzol^TM^ (Invitrogen, Thermo Fisher Scientific, Waltham, Massachusetts, USA). Lysates were collected, and DNA-protein covalent complexes were precipitated by adding ½ volume of cold 100% ethanol. Samples were incubated at -20 °C for 10 min, pelleted by centrifugation, and washed twice with 70% cold ethanol. Pellets were briefly air-dried and resuspended in 8 mM NaOH, keeping samples on ice. DNA concentration was measured using a Nanodrop and samples were normalized to 10 ng/µL. Normalized DNA was loaded onto nitrocellulose membranes using a slot blot apparatus (Bio-Dot®, BioRad, Hercules, CA, USA), blocked with 3% milk in TBS + 0.1% Tween-20, and probed with rabbit anti-topoisomerase antibodies overnight at 4°C. Membranes were washed, incubated with a goat anti-Rabbit IgG Alexa Fluor™ Plus 800 secondary antibody, and scanned using an iBright^TM^ FL1500 imaging system (Thermo Fisher Scientific, Waltham, Massachusetts, USA).

### DNA damage induction assay

Fibroblasts and endothelial cells were incubated with HC-IgG, ATA^+^-IgG (100 µg/mL) and ATAs (25 µg/mL) in FCS containing medium for 24 hours to evaluate DNA damage induction. Topotecan (2 µM, Bio-techne, Minneapolis, Minnesota, USA) was used as a positive control. Following treatment, cells were stained for TOP1cc, γH2AX, and PARP1 as described in the Immunofluorescence section.

### IgG recycling assay

The IgG recycling assay was adapted from Grevys *et al.* (*31*). Fibroblasts and endothelial cells were plated in 6-well-plates at a density of 60,000 cells per well and cultured for 72h. Cells were FCS-deprived for 24h before antibody incubation. After 24h deprivation, cells were incubated with 100 µg/mL of HC-IgG or SSc-IgG for 1h at 37°C. The medium containing IgG was removed, and cells were washed twice with PBS. Pre-warmed medium was added to the cells and incubated for 60 min at 37°C to allow IgG recycling. The concentrations of IgG in supernatants after incubation and recycling were measured with an IgG Human ELISA Kit (Thermo Fisher Scientific, Waltham, Massachusetts, USA), following the manufacturer’s instructions.

### Intracellular trafficking inhibition

Microtubule dynamics were disrupted by treatment with nocodazole (10 µM, MilliporeSigma, Burlington, Massachusetts, USA) for 1h at 37 °C, followed by incubation with ATAs for an additional 1h in the continued presence of nocodazole. Endosomal acidification and maturation were inhibited by treatment with hydroxychloroquine (20 µM, MilliporeSigma, Burlington, Massachusetts, USA) for 20 h at 37 °C, after which cells were incubated with ATAs (25 µg/mL) for 1h in hydroxychloroquine-containing medium. Retrograde transport was inhibited using Retro-2 (20 µM, MilliporeSigma, Burlington, Massachusetts, USA), applied for 1h prior to ATAs addition, followed by a 1h incubation in the continued presence of the inhibitor. Control cells were treated with DMSO or water under identical conditions.

### Pharmacological FcRn inhibition

For pharmacological inhibition of FcRn in cellular models, cells were cultured as described above and pretreated for 1h with either efgartigimod (25 µg/mL, Neo Biotech, Nanterre, France) or nipocalimab (10 µg/mL, Neo Biotech, Nanterre, France). Following pretreatment, ATAs (25 µg/mL) was added in fresh culture medium containing the corresponding inhibitor at the same concentration and cells were incubated for the indicated durations (1-24 h) before processing according to the immunofluorescence workflow described below. Control cells received equivalent volumes of culture medium with ATAs but without inhibitor.

For *ex vivo* experiments, skin explants were injected upon arrival with either 10 µg efgartigimod or an identical volume of culture medium. A second injection containing 300 µg ATAs, 450 µg HC-IgG, or culture medium alone was then administered. Accordingly, explants were assigned to control, efgartigimod-only, ATAs-only, HC-IgG-only, efgartigimod + ATAs, or efgartigimod + HC-IgG groups. Explants were subsequently returned to culture and maintained for 1, 3, or 7 days prior to downstream analyses as described below. Each treatment group consisted of three independent explants per time point (*n* = 3), while control groups consisted of four independent explants per time point (*n* = 4).

### Proximity Ligation Assays

Protein-protein interactions were detected using proximity ligation assay (PLA) with the Duolink® in situ PLA kit (MilliporeSigma, Burlington, Massachusetts, USA) according to the manufacturer’s instructions. In summary, fixed and permeabilized cells or skin samples were blocked with Duolink® blocking solution for 60 min at 37 °C in a humidified chamber. Samples were then incubated with primary antibodies raised in different species, followed by washing and incubation with species-specific PLA probes (PLUS and MINUS) for 1h at 37 °C. After washing, ligation was performed for 30 min at 37 °C in the presence of ligase, followed by rolling circle amplification for 100 min at 37 °C with polymerase. All washes were performed at room temperature with the supplied wash buffers. Samples were mounted using Fluoromount-G mounting medium with Hoechst and imaged using the Zeiss LSM980 confocal microscope with Airyscan 2 detector. Negative controls omitting primary antibodies were included to assess signal specificity.

### Nuclear import and export inhibition assay

To investigate the mechanism of IgG nuclear transport, cells were treated with pharmacological inhibitors targeting distinct steps of nucleocytoplasmic trafficking. Cells were incubated for 2 hours at 37 °C with importazole (20 µM, inhibitor of importin-β–mediated nuclear import, MilliporeSigma, Burlington, Massachusetts, USA), ivermectin (10 µM, inhibitor of importin-α and β–mediated nuclear import, MilliporeSigma, Burlington, Massachusetts, USA) or leptomycin B (20 nM, inhibitor of CRM1/exportin-1–mediated nuclear export, MilliporeSigma, Burlington, Massachusetts, USA). After the first hour of treatment, cells were incubated with ATAs for one more hour, under standard incubation conditions and processed according to the established immunofluorescence protocol. Parallel samples treated with DMSO served as vehicle controls.

### SiRNA

SiRNA-mediated knockdown of caveolin 1 (CAV1), clathrin light chain B (CTLB) and FcRN were performed using ON-TARGETplus SMARTpool siRNAs (Dharmacon, Horizon Discovery, Lafayette, CO, USA) targeting human CAV1, CTLB, and FCGRT respectively. siRNAs were diluted in Dharmacon siRNA buffer to a final concentration of 25 nM per target and transfected using DharmaFECT reagent (Dharmacon, Horizon Discovery, Lafayette, CO, USA) according to the manufacturer’s instructions. Following transfection, fibroblasts and endothelial cells were incubated in the serum and antibiotics-free medium containing siRNA for 20h. Cells were then washed and incubated with ATAs for 1h and processed according to our standard immunofluorescence protocol. A transfection reagent-only condition was included as a negative control in all experiments. Following siRNA transfection, knockdown efficiency for each target (clathrin, caveolin, and FcRn) was confirmed by Western blotting and RT-qPCR.

### Western blot

To examine phosphorylation in STING, endothelial cells were cultured with ATAs (10 µg/ml) or cGAMP (50 µg/ml). Cells were lysed in RIPA buffer containing PhosSTOP™ (Sigma Aldrich), complete protease inhibitor (Sigma Aldrich) and 0.05 % Benzonase nuclease (Sigma Aldrich). A BCA test was done, and lysate concentrations were normalized with RIPA buffer. 4x Laemmli sample buffer (Bio-Rad) and XT reducing agent (Bio-Rad) were added, and lysates were boiled for two minutes at 95°C. Then, the lysates were run at 70 V on an 8-16% Criterion™ TGX™ precast midi protein gel (Bio-Rad) in 1x NuPage SDS running buffer (Thermo Scientific). Precision Plus Protein™ Dual Xtra (Bio-Rad) was used as a standard. Proteins were blotted using a Trans-Blot Turbo Midi 0,2 µm Transfer pack (Bio-Rad) in a Trans-Blot® Turbo™ transfer system (Bio-Rad). The membrane was blocked for 1 h with 5% (w/v) BSA in 1x TBST. Rabbit anti-human STING (Cell Signaling Technology, Cat. 13647) and rabbit anti-human pSTING monoclonal antibodies (Cell Signaling Technology, Cat. 197815) were diluted 1:1,000 in 5% (w/v) BSA in 1x TBST. The mouse anti-human Vinculin monoclonal antibody (Sigma Aldrich, Cat. V9131) was diluted 1:10,000. Primary antibody incubations were done overnight. The membrane waswashed three times with 1x TBST. The secondary antibodies, peroxidase-conjugated donkey anti-rabbit IgG (Jackson Immunoresearch, 711-036-152) and peroxidase-conjugated donkey anti-mouse IgG (Jackson Immunoresearch, 711-036-150) were diluted 1:10,000 in 1x TBST and added for 1 h. The membrane was washed again three times and imaged on a Azure300 imager using the SuperSignal West Dura Extended Substrate (Thermo Scientific) and the SuperSignal West Femto Maximum Sensitivity Substrate (Thermo Scientific).

To examine ATAs function as a primary antibody, 2x10^6^ CaCo2 cells lysed in buffer containing 0.1% NP-40, 10 mM Tris pH 7.9, 10 mM MgCl_2_, 15 mM NaCl, 2 µM DTT and 2 mM PMSF followed by 10 min incubation on ice. Pellet formed by centrifugation at 2200 RPM at 4°C for 5 min. The supernatant was discarded, and the pellet was incubated rotating (4 RPM) with buffer containing 0.5 M NaCl, 20 mM HEPES pH 7.9, and 20% glycerol at 4°C for 1 hour. Cells were centrifugated at 11,000 RPM at 4°C for 10 min. 2.5-20 µL supernatant mixed with SDS loading buffer (2% SDS, 2 mM β-mercapto-ethanol, 4% (v/v) glycerol, 50 mM Tris–HCl pH 7, 0.05% bromophenol blue) and loaded on a 10% SDS gel. As a positive control, 20 and 40 ng purified TOP1 were loaded on the SDS gel. Size marker was PageRuler prestained Protein ladder (Thermo Scientific). The gel was running at 200 V in 1x SDS running buffer (25 mM Tris–HCl, 192 mM glycine, 0.1% SDS) for 1 hour. The proteins were blotted onto a 0.45 Nitrocellulose Blotting Membrane (Amersham™ Protran™) at 100V for 1h and 10 min in transfer buffer (10 mM CAPS, 10% EtOH). The nitrocellulose membrane was incubated in blocking buffer (10 mM Tris pH 7.5, 500 mM NaCl, 0.25% Tween 20, and 5% skimmed milk powder) overnight at 4°C. Subsequently, the membrane was incubated for 1 h at room temperature with 10 ng/mL purified ATA diluted in blocking buffer. The membrane was washed for 3 × 30 minutes in TBS buffer (20 mM Tris pH 7.5, 0.5 M NaCl, 0.25% Tween 20) at room temperature and probed with horseradish peroxidase-conjugated donkey anti-human IgG (Jackson Immunoresearch, 709-035-149) at room temperature for 1 h. The membrane was washed 3x15 min in washing buffer at room temperature, and bound antibodies were detected using the ECL Prime Western Blotting Detection Reagents (Cytiva). The membrane was exposed for appropriate time periods in order to obtain a clear visualization of the blotted bands using Amersham Imager 600.

For silver staining, 20 ng/µl ATAs and HC-IgG were mixed with SDS loading buffer (2% SDS, 4% (v/v) glycerol, 50 mM Tris–HCl pH 7, 0.05% bromophenol blue) with or without 2 mM β-mercaptoethanol. The IgG was desaturated by 5 min at 95°C before loading on a 10% SDS gel. Size marker was PageRuler prestained Protein ladder (Thermo Scientific). The gel was running at 200V in SDS running buffer (25 mM Tris–HCl, 192 mM glycine, 0.1% SDS) for 42 min. Silver staining was performed according to manufacturer protocol (Invitrogen A/S).

To analyze the presence of TOP1 after ATA treatment, 25 µL nuclear extractions from endothelial cells were mixed with 6.25 µL 4x SDS loading buffer (250mM Tris-HCl pH 6.8, 8% SDS, 40% glycerol, 0.03% bromophenol blue, 20% β-Mercaptoethanol) and heated at 95°C for 5 min. The entire sample was loaded on a 10% SDS gel (Invitrogen) together with 10 µL protein marker (Thermo Fisher, 26619). The gel was running at 200V for 55 min in 1x SDS running buffer (25mM Tris, 192mM glycine, 0.1% SDS). The gel was transferred to a 0.45 µM nitrocellulose membrane (GE Healthcare Life Sciences) in a “sandwich” with one sponge and three Whatman papers on each side of the membrane and gel. The sandwich is placed in a blotting apparatus in 1x CAPS (10mM CAPS, 10% v/v 96% EtOH) at 4°C at 100V for 80 min. The membrane was cut in two around 70 kDa, and both membranes were placed in blocking buffer (TBST, 5% w/v skimmed milk powder) at RT for 1h. The blocking buffer for the upper part of the membrane was replaced with blocking buffer containing 2000x diluted rabbit anti-TOP1 (Betnyl, A302-590A) antibody. The blocking buffer for the lower part of the membrane was replaced with blocking buffer containing 1000x diluted rabbit anti-Histone H3 (Abcam, ab1791) antibody. The membranes were incubated with primary antibody rotating O/N at 4°C. The membranes were washed three times with TBST for 30 min. Blocking buffer containing 2000x diluted goat anti-rabbit (Dako, P0448) was added to each membrane and incubated rotating at RT for 1h. The membranes were washed three times in TBST for 15 min, followed by detection with ECL reagents (Cytiva) diluted 1:1. Images were taken with Amersham Imager 600.

### Functional type I IFN measurements

Bioactive functional type I IFN quantified in supernatants using the reporter cell line HEK-Blue IFN-α/β (Invivogen) according to manufacturer’s instructions. The cell line was maintained in DMEM supplemented with GlutaMax-I (Gibco, Life Technologies), 10% heat-inactivated FCS, 100 µg/mL streptomycin and 200 U/mL penicillin, 100 µg/mL normocin (InvivoGen), 30 µg/mL blasticidin (InvivoGen) and 100 µg/mL zeocin (InvivoGen). For measurement of functional type I IFN, cells were seeded at 3 × 10^4^ cells/well in 96-well plates in 150 µL medium devoid of Blasticidin and Zeocin and treated either HC-IgG and ATAs (10 µg/mL); cGAMP (16 µg/mL). The following day, 50 µl of the supernatant for analysis was added to the HEK-Blue cells. SEAP activity was assessed by measuring optical density (OD) at 620 nm on a microplate reader (ELx808, BioTEK). The concentration was determined from a standard curve made with IFN-α (IFNa2 PBL Assay Science) ranging from 2 to 500 U/mL.

### Enzyme-linked Immunosorbent assay (ELISA)

CXCL-10 (Invitrogen) was measured according to the manufacturer’s instructions. IFN-α 2a in the cell culture supernatants were determined using the U-PLEX Interferon kit (MSD) according to the manufacturer’s instructions.

### Detection of TOP1 activity by REEAD assay

The REEAD assay was performed essentially as previously described (*14*, *32*). TOP1 reactions were carried out in a 20 μL reaction volume containing divalent cation depletion buffer (10 mM Tris-HCl, pH 7.5, 5 mM EDTA, and 50 mM NaCl). Reaction mixtures were supplemented with 500 nM (TOP1 DNA substrate. Reactions were initiated by the addition of 8 μL inuclear extract (obtained from the same cell samples used for quantification of TOP1 protein level by Bratford) or purified TOP1 (purified as previously reported (*33*) and used for all experiments as positive control). For experiments with purified ATAs, reactions were initiated with 8 fmol/μl purified TOP1 mixed with 5-20 ng/μL ATAs from four patients or 20 ng/μL ATA-depleted IgG. Incubation was continued for 1h at 37°C. Circularized closed circles were then hybridized to a 5’amino primer-coated slide followed by RCA with incorporation of biotinylated dCTP. Subsequently, the slide was blocked in 1× TBST (20 mM Tris-HCl pH 9, 150 mM NaCl, 0.05% Tween20 pH 9) supplemented with 5% skimmed milk powder and 5% BSA for 1 h at room temperature, followed by incubation with 1:300 HRP-conjugated anti-biotin antibody in 1× TBST supplemented with 5% skimmed milk powder and 5% BSA, for 1 h at room temperature. The slide was washed 3 × 3 min in 1× TBST before the addition of 2 µL of 1:1 ECL mixture to allow chemiluminescence readout using a CCD camera. For all cell lines, TOP1 activity was normalized to the activity of purified TOP1 circles and protein concentration in nuclear extracts as determined with Bradford’s protocol. Results were plotted as the average of three independent experiments.

### Bradford

Protein concentration in nuclear extractions from endothelial cells was analyzed with the Bradford assay. BSA was diluted in ddH_2_O to make a serial dilution (0-1 mg/mL). 5 µL of each dilution of BSA was loaded in a Maxisorp 96-well plate (Sarstedt, 82.1583) and 10 µL of nuclear extraction. To each well, 200 µL Bradford reagent (Sigma Aldrich, B6916) was loaded and incubated for 10 min at RT. The absorbance was measured with Multiplate Go (Thermo Scientific) at 595 nm. The concentration of each BSA dilution was calculated with linear interpolation and used for the calculation of the protein concentration in nuclear extractions.

### Relaxation Assay

TOP1 activity was assayed using a DNA relaxation assay by incubating 125 fmol/μL of TOP1 with 10 fmol/μL of negatively supercoiled plasmid pUC18 in 20 μl of reaction buffer (10 mM Tris-HCl, 5 mM EDTA, 150 mM NaCl, pH 7.5) in the presence or absence of 5-10 ng/µL ATAs. The effect of ATAs on the relaxation activity of TOP1 was assayed by incubating ATAs and TOP1 for 5 min at 37°C before the addition of the plasmid. The reactions were carried out at 37°C for 10 min and stopped by the addition of 0.1% SDS. The samples were digested with 1 µg/µL Proteinase K at 37°C for 1h and electrophoresed in a horizontal 1% agarose gel in 1xTAE (40 mM Tris-acetate and 1 mM EDTA) at 20Vfor 16 h. The agarose gel was stained with 0.5 ug/mL EtBr for 30 min and destained, and the bands were visualized using Gel Doc XR+ (Bio-Rad).

### Nicking assay

TOP1 activity was assayed using a DNA nicking assay by incubating 270 fmol/μL of TOP1 with 10 fmol/μL of negatively supercoiled plasmid pUC18 in 20 μl of reaction buffer (10 mM Tris-HCl, 5 mM EDTA, 150 mM NaCl, pH 7.5) in the presence or absence of 40 ng/μl of ATA. Camptothecin (CPT) was used as a positive control as stabilized TOP1-bound DNA nicks (TOP1cc) generated by CPT are well known to induce DNA fragmentation and STING activation in human cells (*34*). The effect of ATAs in stabilizing the TOP1cc was assayed by incubating ATAs and TOP1 for 5 min at 37°C before the addition of the plasmid. The reactions were carried out at 37°C for 30 min and stopped by the addition of 0.1% SDS. The samples were digested with 1 µg/µL Proteinase K at 37°C for 1h and electrophoresed in a horizontal 1% agarose gel containing 0.5 µg/mL EtBr in 1xTAE (40 mM Tris-acetate and 1 mM EDTA) at 65V during 2 h. The bands were visualized using Gel Doc XR+ (Bio-Rad).

### Immunofluorescence

#### Antibodies and reagents for immunofluorescence

The antibodies and reagents utilized for immunofluorescence are listed in this table:

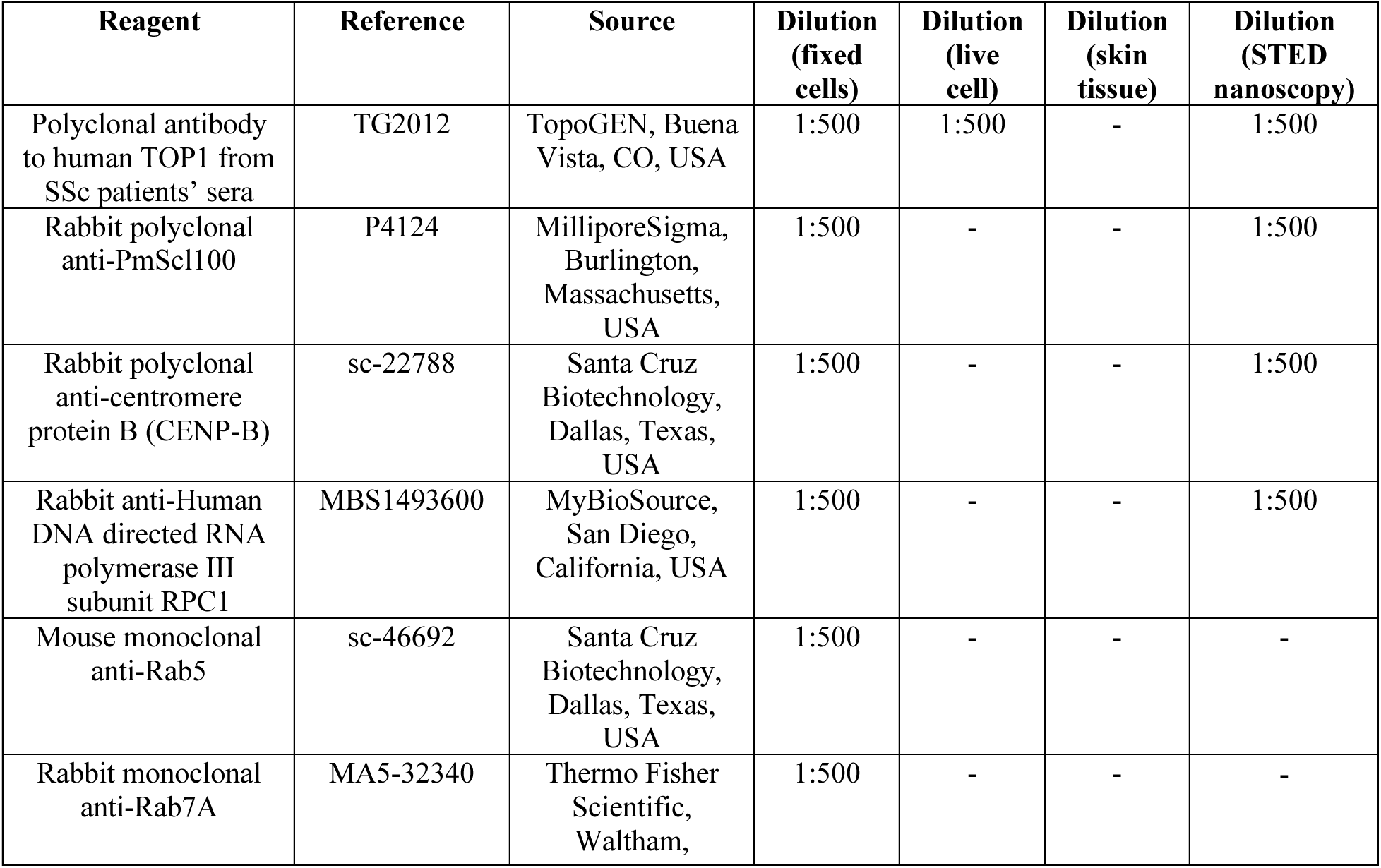
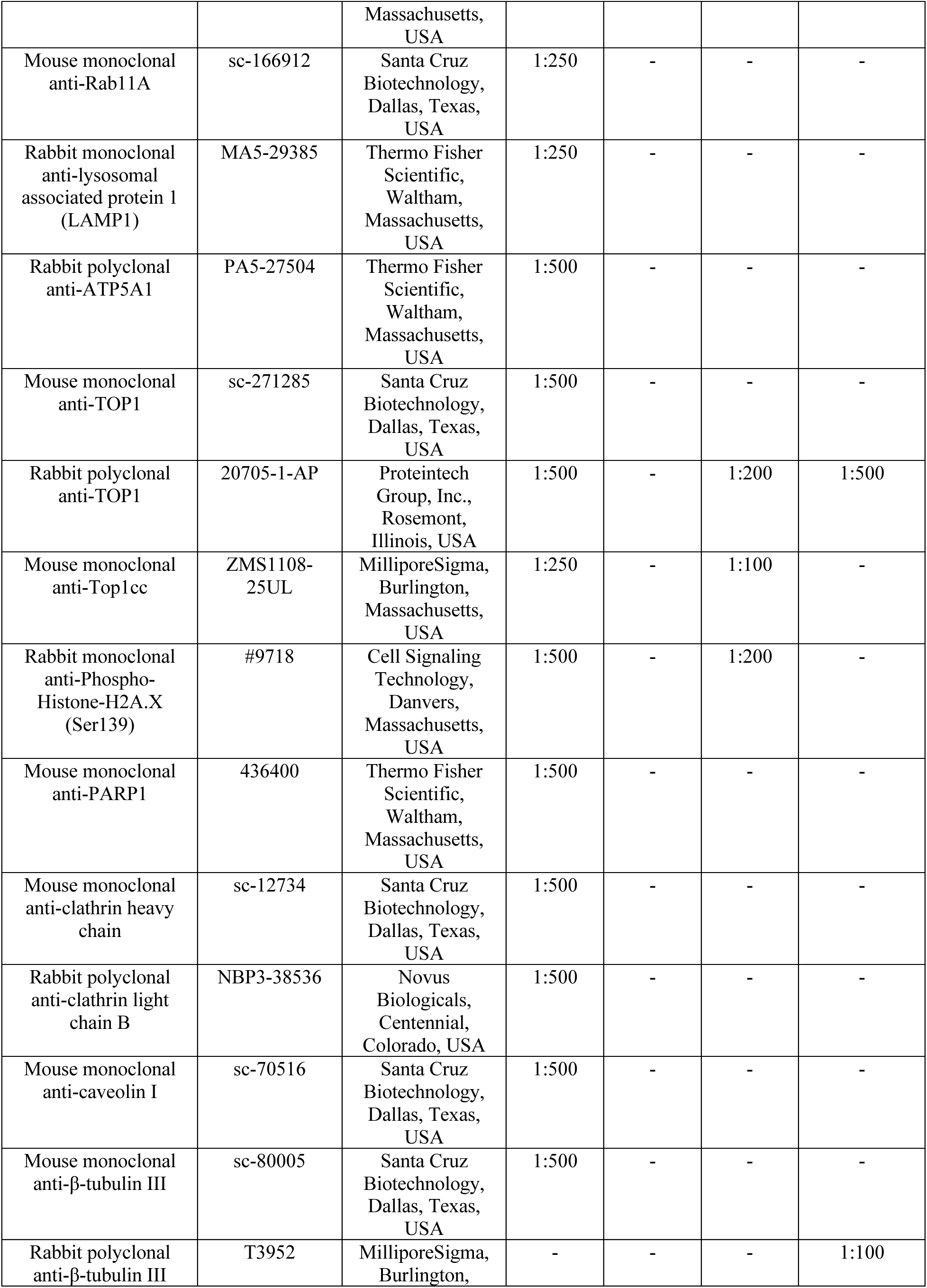
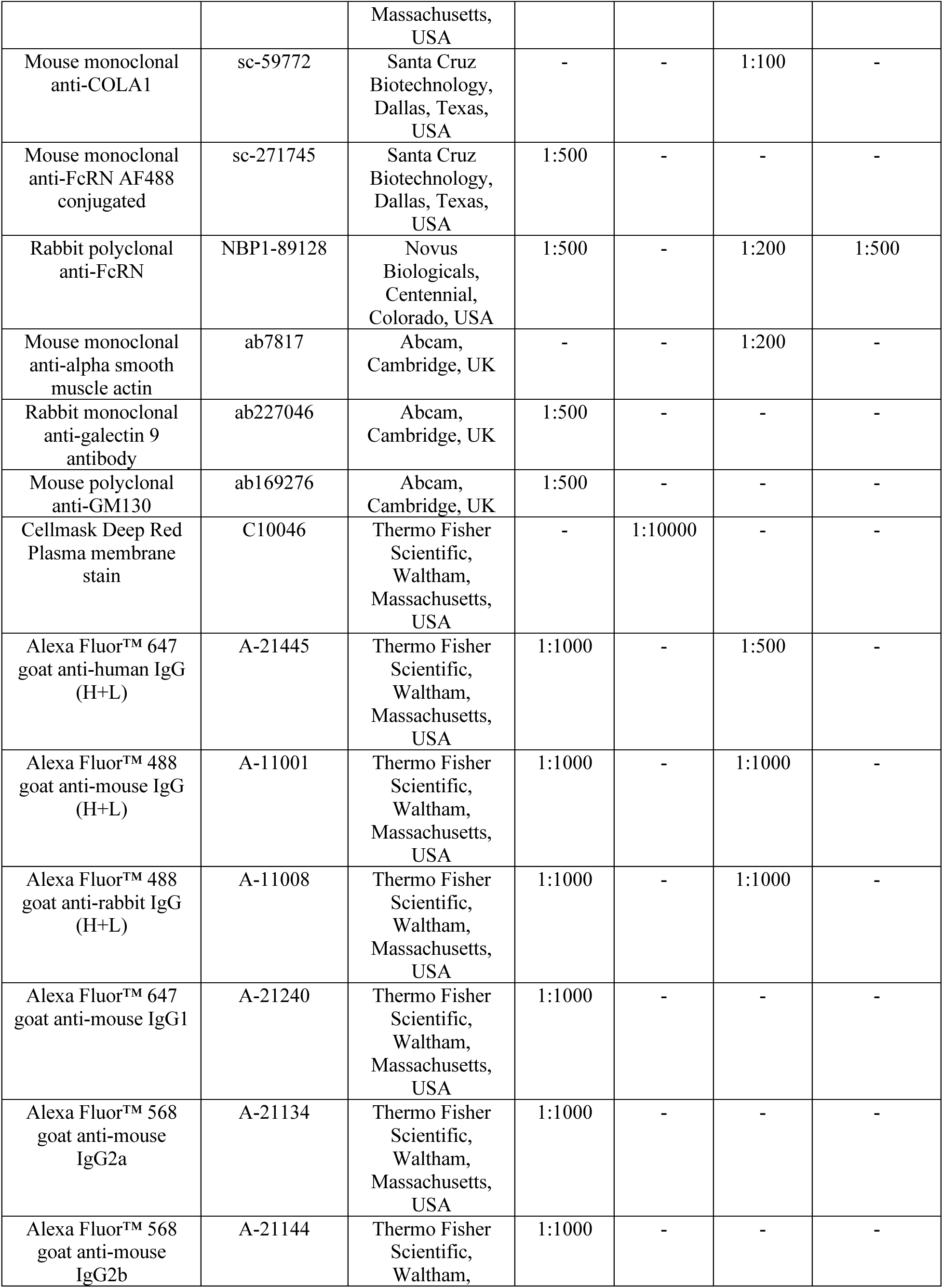
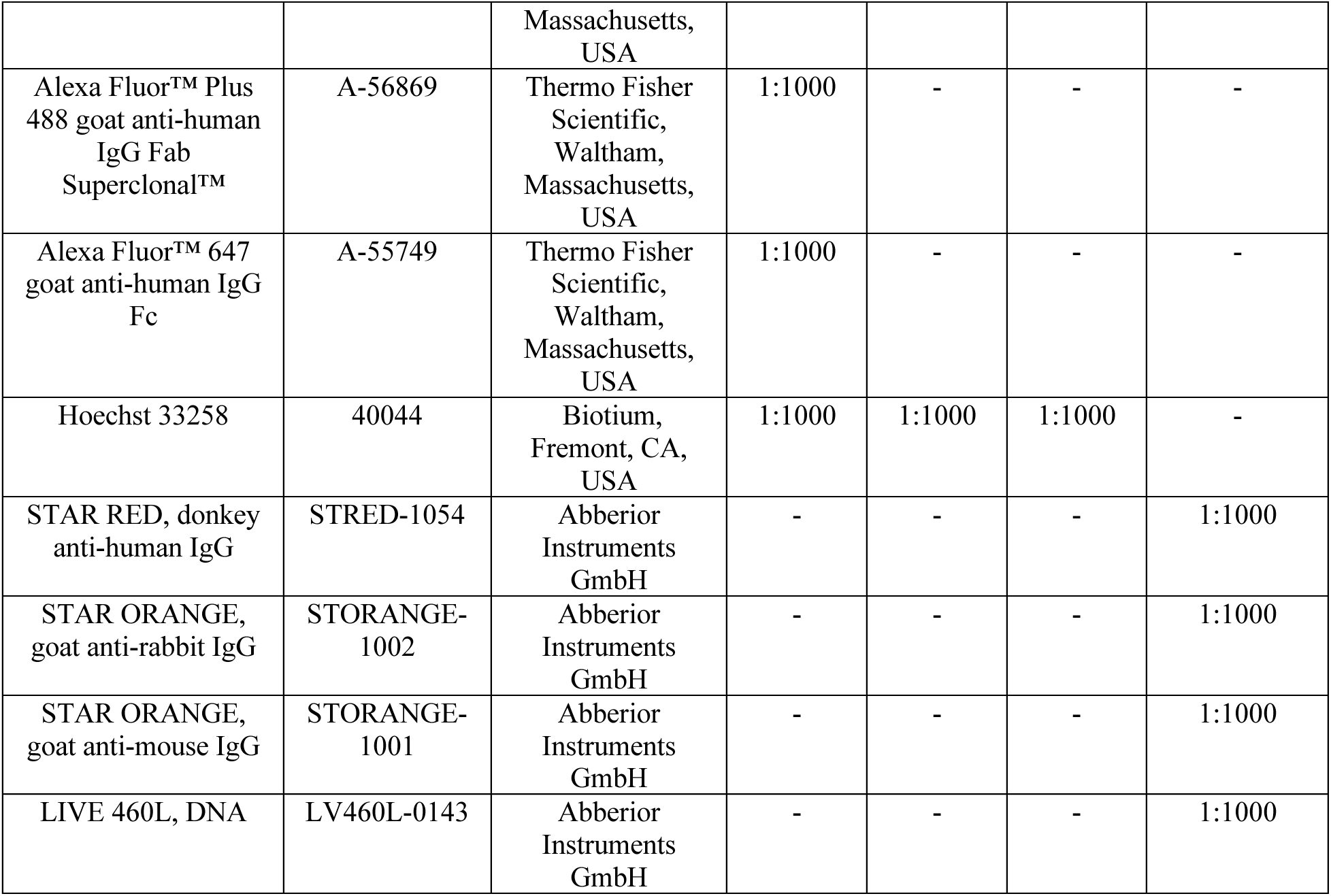

#### Fixed immunofluorescence

Fibroblasts and endothelial cells were plated on a coverslip in 6-well-plates at a density of 30,000 cells per well and cultured for 72h. To avoid interference due to the presence of FCS, cells were FCS-deprived for 24h before antibody incubation in all experiments, unless specified otherwise. After 24h deprivation, cells were incubated with ATAs or purified IgG diluted in fresh culture medium for durations ranging from 15 minutes to 72h, as mentioned in the corresponding results. After the incubation period, cells were washed, fixed in 4% buffered formaldehyde (MM France, Brignais, France) and permeabilized with PBS-triton 0.1%. Cells were then saturated with PBS/bovine serum albumin (BSA) 1% for 30 minutes. Primary and secondary antibodies were diluted in a PBS/BSA 1% solution. Primary antibodies were incubated overnight after two washing steps with PBS/triton 0.05%. Secondary antibodies and Hoechst (Biotium, Fremont, CA, USA) were incubated for 1h. After the secondary antibody incubation step, the slats were mounted on a microscope slide using Fluoromount-G mounting medium (Thermo Fisher Scientific, Waltham, Massachusetts, USA). Microscope slides were read using a Zeiss LSM980 confocal microscope with Airyscan 2 detector, and images were analyzed with IMARIS software (version 10.2, Oxford Instruments).

#### Ex-vivo skin biopsies immunofluorescence

Forearm skin biopsies of SSc patients and HC were collected and embedded in OCT compound, frozen in liquid nitrogen and stored at -80°C. Blocks were sectioned (8 µm thickness) on a cryostat onto slides, fixed in cold acetone for 10 minutes and stored at -80°C. For staining, samples were thawed, rehydrated in PBS, washed in PBS-Tween 0.05%, and blocked for 60 min in 1% BSA, 5% goat serum, 0.1% Tween-PBS solution. Samples were incubated overnight at 4°C with primary antibodies diluted in blocking buffer. Sections were then washed in PBS-Tween 0.05% (3 × 5 min) and incubated for 1h at room temperature with fluorophore-conjugated secondary antibodies. Samples were then washed and mounted in Fluoromount-G mounting medium (Thermo Fisher Scientific, Waltham, Massachusetts, USA). Images were acquired using a Zeiss AxioObserver Z1-Yokogawa CSU-X1 spinning disk confocal microscope for quantitative analyses and illustrations were obtained with a Zeiss LSM980 confocal microscope with Airyscan 2 detector.

#### Skin explants immunofluorescence

After 1 day, 3 days or 7 days post antibody injection, skin explants were extracted, embedded in OCT snap frozen in ice-cold isopentane and stored at -80°C. Blocks were sectioned (20 µm thickness) on a cryostat onto slides, fixed in acetone for 15 minutes and stored at -80°C. For staining, samples were thawed, rehydrated in PBS, washed in PBS-Triton 0.1%, permeabilized in PBS-Triton 0.2% for 20 minutes and blocked for 60 min in 1% BSA, 5% goat serum, 0.1% Triton-PBS solution. Samples were incubated overnight at 4°C with primary antibodies diluted in blocking buffer. Sections were then washed in PBS-Triton 0.1% (3 × 5 min) and incubated for 1h at room temperature with fluorophore-conjugated secondary antibodies. Samples were then washed and mounted in Fluoromount-G mounting medium (Thermo Fisher Scientific, Waltham, Massachusetts, USA). Images were acquired using a Zeiss AxioObserver Z1-Yokogawa CSU-X1 spinning disk confocal microscope or Zeiss Axioscan Z1 slide scanner for quantitative analyses and illustrations acquired using a Zeiss LSM980 confocal microscope with Airyscan 2 detector.

#### Incucyte

HUVEC’s cultured in supplemented ECBM2 medium (PromoCell, C-22210) were seeded 5000 cells per well in a gelatine-coated 96-well plate three days before stimulation. HC-IgG and purified ATAs from three donors each were conjugated with FITC, and excess dye was removed on Zeba Spin Desalting Columns (7 kDa MWCO). The conjugated antibody mixes were added to HUVEC’s for 24 hours at a concentration of 10µg/mL. Wells were washed twice and the plate was incubated in the Incucyte® S3 Live-Cell Analysis Instrument for three days with three images being taken per well every hour. The total area of green fluorescence for each condition over time was obtained using the Incucyte software®.

#### Live cell imaging

##### Classical live cell microscopy

FB were plated on clear flat bottom black 24-well-plates (Ibidi GmbH, Gräfelfing, Germany) at a density of 5,000 cells per well, after 30 minutes of gelatin 0.1% coating, and cultured for 72h. To avoid interference due to the presence of FCS, cells were FCS-deprived for 24h before antibody incubation. After 24h deprivation, cells were stained with Hoechst and CellMask™ Deep Red plasma membrane stain (Thermo Fisher Scientific, Waltham, Massachusetts, USA) for 1h, before washing. Cells were then incubated with 25 µg/mL fluo-ATAs and imaged immediately using Zeiss LSM980 confocal microscope. Time series were analyzed using IMARIS software (version 10.2, Oxford Instruments).

##### 3D-high resolution live microscopy

FB were seeded at a density of 5,000 cells per well in an 8-well polymer-bottom chamber slide (Ibidi GmbH, Gräfelfing, Germany) coated with 0.1% gelatin and cultured for 72h prior to imaging. One hour before acquisition, the culture medium was supplemented with Hoechst nuclear stain. Immediately before imaging, 25 µg/mL of fluo-ATAs were added to culture medium, and image acquisition was initiated without delay. Live-cell imaging was performed using a Zeiss LSM 980 confocal microscope equipped with an Airyscan 2 detector under physiological conditions. Z-stacks spanning the full thickness of the nuclei were acquired using the Airyscan 2 Multiplex 4Y mode to maximize imaging speed and temporal resolution. Time-lapse sequences were recorded continuously, for a total duration of 15 to 30 minutes, immediately following fluo-ATAs addition. Three-dimensional image reconstruction, nuclear segmentation, antibody spot segmentation, and single particle tracking analyses were performed using Imaris software (version 10.2, Oxford Instruments).

#### Tracking

Time-series Z-stack images were imported into Imaris (version 10.2, Oxford Instruments). Nuclear surfaces were reconstructed from the Hoechst channel and used to define nuclear boundaries in three dimensions. For trajectory visualization, ATAs particles were detected using the Spots module, as described above, with estimated diameters ranging from 0.15 to 0.8 µm and tracked using the Imaris autoregressive motion algorithm (maximum frame-to-frame linking distance 5 µm). Particles were subsequently classified according to their position relative to the reconstructed nuclear surface and visualized as distinct color-coded populations inside or outside the nucleus. Trajectories are shown for illustrative purposes only and were not subjected to quantitative analysis.

#### Super-resolution microscopy by Stimulated emission depletion (STED)

Cells were fixed, permeabilized, and immunostained following the standard protocol described above. Cells were then counterstained with Abberior fluorophore-conjugated secondary antibodies and mounted in the Abberior STED-compatible mounting medium. Imaging was performed on an Abberior STEDYCON system, using the appropriate depletion lasers and detection settings for super-resolution acquisition. Post-acquisition image processing was performed using ImageJ software (version 1.53c, National Institutes of Health, USA). Deconvolution of STED images was performed using Huygens Essential software (version 25.10, Scientific Volume Imaging, Hilversum, The Netherlands).

#### Immunofluorescence analysis

Immunofluorescence images were acquired by Airyscan confocal microscopy unless specified otherwise. Quantitative image analysis and colocalization measurements were performed on single optical sections acquired at the plane of maximal nuclear signal. Selected samples were additionally imaged as z-series with a step size of 0.15 µm to illustrate spatial organization, which was consistent with observations from single optical sections. Airyscan image analysis was performed using Imaris software (version 10.2, Oxford Instruments). For all experiments, a secondary antibody-only control (without primary antibody or human IgG in culture medium) was included for each batch and treatment group to assess nonspecific background signal and accurately determine true signal intensity.

#### IgG spot detection

IgG spot detection was performed using the Imaris Spots module based on local intensity maxima, with an estimated spot diameter of 0.3 µm. Automatic background subtraction was applied, and spots were filtered using a quality threshold that was kept constant across all images from the same experiment.

#### Surface & compartments

Surface reconstruction (e.g. nuclei from Hoescht channel) and cell compartment segmentation were performed using Imaris Surfaces module with background subtraction enabled and a fixed intensity threshold applied to all datasets from the same experiment.

#### Colocalization analyses

Intensity-based colocalization between IgG and target nuclear antigens (e.g. TOP1) were assessed using M_1_ and M_2_ Manders coefficient in Imaris (version 10.2, Oxford Instruments). Analysis was performed on raw Airyscan images without additional filtering. Regions of interest corresponding to individual nuclei or cells were defined based on surface masks generated in Imaris. Manual intensity thresholds were applied independently to each channel to exclude background signal. Threshold values were determined using negative control samples lacking specific staining and were kept constant across all experimental conditions.

For object-based colocalization, spots representing IgG puncta were segmented with the Spots tool, and the relevant compartment (e.g. Rab5) was segmented with the Surfaces tool. For each image, the fraction of POI spots located inside the compartment surface was determined and normalized to the fraction expected from 1000 complete spatial randomness (CSR) simulations performed in Imaris Vantage Spatial View, yielding an enrichment score (Observed ÷ Expected). Enrichment scores >1 indicate preferential localization of IgG spots to the compartment. One fraction of colocalized spots with POI and one enrichment score were calculated per image and used as the unit of replication.

#### Picrosirius red staining

Frozen skin samples were sectioned at 8 µm thickness and fixed in acetone for 15 minutes. Samples were thawed, rehydrated in PBS and through graded ethanol solutions, sections were stained with 0.1% Direct Red 80 (MilliporeSigma, Burlington, Massachusetts, USA) dissolved in saturated aqueous picric acid (Picrosirius Red solution) for 30 minutes at room temperature. Slides were subsequently washed 2 x 2 minutes in acidified water (0.5% acetic acid), dehydrated through graded ethanol, cleared in xylene, and mounted with a resinous medium (*35*). Brightfield and polarized light images were acquired using a Zeiss Axiophot 2 microscope under identical exposure settings. Collagen-positive areas were quantified using ImageJ software (version 1.53c, National Institutes of Health, USA) on randomly selected fields and normalized to total tissue area.

### Statistical analysis

For histology data, normality assumptions were assessed using the Shapiro-Wilk test and visual inspection of Q-Q plots. Where normality assumptions were met, group comparisons were performed using one-way ANOVA with Tukey’s post hoc correction for multiple comparisons, or Welch’s t-test for pairwise comparisons.

For cellular microscopy data and all other experiments, data did not meet normality assumptions; group comparisons were therefore performed using the Kruskal-Wallis test with Dunn’s post hoc correction for multiple comparisons, or the Mann-Whitney U test for pairwise comparisons. Statistical significance is reported as follows: ns, non-significant; *P < 0.05; **P < 0.01; ***P < 0.001; ****P < 0.0001. All statistical analyses were performed using GraphPad Prism (version 11.0.0, GraphPad Software, San Diego, CA, USA).

### Oligonucleotide substrates, primers, and probes

Oligonucleotides for construction of the S (hTopI) substrate, TDP1 substrate, and amino primer (p) (DNA Technology A/S, Denmark) and synthesized using the 394 DNA synthesizer (Applied Biosystems). Oligonucleotide sequences were as follows: S (Top1): 5’AGAAAAATTTTTAAAAAAACTGTGAAGATCGCTTATTTTTTTAAAAATTTTTCTAAGTCTTTTAGATC-CCTCAATGCACATGTTTGGCTCC-GATCTAAAAGACTT3’ RCA primer: 5′ Am-CCAACCAACCAACCAAATAAGCGATCTTCACAGT3’ TDP1 sensor: 6FAM-AAA GCA GGC TTC AAC GCA ACT GTG AAG ATC GCT TGG GTG CGT TGA AGC CTG CTT T-BHQ1, where 6FAM was attached to the DNA through a phosphothioate and to 3′BHQ1 through a phosphodiester linkage.

RNA was collected and purified using RNeasy Mini Kit (Qiagen Cat no 74104) and converted to cDNA using the iScript cDNA synthesis kit (Bio-Rad) following the manufacturer’s instructions. RNA quality and quantity were measured on a Nanodrop (Thermo Fisher Scientific). RNA was reverse transcribed using SuperScript II Reverse Transcriptase (Invitrogen) into cDNA that was then used for qPCR with SYBR green PCR kit (KK4611). CT values were normalized to ACTB (ΔCT). SYBR green prime probes used include ACTB (fw: 5’-GCA CTC TTC CAG CCT TCC TT-3’ rev: 5’-CTC CTT CTG CAT CCT GTC GG-3’), CXCL-10 (fw: 5’-GAA CCT CCA GTC TCA GCA CC-3’ rev: 5’-GCA GGT ACA GCG TAC AGT TCT-3’), IL-6 (fw: 5’-TGT GAA AGC AGC AAA GAG GC-3’ rev: 5’-ACC TCA AAC TCC AAA AGA CCA GT-3’), IFN-b1 (fw: 5’-TGC TCT CCT GTT GTG CTT CT-3’ rev: 5’-AGC CTC CCA TTC AAT TGC CA-3’).

### Bulk RNA sequencing

Endothelial cells were cultured as previously described. After 4 h of pre-stimulation with IFNα, the medium was replaced and cells were incubated for 6 h with HC-IgG or ATAs, both at 10 mg/mL, with three biological replicates per condition. Cells were then lysed.

RNA was purified using the miRNeasy Tissue/Cell Kit, and genomic DNA was removed using the RNase-Free DNase Set (QIAGEN, Cat. No. 79254). RNA integrity and concentration were assessed using a Bioanalyzer RNA Nano chip. Libraries were prepared from 200 ng purified RNA using the SMARTer Stranded Total RNA Sample Prep Kit - HI Mammalian (Takara, Japan), according to the manufacturer’s instructions. Library fragment size was assessed using the Bioanalyzer High Sensitivity DNA Analysis Kit (Agilent), and library concentration was measured using the Qubit dsDNA High Sensitivity Kit (Invitrogen). Libraries were pooled in equimolar amounts and sequenced on a NovaSeq X Plus using a 10B flow cell with paired-end 150-cycle sequencing (Illumina).

### Proteomics sample preparation, LC-MS/MS acquisition, and preprocessing

Cytoplasmic and nuclear proteins were extracted and analyzed separately by LC-MS/MS. Protein digestion using the FASP protocol, LC-MS/MS acquisition, and raw data processing were performed as previously described (*36*). For each sample, 1 µg digested peptide was analyzed using a Proxeon Easy-nLC-1000 system (Thermo Scientific) coupled to a Q-Exactive Orbitrap mass spectrometer (Thermo Fisher Scientific).

### Bioinformatics

#### RNA-seq data processing and differential expression analysis

Raw transcriptomic count matrices were analyzed in R using DESeq2 (v1.50.2). Samples were assigned to three conditions: HC-IgG, ATAs and NS, with three biological replicates per condition. A DESeqDataSet object was constructed using the design formula *∼ condition*.

Genes with low expression were removed before model fitting. Genes were retained if they had counts greater than 10 in at least three samples. Differential expressions were modeled using negative binomial generalized linear models. Pairwise contrasts were extracted for ATAs versus HC-IgG, ATAs versus NS, and NS versus HC-IgG. ATA^+^ IgG versus healthy control IgG and ATA^+^ IgG versus IFNα were retained for downstream enrichment analyses.

For each contrast, result tables included base mean expression, log2 fold change, standard error, Wald statistic, nominal P value, and Benjamini–Hochberg-adjusted P value. Log2 fold changes were additionally shrunk using lfcShrink with the ashr method for visualization. Ensembl gene identifiers were mapped to human gene symbols using AnnotationDbi (v1.72.0) and org.Hs.eg.db (v3.22.0); genes without mapped symbols or adjusted P values were removed from gene-symbol-based analyses. Genes were considered differentially expressed when |log₂ fold change| > 1 and FDR < 0.05.

Variance-stabilizing transformation was performed using vst (blind = TRUE) for visualization and quality control. Sample-to-sample distances, hierarchical clustering, PCA, volcano plots, and heatmaps were generated from transformed or processed expression matrices as appropriate.

#### Proteomic data processing and differential abundance analysis

Raw LC-MS/MS data were processed using MaxQuant with the MaxLFQ algorithm, version 1.5.2.8, and the Andromeda search engine. Spectra were searched against the UniProtKB/Swiss-Prot human protein database, release 2021-03, restricted to Homo sapiens entries. Normalization was performed in Perseus after retaining proteins with at least two valid LFQ intensity values in each condition: HC-IgG, ATA^+^-IgG, ACA^+^-IgG, ACA^-^/ATA^-^-IgG. LFQ intensities were log2-transformed, missing values were imputed from a normal distribution, and proteins were mapped to unique gene symbols. The resulting cytoplasmic and nuclear matrices were exported as ‘cytoplasm_matrix.txt’ and ‘nucleus_matrix.txt’.

Cytoplasmic and nuclear proteomic matrices were analyzed independently in R. One cytoplasmic NS sample and one nuclear ATA^+^ sample were excluded before modeling based on exploratory quality-control assessment. PCA and PLS-DA were performed using mixOmics (v6.34.0) to assess sample-level structure; these analyses were used for visualization and quality control only.

Differential protein abundance was assessed using limma (v3.66.0). For each fraction, a no-intercept linear model was fitted with condition as the model factor. Contrasts compared ATA^+^-IgG, ACA^+^-IgG, ACA^-^/ATA^-^-IgG, and ns control samples against HC-IgG. Additional pairwise contrasts were computed for exploratory downstream analyses. Models were fitted using lmFit, contrasts were applied with contrasts.fit, and moderated statistics were obtained using empirical Bayes moderation with eBayes.

Proteomic log2 fold changes were further stabilized using adaptive shrinkage with ashr. Standard errors were estimated from the ratio between log2 fold change and moderated t statistic for proteins with finite, non-zero statistics. Posterior mean shrunken log₂ fold changes and local false sign rates were retained for visualization. Proteins were considered differentially abundant when |log₂ fold change| > 1 and FDR < 0.05.

#### Pathway enrichment and multi-omics integration

Pathway enrichment analyses were performed using Gene Ontology Biological Process (GOBP) and Reactome gene sets. These gene sets ressources were selected to balance curation quality, hierarchical annotation depth, genome coverage, enrichment performance and specificity, as previously benchmarked by Gable *et al.* (*37*). GOBP and Reactome gene sets were downloaded from MSigDB for *Homo Sapiens* in their curated v2026.1 release and imported into R as GMT files.

Gene set enrichment analysis was performed using clusterProfiler::GSEA (v4.18.4) with the fgsea backend. Proteomic ranked lists used the moderated limma t statistic, whereas transcriptomic ranked lists used the DESeq2 Wald statistic. Genes were ranked in decreasing order of the corresponding statistics.

GSEA was run with 50,000 random permutations, exponent set to 1, minimum gene-set size of 15, maximum gene-set size of 250, and Benjamini-Hochberg correction. Complete result tables were retained and subsequently filtered at adjusted P value < 0.05.

Over-representation analysis was performed using clusterProfiler::enricher (v4.18.4). Input lists comprised differentially abundant proteins or differentially expressed genes satisfying |log₂ fold change| > 1 and FDR < 0.05. Because ORA is sensitive to the definition of the background universe, omics-layer-specific backgrounds were restricted to features detectable under comparable experimental conditions.

For proteomics, the background was defined as the non-redundant set of gene symbols detected in the present cytoplasmic and nuclear proteomes, combined with a previously generated LFQ LC-MS/MS fibroblast proteome (Proteomic Xchange dataset number: PXD049364, (*25*)). For transcriptomics, the background was defined as the non-redundant set of genes detected in the present RNA-seq dataset, combined with a previously generated RNA-seq count matrix (SRA depository from NCBI: SUB14844065). GOBP and Reactome annotations were filtered against the corresponding background before ORA. ORA used the same gene-set size limits as GSEA. Results were retained when Benjamini-Hochberg-adjusted P value < 0.05 and the pathway overlap count was ≥15.

For each comparison and gene-set database, GSEA and ORA outputs were integrated by pathway identifier. Integrated tables retained the GSEA normalized enrichment score, GSEA adjusted P value, ORA fold enrichment, ORA adjusted P value, and overlapping genes. Pathways significant in only one enrichment framework were assigned an adjusted P value of 1 for the missing method to enable joint visualization. Integrated plots compared -log10 transformed adjusted P values from GSEA and ORA.

Pathways significant by both approaches were highlighted and grouped into broad biological categories using keyword-based annotation.

## Supplementary Figures S1 to S19

**Figure S1.**
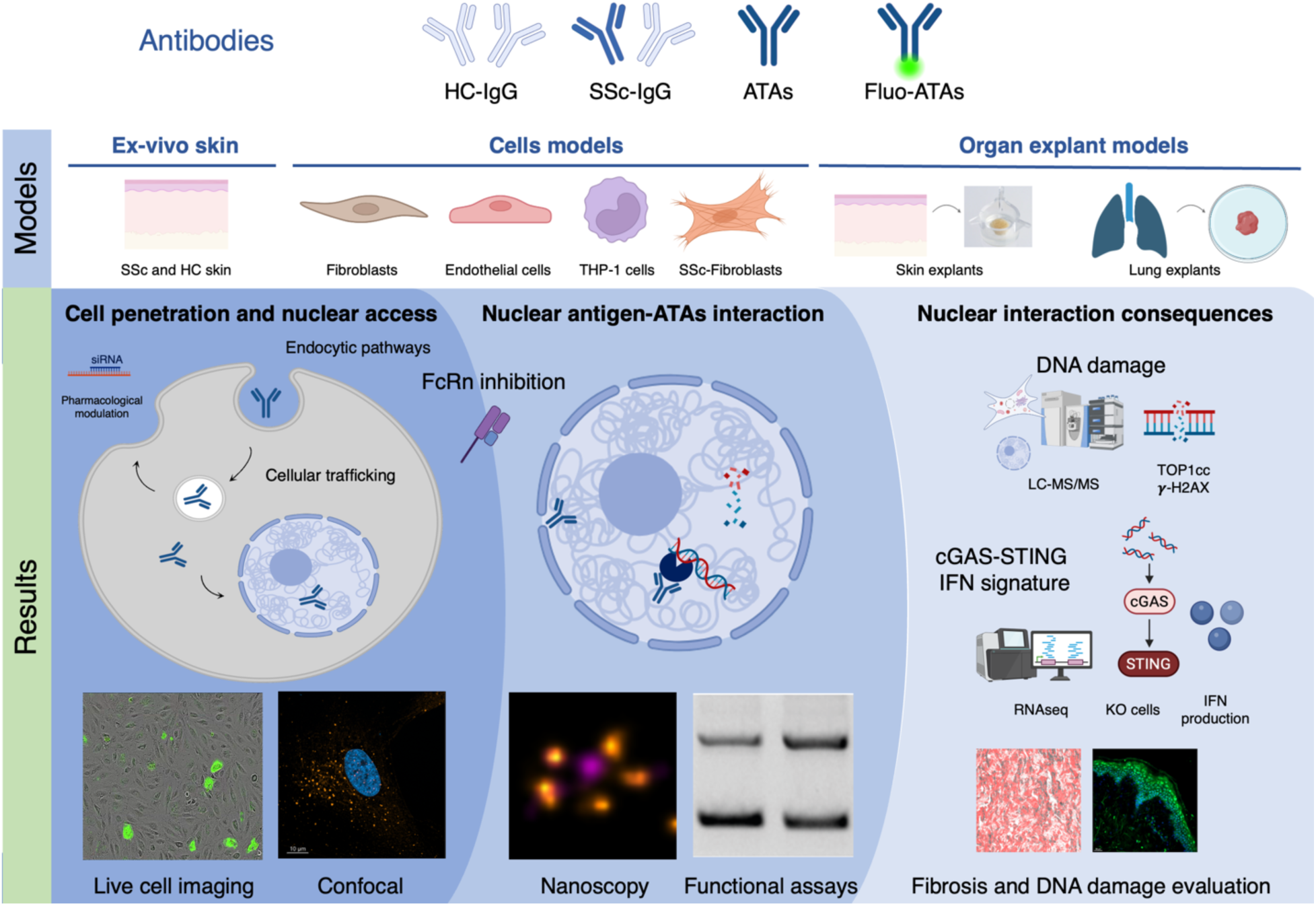
Study workflow. To investigate the capacity of ANAs to reach their nuclear targets, engage intranuclear antigens, and elicit functional consequences, we used antibodies from multiple sources: IgG from patients with systemic sclerosis (SSc-IgG) and healthy controls (HC-IgG). Total IgG were isolated from ATA-positive SSc patients (ATA^+^-IgG); from a subset of these individuals, ATAs were affinity-purified and the corresponding ATAs-depleted IgG fractions were retained. Fluorescently labeled ATAs (fluo-ATAs) and HC-IgG (fluo-HC-IgG) were also prepared. Experiments were conducted across a range of model systems, including cellular models (fibroblasts, endothelial cells, THP-1 monocytes, and SSc-derived fibroblasts), skin biopsies, and organ explant models (skin and lung). Using confocal microscopy, live-cell imaging, STED nanoscopy, functional assays (including the REEAD assay), and omics approaches, we demonstrate that ANAs enter living cells, accumulate in the nucleus, and engage their intracellular antigen. Nuclear ATAs inhibit TOP1 catalytic activity, induce DNA damage, and activate type I interferon production via the cGAS–STING pathway. We further identify neonatal Fc receptor (FcRn)-dependent intracellular trafficking as a key determinant of ANA nuclear access and show that pharmacological FcRn blockade impairs ATAs functionality.

**Figure S2.**
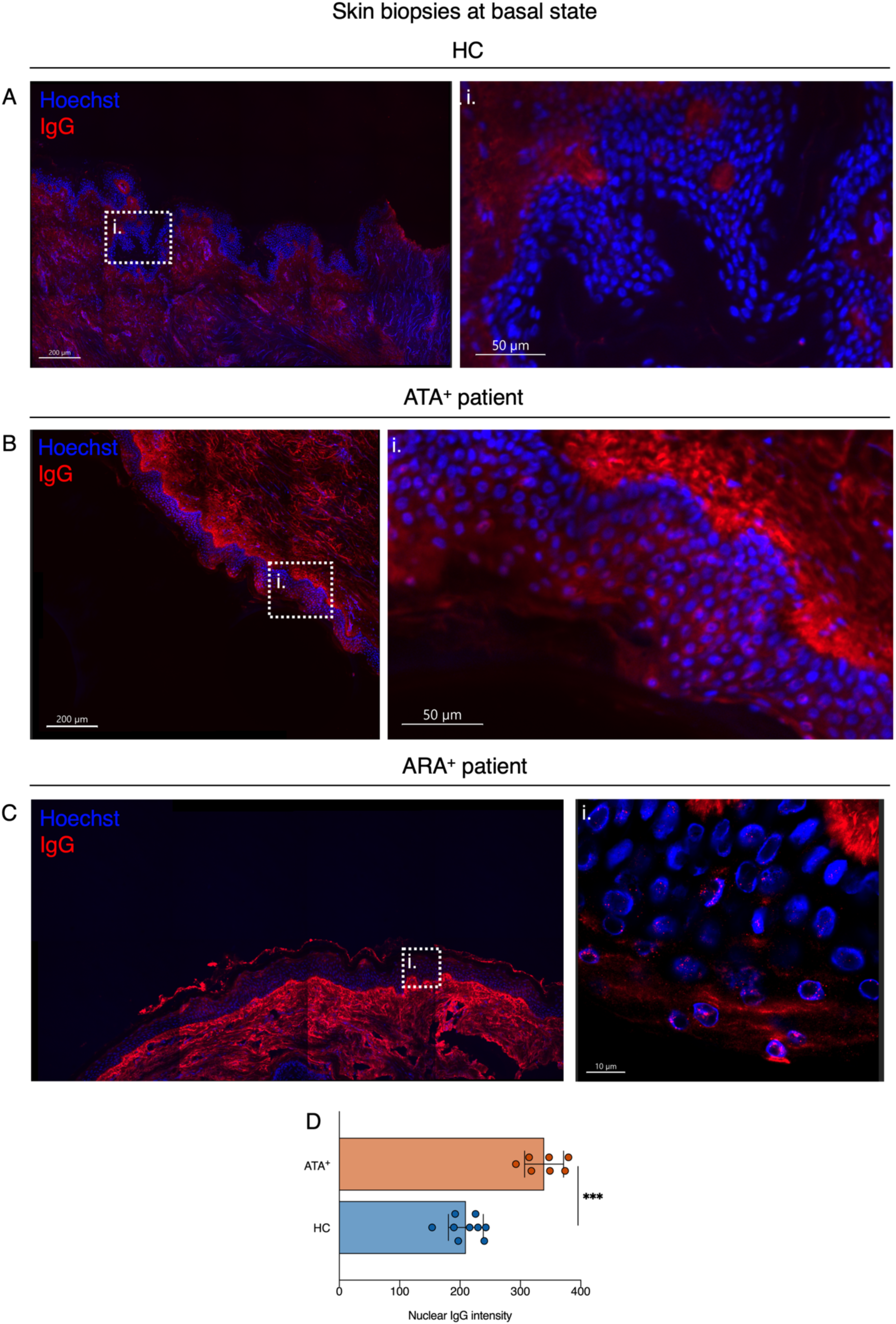
HC and SSc skins microscopy. **(A–C)** Large-scale confocal imaging of skin sections from healthy controls (HC; A), ATA-positive SSc patients (ATA^+^; B), and ARA-positive SSc patients (ARA^+^; C). Tissue sections were imaged in their entirety using a Zeiss LSM 980 confocal microscope, and full large-scale images were reconstructed by tile stitching. Human anti-IgG antibodies was used to revealed intracellular and intranuclear IgG staining In SSc. Nuclei were delimited by Hoechst staining. **(D)** Quantification of intranuclear IgG signal intensity in skin sections from SSc patients and HC. *n* = 3 subjects per condition. Box plots indicate median and full data range. ***P < 0.001 by one-way ANOVA with Tukey’s post hoc correction for multiple comparisons.

**Figure S3.**
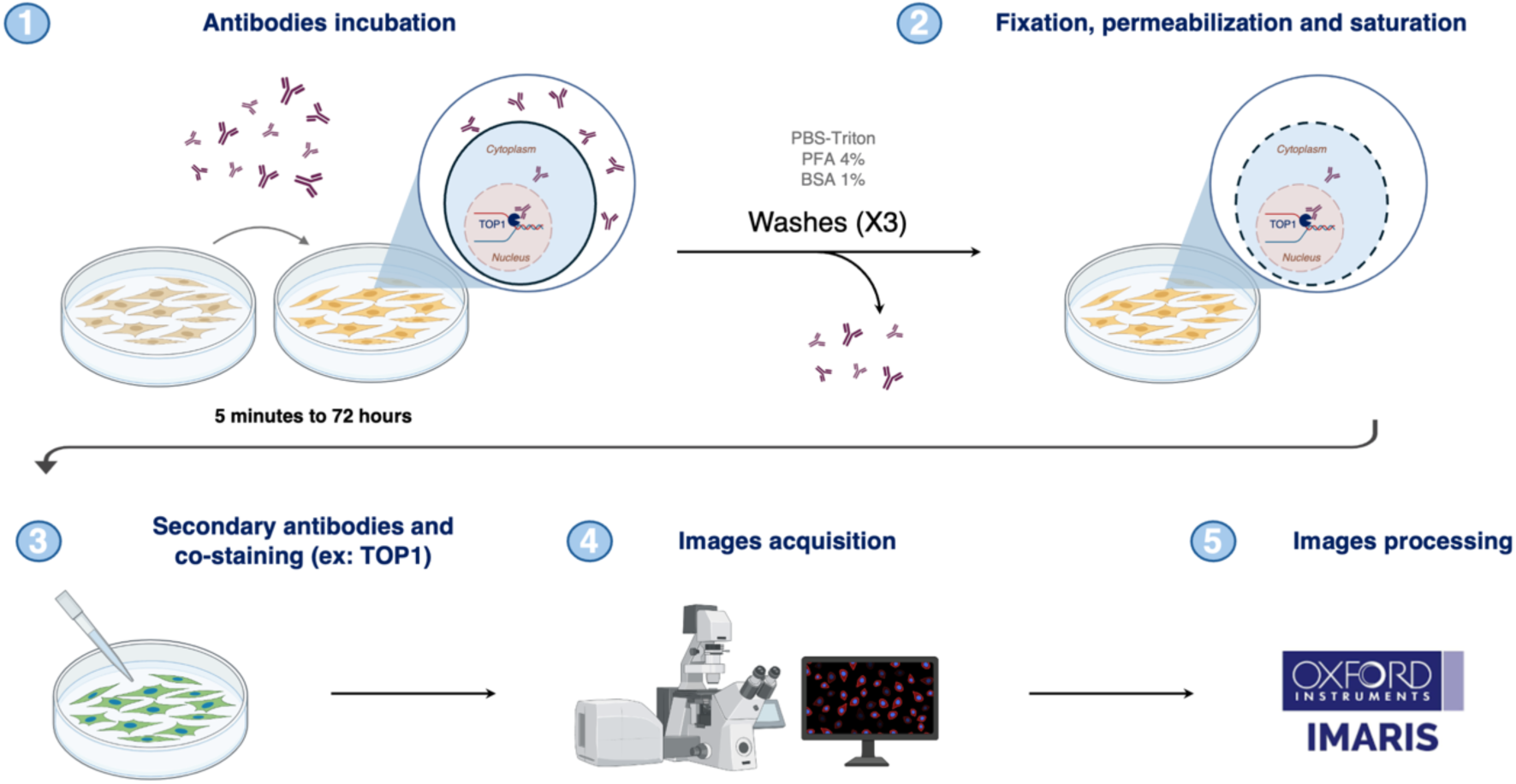
Fixed microscopy workflow for cellular models. Fibroblasts and endothelial cells were cultured in the presence of various antibodies preparations (HC-IgG, SSc-IgG, ATAs, fluo-ATAs, fluo-HC-IgG, or SLE-IgG) for durations ranging from 5 minutes to 24 hours. The medium was then aspirated and cells were washed three times to remove residual surface-bound antibodies. Cells were subsequently fixed, permeabilized, and blocked, before incubation with a human-specific secondary anti-IgG antibody together with co-staining reagents (e.g., TOP1). Under these conditions, secondary antibody labeling is restricted to cells that had internalized the primary antibodies, thereby ensuring specificity of the intranuclear signal. Images were acquired on an Airyscan confocal microscope and processed using IMARIS software.

**Figure S4.**
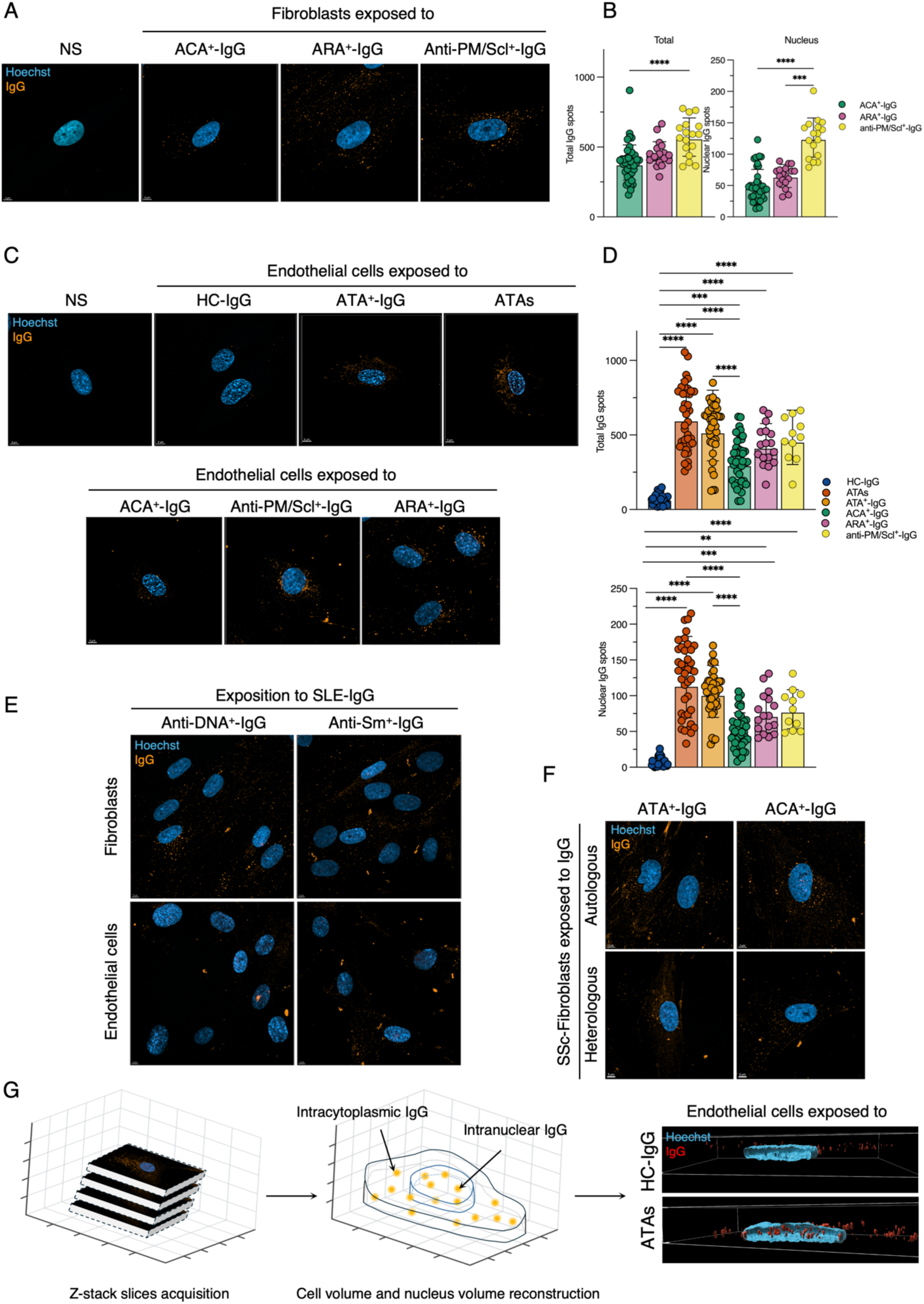
IgG enter cells and ANAs nuclei in cellular models. **(A)** Representative confocal micrographs of fibroblasts showing total intracellular and nuclear IgG staining following incubation with SSc-IgG preparations (ACA^+^-IgG, PM/Scl^+^-IgG, and ARA^+^-IgG, 100 µg/mL). NS: non-stimulated fibroblasts. **(B)** Quantification of total intracellular (left) and nuclear (right) IgG signal intensity in fibroblasts following incubation with the indicated IgG preparations. ACA^+^-IgG: 10 individuals, *n* ≥ 300 cells analyzed per condition, ARA^+^-IgG and Pm/Scl^+^-IgG: 3 individuals for each serotype, *n* ≥ 100 cells analyzed per condition. Box plots indicate median and full data range. ns: non-significant; *P < 0.05; **P < 0.01; ***P < 0.001; ****P < 0.0001; Kruskal-Wallis test with Dunn’s post hoc correction for multiple correction. **(C)** Representative confocal micrographs of endothelial cells showing total intracellular and nuclear IgG staining following incubation with HC-IgG or SSc-IgG (100 µg/mL) and ATAs (25 µg/mL). NS: non-stimulated endothelial cells. **(D)** Quantification of total intracellular (left) and nuclear (right) IgG signal intensity in endothelial cells following incubation with the indicated IgG preparations. ATA^+^-IgG, ACA^+^-IgG, HC-IgG: 10 individuals per serotype, *n* ≥ 300 cells analyzed per condition; ARA^+^-IgG and anti-PM/Scl^+^-IgG: 3 individuals per serotype, *n* ≥ 100 cells analyzed per condition; ATAs: 3 conditions, *n* ≥ 300 cells analyzed per condition. Box plots indicate median and full data range. ns: non-significant; **P < 0.01; ***P < 0.001; ****P < 0.0001; Kruskal-Wallis test with Dunn’s post hoc correction for multiple correction. **(E)** Representative confocal micrographs of fibroblasts and endothelial cells exposed to SLE-IgG preparations (IgG-anti-dsDNA^+^ and IgG-anti-Sm^+^, 100 µg/mL). **(F)** Representative confocal micrographs of SSc-derived fibroblasts exposed to autologous (i.e. fibroblasts and IgG from the same patient) and heterologous (i.e. fibroblasts and IgG from different patients) SSc-IgG. **(G)** Z-stack image acquisition workflow and representative Z-stack images of endothelial cell nuclei following 1-hour exposure to HC-IgG (100 µg/mL) or ATAs (25 µg/mL). All fluorescence images were acquired using an Airyscan confocal microscope (∼140 nm lateral resolution).

**Figure S5.**
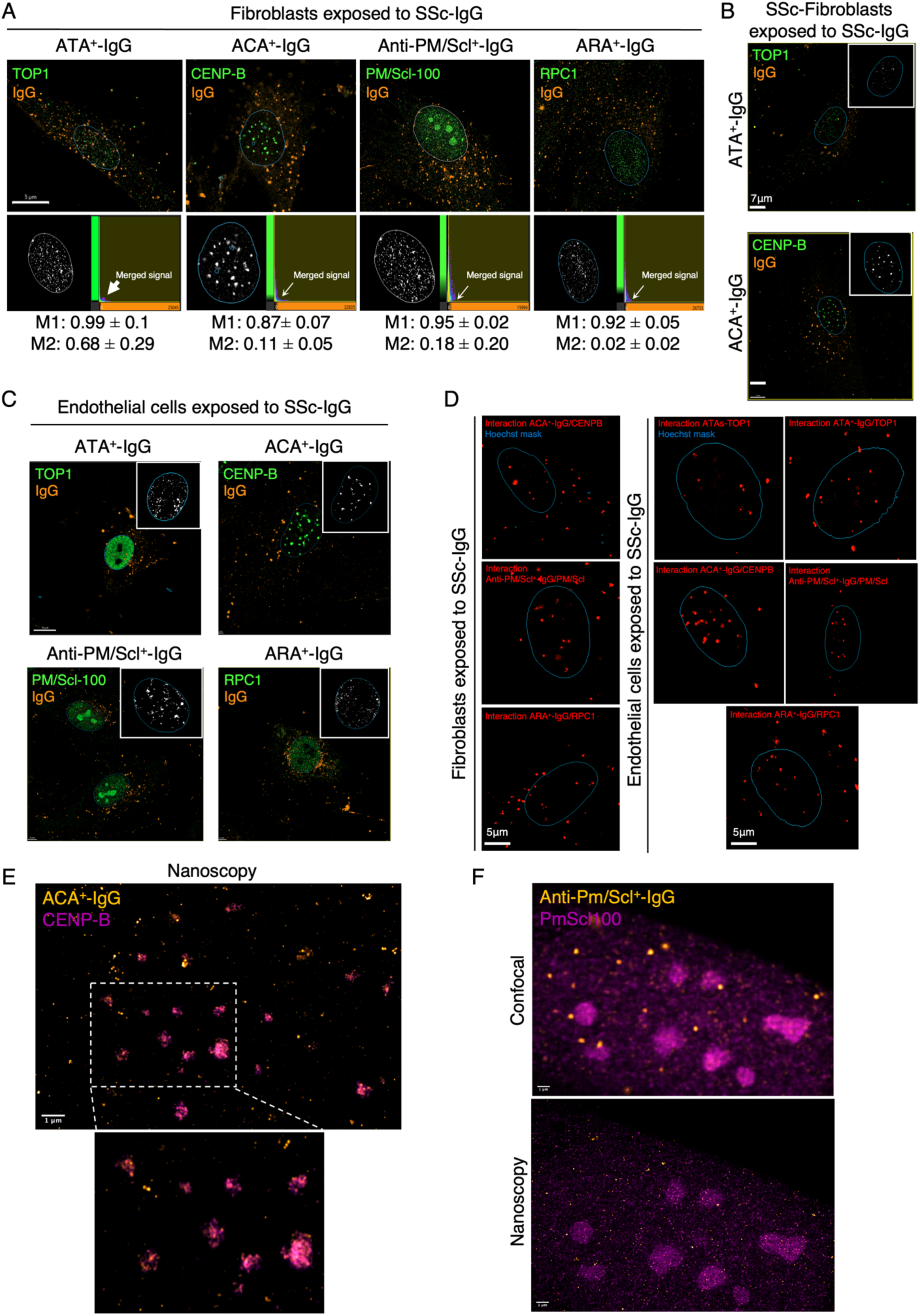
ANAs interacts with their intracellular antigen. **(A)** Representative confocal micrographs showing antigen-IgG colocalization in fibroblasts exposed to SSc-IgG (100 µg/mL) for 1 hour. Colocalization was quantified using the Manders overlap coefficient. M1 and M2 values are expressed as median ± interquartile range. **(B)** Representative confocal micrographs showing antigen-IgG colocalization in SSc-derived fibroblasts (SSc-FB) exposed to SSc-IgG (100 µg/mL) for 1 hour. **(C)** Representative confocal micrographs showing antigen-IgG colocalization in endothelial cells exposed to SSc-IgG (100 µg/mL) for 1 hour. **(D)** Proximity ligation assay (PLA) of target antigens and their corresponding SSc-IgG in fibroblasts (left) and endothelial cells (right) exposed to SSc-IgG (100 µg/mL) or ATAs (25 µg/mL) for 1 hour. Individual fluorescent spots indicate antigen-IgG interactions occurring within 40 nm. **(E-F)** STED (stimulated emission depletion) nanoscopy revealing nuclear interaction between CENP-B and ACA^+^-IgG (**E**), and between PM/Scl-100 and Anti-PM/Scl^+^-IgG (**F**), in fibroblasts exposed to SSc-IgG for 1 hour. All fluorescence images were acquired using an Airyscan confocal microscope (∼140 nm lateral resolution). Super-resolution imaging in (**E**) and **(F**) were performed by stimulated emission depletion (STED) microscopy (∼41 nm lateral resolution).

**Figure S6.**
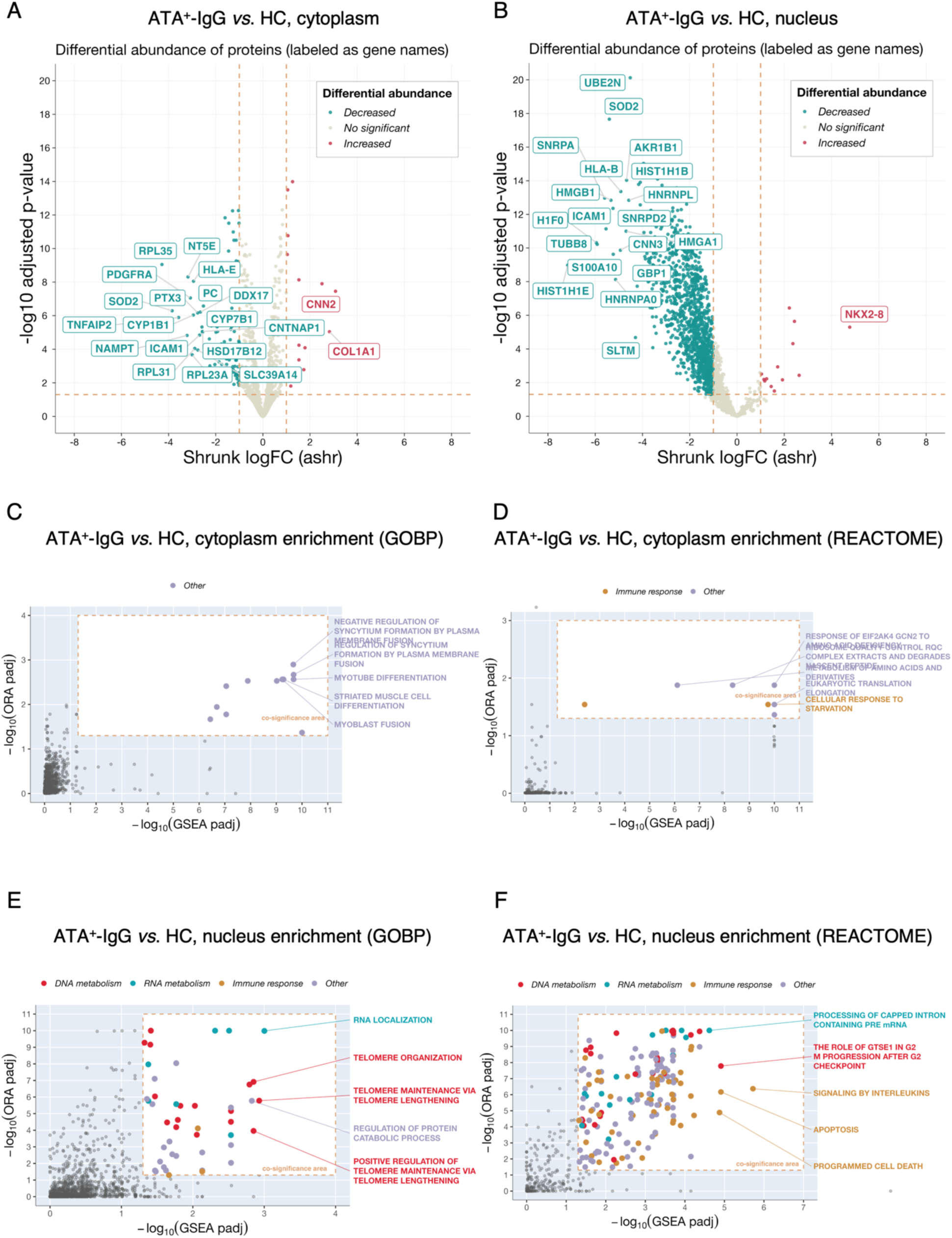
Cytoplasmic and nuclear proteomic profiles of fibroblasts exposed to ATAs. **(A–B)** Volcano plots of differentially expressed proteins in ATA^+^-IgG versus HC-IgG exposed fibroblasts, in cytoplasmic (A) and nuclear (B) fractions. Significantly dysregulated proteins are labeled by gene name (adjusted P < 0.05; |log₂ fold change| > 1). **(C–D)** Enrichment analysis of dysregulated proteins in the cytoplasmic fraction (ATA^+^-IgG *vs*. HC-IgG). The plot displays pathways reaching significance in both over-representation analysis (ORA; y-axis) and gene set enrichment analysis (GSEA; x-axis), using the GO Biological Process (C) and Reactome (D) databases as background. Only the five most significant co-enriched pathways are labeled. **(E–F)** Enrichment analysis of dysregulated proteins in the nuclear fraction (ATA^+^-IgG *vs*. HC-IgG). The plot displays pathways reaching significance in both over-representation analysis (ORA; y-axis) and gene set enrichment analysis (GSEA; x-axis), using the GO Biological Process (E) and Reactome (F) databases as background. Only the five most significant co-enriched pathways are labeled.

**Figure S7.**
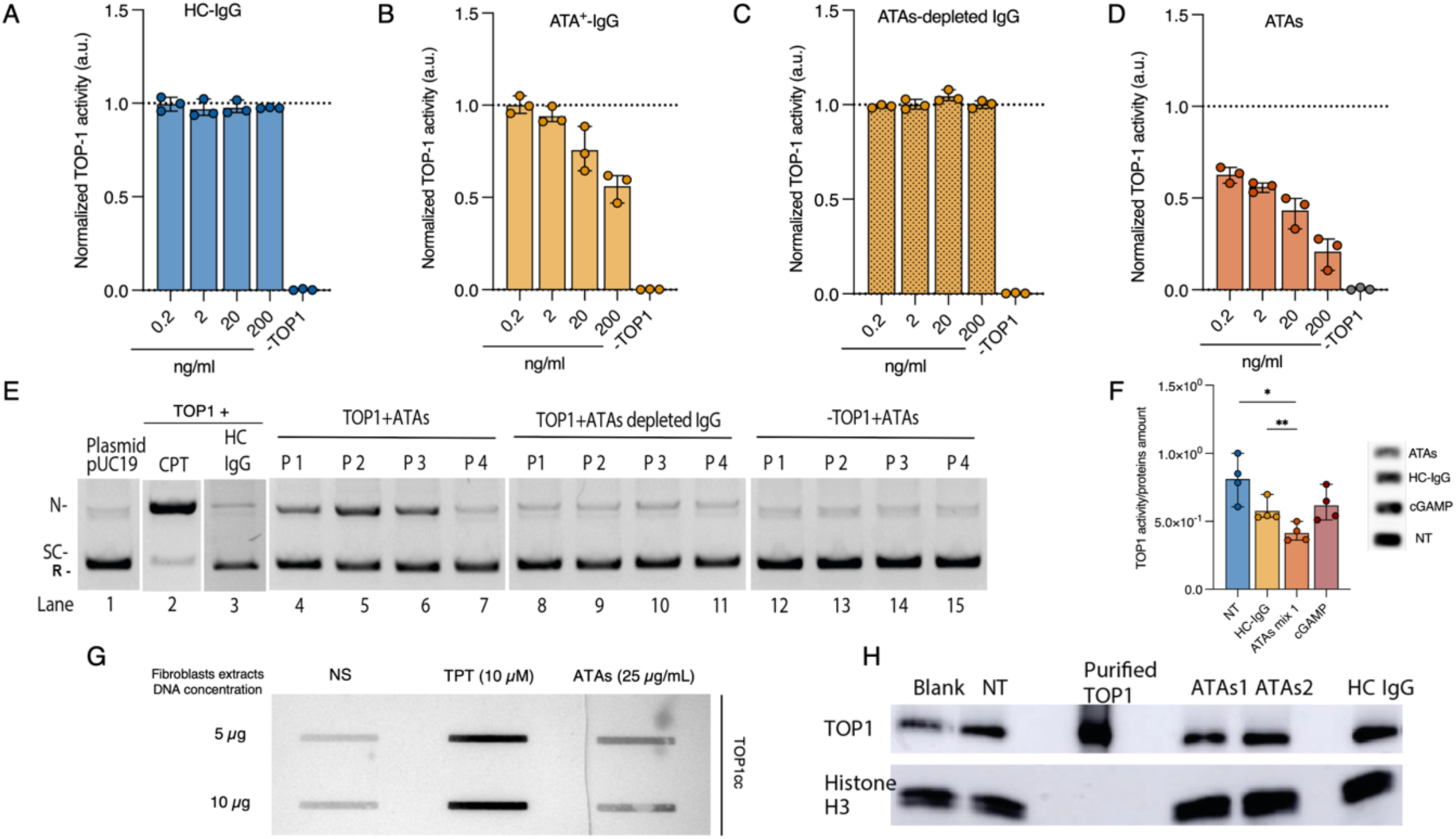
Bioactivity of ATA^+^-IgG, and ATAs. **(A–D)** Topoisomerase I (TOP1) activity measured by REEAD assay in the presence of HC-IgG, ATA^+^-IgG, ATAs-depleted IgG, or ATAs. TOP1 inhibition was observed exclusively in the ATA^+^-IgG and ATA conditions and was retained in the ATA-depleted IgG fraction (*n* = 3 per condition). **(E)** DNA Nicking assay in the presence of HC-IgG, ATA^+^-IgG, from patients (P1-27, *n* = 8), or ATA-depleted IgG, demonstrating dose-dependent TOP1 inhibition restricted to the ATA. The mobility of supercoiled plasmid DNA is indicated to the left of the figure (SC) as is the mobility of relaxed plasmid DNA (R). **(F)** TOP1 activity measured by REEAD assay in whole cell extract from endothelial cells (HUVECs) following exposure to ATAs, demonstrating a reduction in TOP1 catalytic activity. *n* = 4 per condition. **(G)** Rapid Approach to DNA Adduct Recovery (RADAR) Assay showing TOP1-DNA cleavage complex (TOP1cc) in DNA extracted from fibroblasts treated with topotecan (TPT) or ATAs. NS, non-stimulated fibroblasts. **(H)** TOP1 protein expression in isolated nuclei from endothelial cells exposed to HC-IgG or ATAs (10 µg/ml) for 16h, showing no significant changes in expression levels.

**Figure S8.**
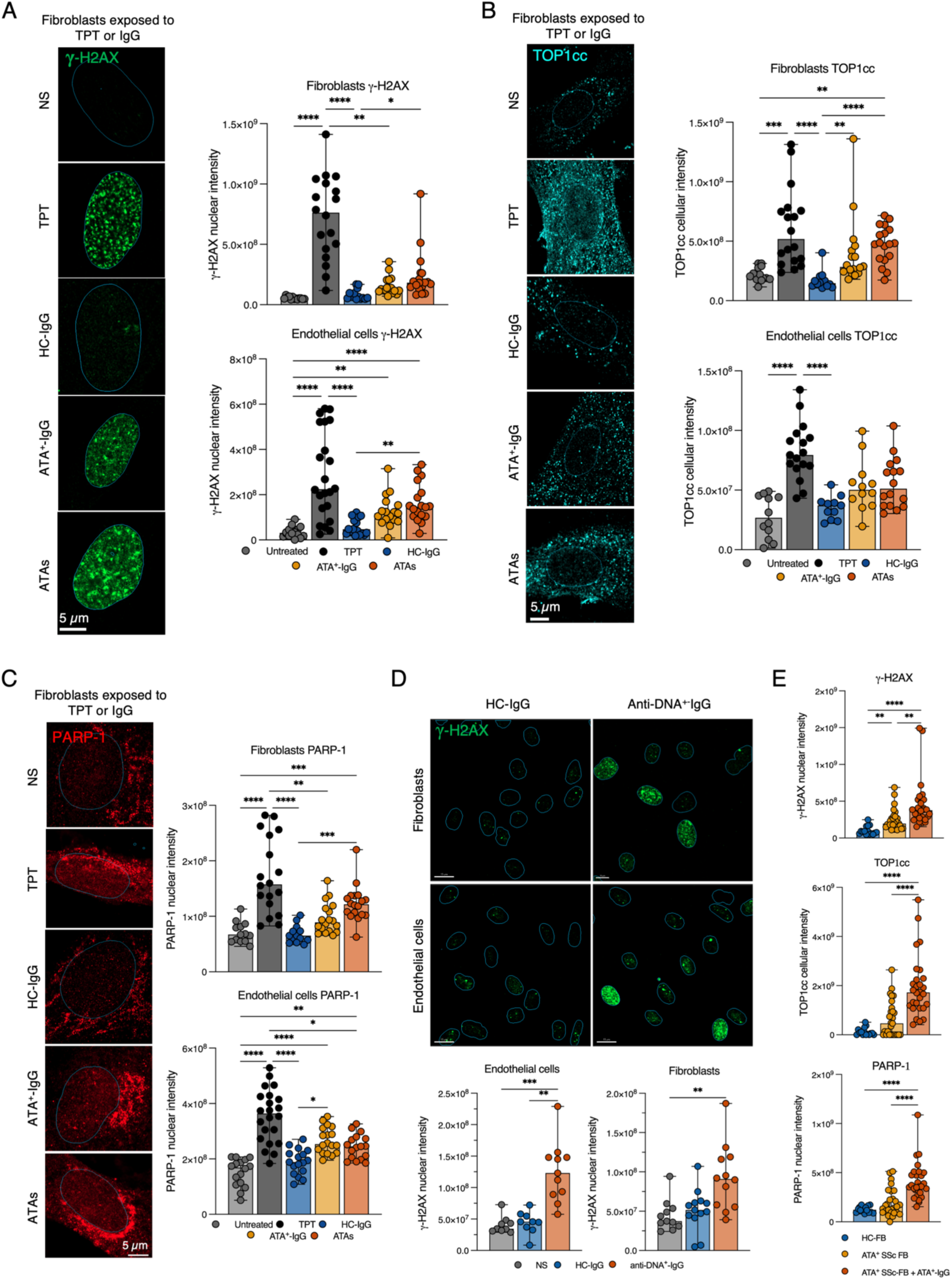
Induction of DNA damages and repair, TOP1cc production in cellular models. **(A)** Representative confocal micrographs (left) and quantification (right) of γ-H2AX staining in fibroblasts exposed for 24h to topotecan (TPT, 2 µM), HC-IgG, ATA^+^-IgG or ATAs (100 µg/mL for total purified IgG; 25 µg/mL for ATAs). Box plots indicate median and full data range. *P < 0.05; **P < 0.01; ***P < 0.001; Kruskal-Wallis test with Dunn’s post hoc correction for multiple comparisons. **(B)** Representative confocal micrographs (left) and quantification (right) of TOP1cc staining in fibroblasts exposed for 24 hours to TPT (2 µM), HC-IgG, ATA^+^-IgG or ATAs (100 µg/mL for total purified IgG; 25 µg/mL for ATAs). Box plots indicate median and full data range. *P < 0.05; **P < 0.01; ***P < 0.001; Kruskal-Wallis test with Dunn’s post hoc correction for multiple comparisons. **(C)** Representative confocal micrographs (left) and quantification (right) of PARP-1 staining in fibroblasts exposed for 24 hours to TPT (2 µM), HC-IgG, ATA^+^-IgG or ATAs (100 µg/mL for total purified IgG; 25 µg/mL for ATAs). Box plots indicate median and full data range. *P < 0.05; **P < 0.01; ***P < 0.001; Kruskal-Wallis test with Dunn’s post hoc correction for multiple comparisons. **(D)** Representative confocal micrographs (top) and quantification of γ-H2AX (bottom) in fibroblasts exposed for 24 hours to HC-IgG or anti-dsDNA^+^-IgG (100 µg/mL). Box plots indicate median and full data range. *P < 0.05; **P < 0.01; ***P < 0.001; Kruskal-Wallis test with Dunn’s post hoc correction for multiple comparisons. **(E)** Quantification of γ-H2AX, TOP1cc, and PARP-1 levels in HC fibroblasts (HC-FB), fibroblasts from an ATA^+^ SSc-patient (ATA^+^ SSc-FB), or ATA^+^ SSc-FB additionally exposed to ATA^+^-IgG (100 µg/mL) for 24 hours. Box plots indicate median and full data range. *P < 0.05; **P < 0.01; ***P < 0.001; Kruskal-Wallis test with Dunn’s post hoc correction for multiple comparisons.

**Figure S9.**
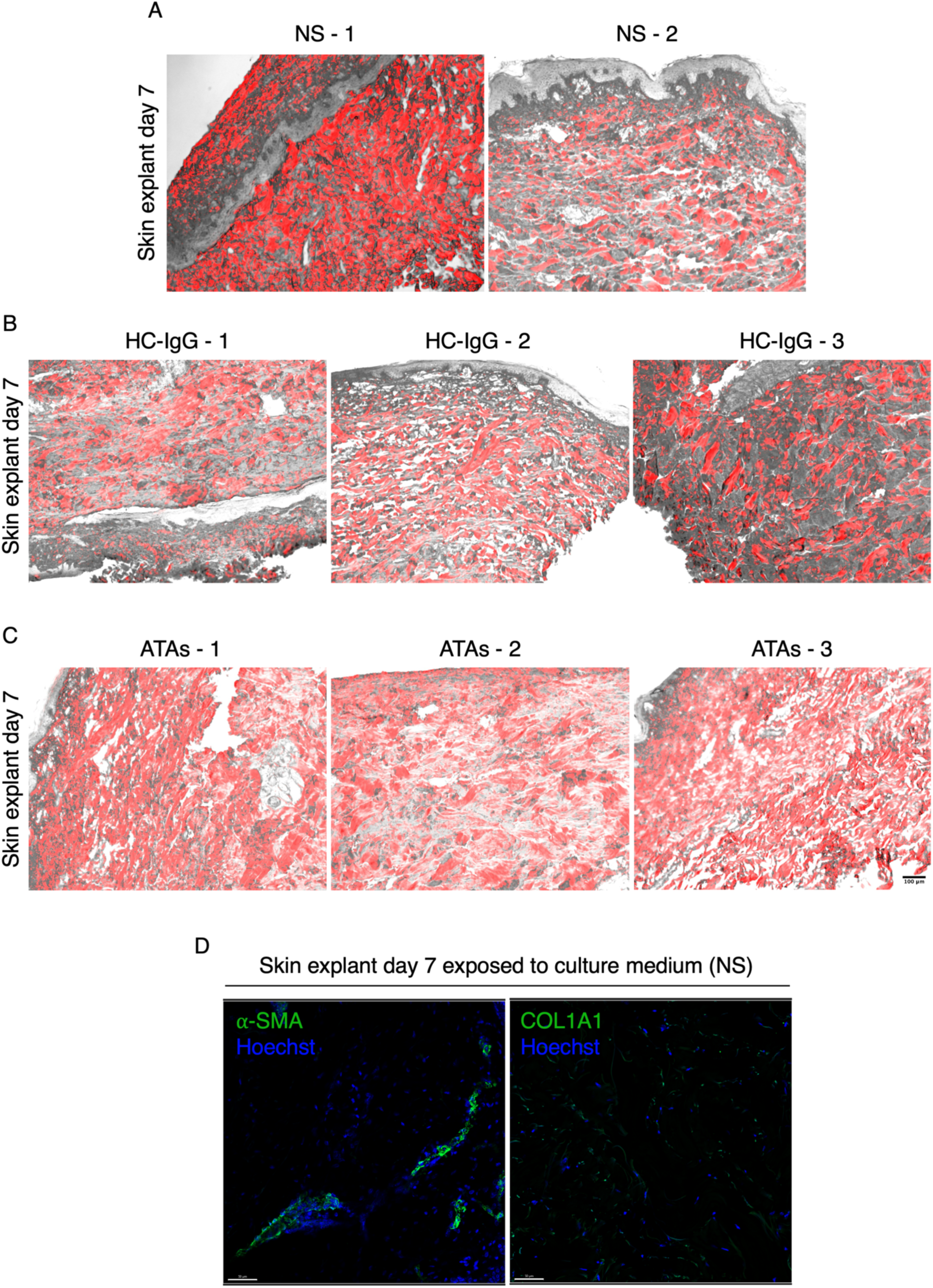
ATA induced fibrosis in skin explant models. **(A–C)** Representative images of collagen deposition assessed by picrosirius red staining in skin explants exposed for 7 days to culture medium alone (NS; A), HC-IgG (B), or ATAs (C). **(D)** Representative confocal micrographs showing COL1A1 and α-smooth muscle actin (α-SMA) expression in skin explants exposed to culture medium alone (NS) for 7 days.

**Figure S10.**
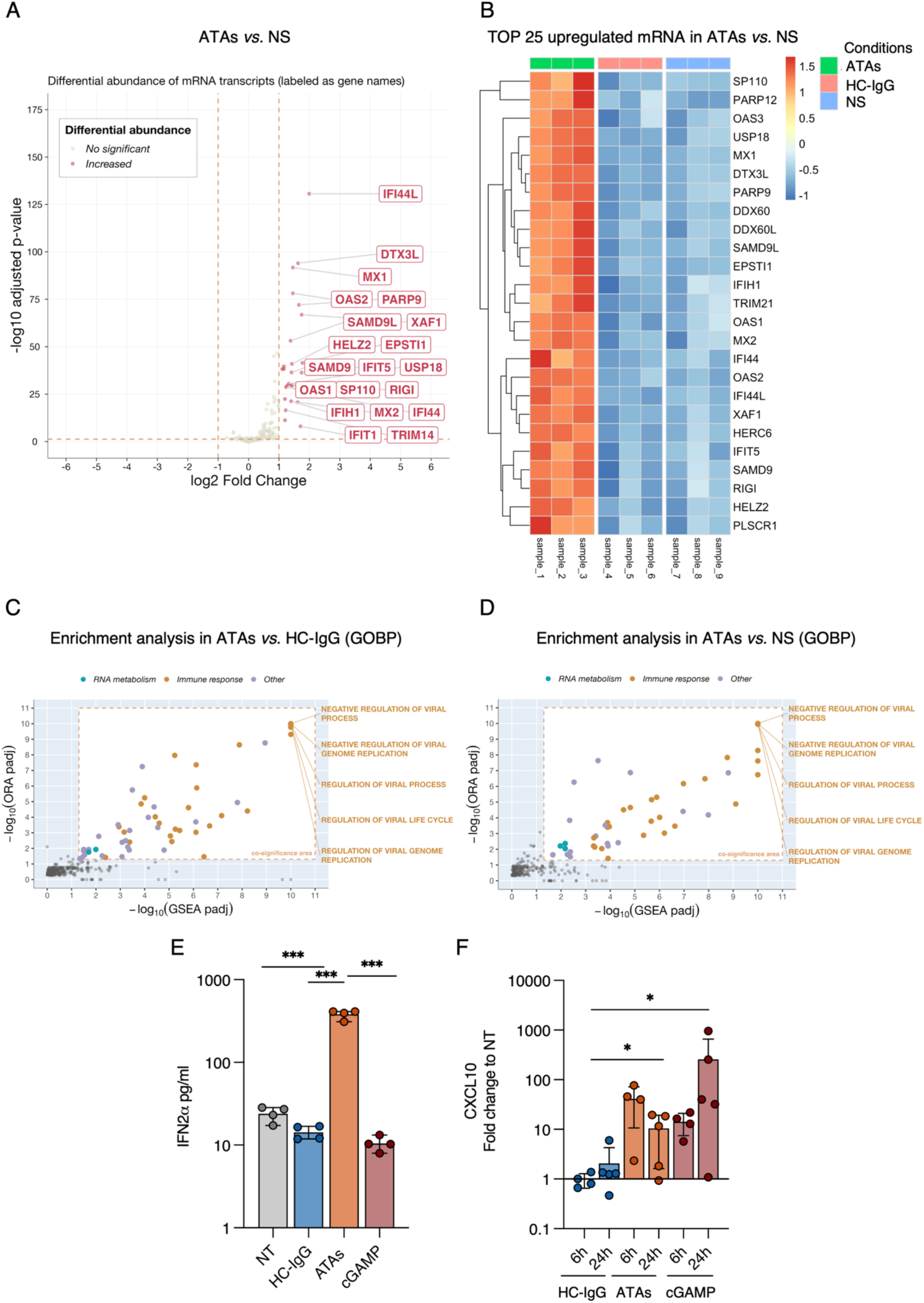
ATAs induce IFN signature in cells and lung explants. **(A)** Volcano plot showing differential mRNA expressions in endothelial cells pretreated with IFN-α and exposed to ATAs versus NS. Significantly dysregulated genes are labeled (adjusted P < 0.05; log fold change > 1 or < −1). NS: non-simulated. **(B)** Heatmap of the top 25 dysregulated mRNAs in ATAs, HC-IgG, and NS based on the ATA versus NS comparison (adjusted P < 0.05; log fold change > 1 or < −1). NS: non-simulated. **(C-D)** Enrichment analysis of mRNAs upregulated in ATAs-versus HC-IgG-treated (C) and ATAs-versus NS comparisons. The plot displays pathways reaching significance in both over-representation analysis (ORA; y-axis) and gene set enrichment analysis (GSEA; x-axis), using the Reactome database as background. Only the five most significant co-enriched pathways are labeled. **(E)** Interferon-2-α secretion in endothelial cells exposed to HC-IgG, ATAs or cGAMP. n = 4 per condition; box plots show median and full data range. *P < 0.05, **P < 0.01, ***P < 0.001, ****P < 0.0001 by Kruskal-Wallis test with Dunn’s post hoc correction for multiple comparison. **(F)** CXCL10 production measured in lung explants supernatant exposed to HC-IgG, ATAs or cGAMP. n = 4 per condition; box plots show median and full data range. *P < 0.05 by Kruskal-Wallis test with Dunn’s post hoc correction for multiple comparisons.

**Figure S11.**
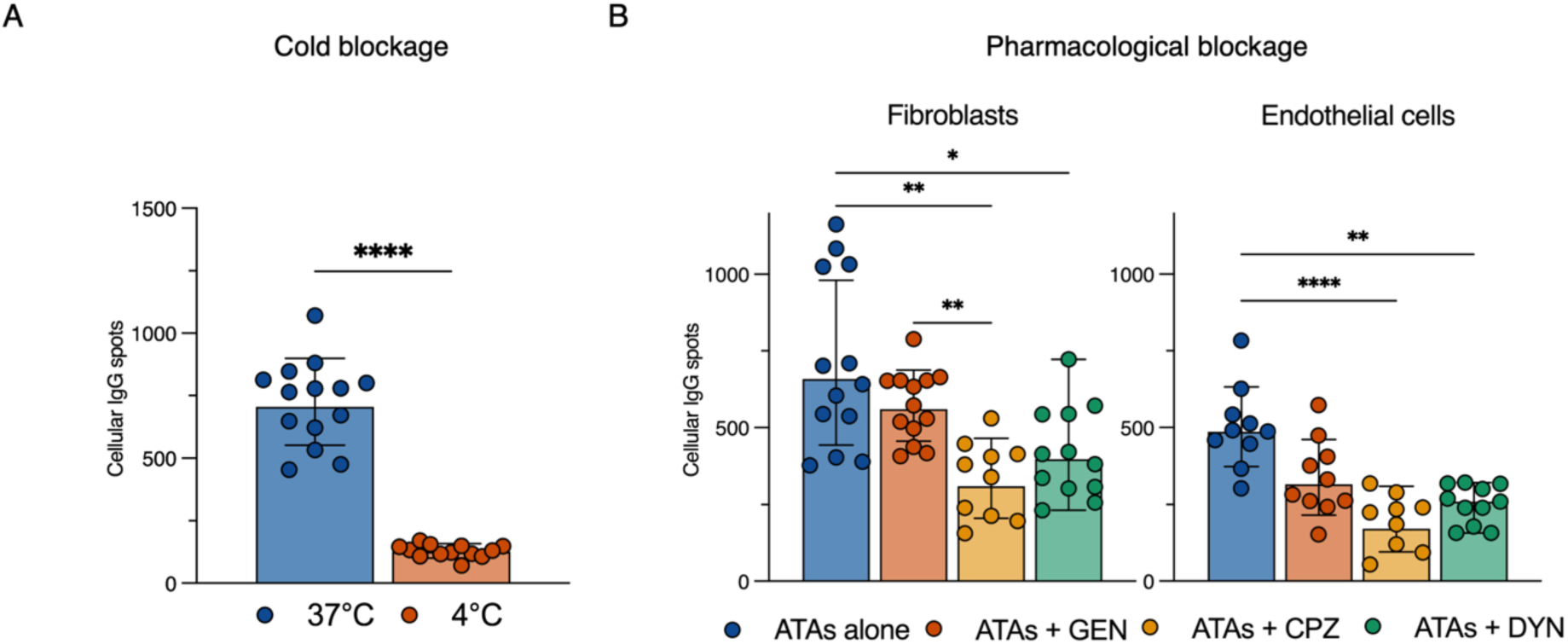
Cold blockage and pharmacological blockage of endocytosis. **(A)** Quantification of cytoplasmic and nuclear IgG fluorescent spots in fibroblasts exposed to ATAs at 37°C or 4°C. *n* = 3 experiments with *n* ≥ 100 cells analyzed per condition. Box plots indicate median and full data range. ****P < 0.0001 by Mann-Whitney U test for pairwise comparisons. **(B)** Quantification of intracellular IgG fluorescent spots in fibroblasts and endothelial cells exposed to ATAs (25 µg/mL) following pharmacological inhibition of the caveolin-mediated endocytic pathway (genistein, GEN) or the clathrin-mediated endocytic pathway (chlorpromazine, CPZ; dynasore, DYN). *n* = 3 experiments with *n* ≥ 100 cells analyzed per condition. Box plots indicate median and full data range. ns: non-significant; *P < 0.05; **P < 0.01; ***P < 0.001; ****P < 0.0001; Kruskal-Wallis test with Dunn’s post hoc correction for multiple comparisons.

**Figure S12.**
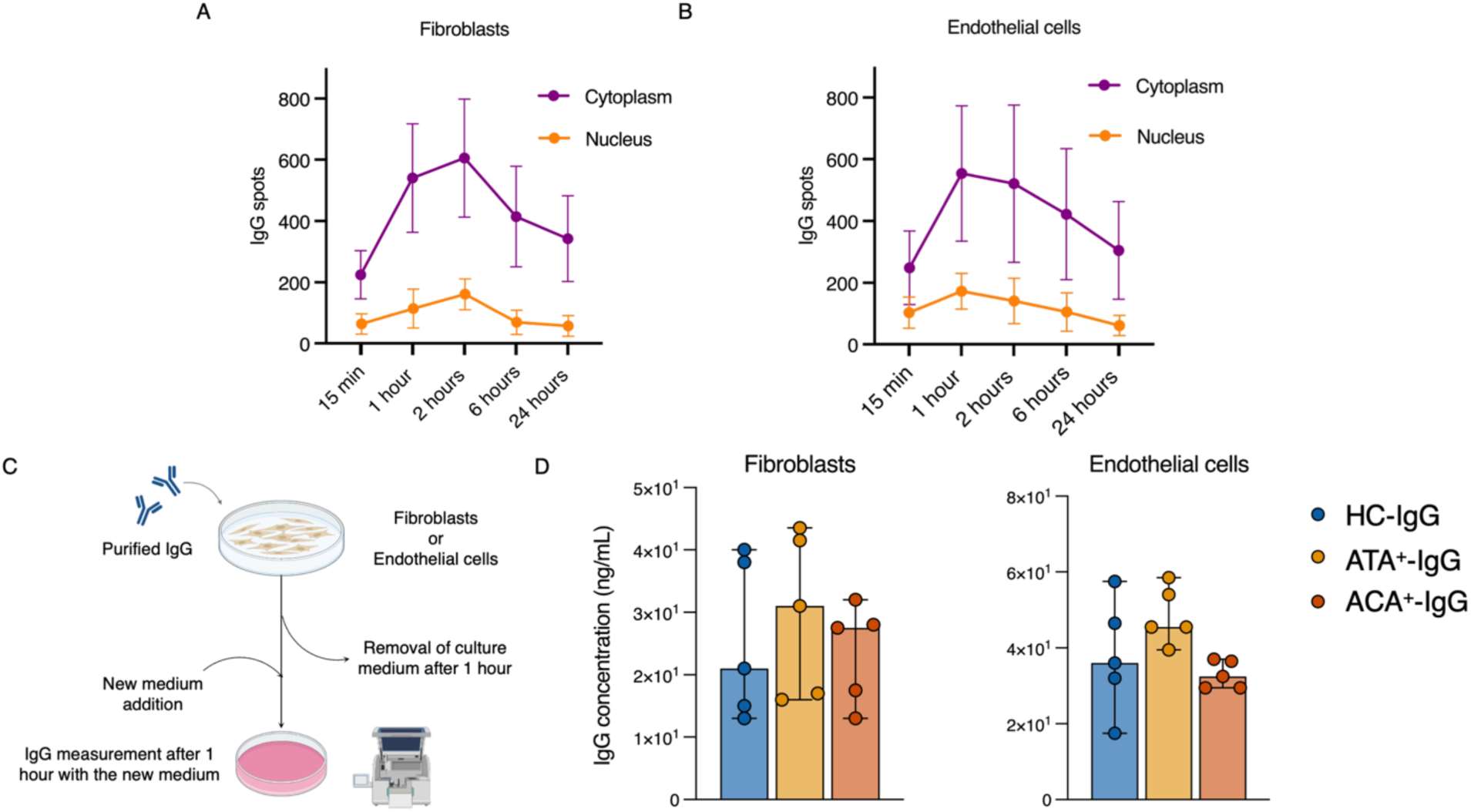
IgG uptake and recycling in cells. **(A–B)** Kinetics of cytoplasmic and nuclear IgG uptake in fibroblasts (A) and endothelial cells (B) following exposure to ATAs (25 µg/mL). **(C)** Cellular IgG recycling assay demonstrating sustained IgG uptake and intracellular recycling. Briefly, cells were exposed to HC-IgG or ATA^+^-IgG or ACA^+^-IgG (100 µg/mL) for 1 hour, after which the medium was removed and replaced with fresh medium. IgG concentration in the conditioned medium was measured by ELISA 1 hour after medium exchange. **(D)** IgG recycling assay in fibroblasts (left) and endothelial cells (right). Recycled IgG were measured in the conditioned medium by ELISA 1 hour after medium exchange.

**Figure S13.**
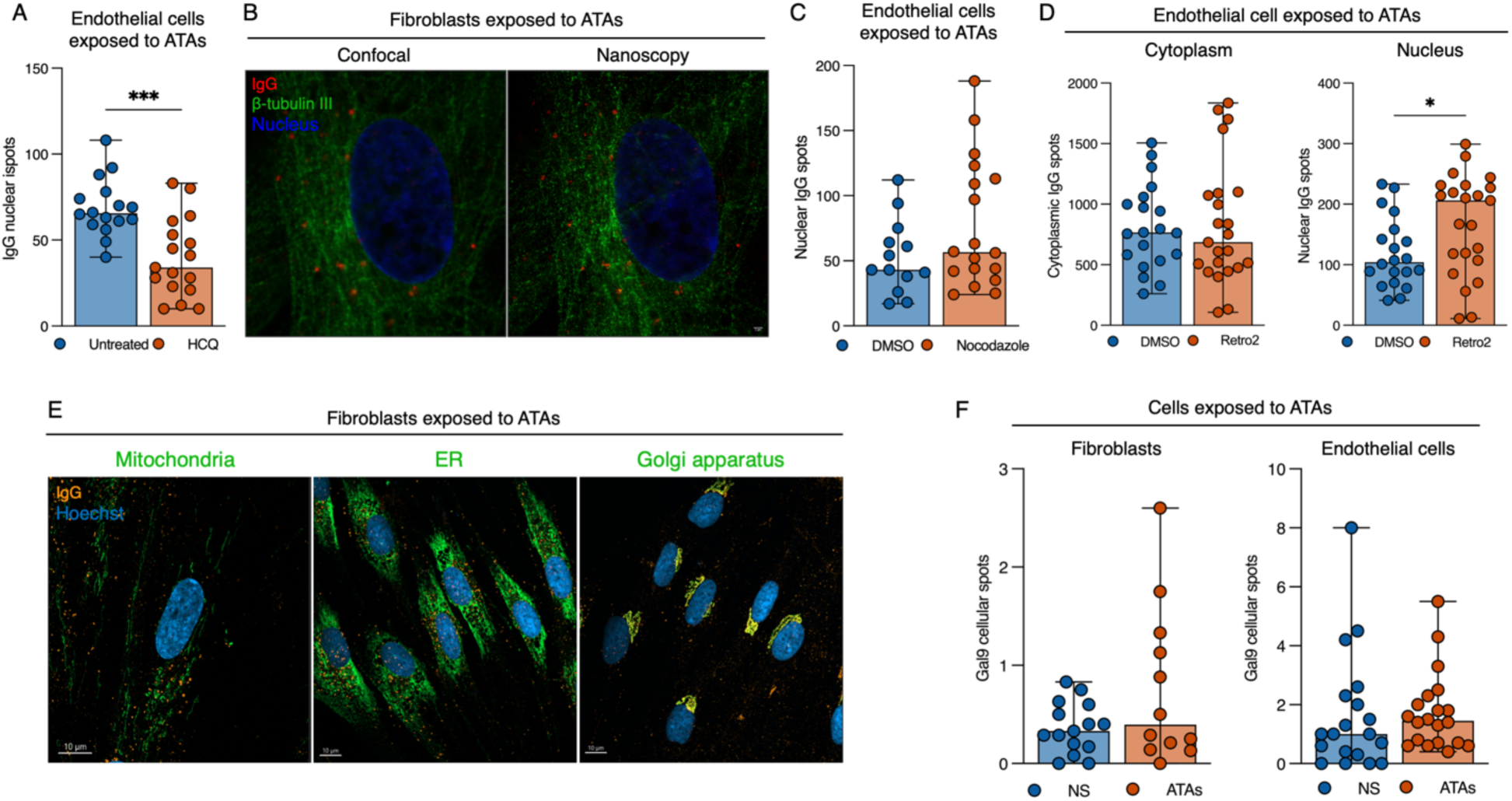
ATAs intracellular trafficking. (A) quantification of intranuclear IgG spots (right) in endothelial cells exposed to ATAs for 1 hour following inhibition of endosomal acidification and maturation with hydroxychloroquine (HCQ, 20 µM) for 2 hours at 37°C. *n* = 3 experiments per condition, *n* ≥ 100 cells per condition. Box plots indicate median and full data range. *P < 0.05; Mann-Whitney U test (B) â-tubulin III staining (confocal microscopy left; nanoscopy: right) in fibroblasts exposed to ATAs (25 µg/mL) for 20 hours. (C) quantification of intranuclear IgG spots in endothelial cells following pharmacological microtubule disruption with nocodazole (10 µM, 37°C) for 1 hour prior to ATAs exposure (25 µg/mL) for an additional hour. β-tubulin III was used as a microtubule integrity marker. *n* = 3 experiments per condition, *n* ≥ 100 cells per condition. Box plots indicate median and full data range. ns: non-significant; Mann-Whitney U test. (D) Quantification of cytoplasmic (left) and nuclear (right) IgG spots in endothelial cells exposed to ATAs (25 µg/mL) for 1 hour following pharmacological inhibition of Golgi retrograde transport with Retro-2 (20 µM) for 1 hour. *n* = 3 experiments per condition, *n* ≥ 100 cells per condition. Box plots indicate median and full data range. ns: non-significant; **P < 0.01; Mann-Whitney U test. (E) Representative confocal micrographs of ATAs localization within cell compartments of fibroblasts after 1 hour of ATAs (25 µg/mL) exposure: mitochondria, endoplasmic reticulum and Golgi apparatus. (F) Quantification of galectine-9 (GAL9) in fibroblasts (left) and endothelial cells (right) exposed to ATA (25 µg/mL) for 1 hour. *n* = 3 experiments per condition, *n* = 30 cells per experiment. Box plots indicate median and full data range. ns: non-significant; **P < 0.01; Mann-Whitney U test.

**Figure S14.**
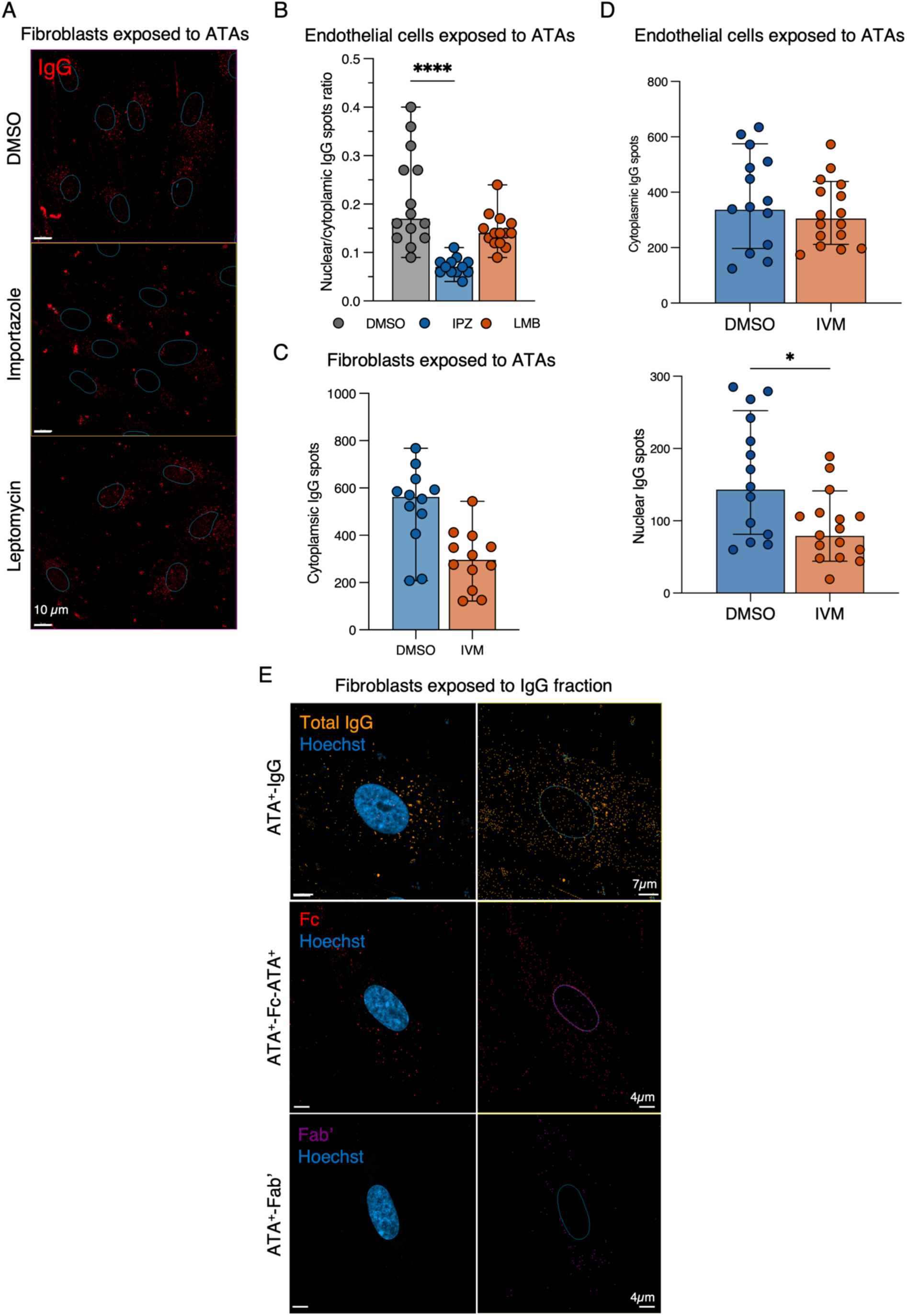
ATAs nuclear importation. **(A)** Representative confocal microscopy of fibroblasts exposed 1h to ATAs (25 µg/mL) after DMSO, importazole of Leptomycin pretreatment. **(B)** Quantification of nuclear ATAs localization following treatment with nuclear import or export inhibitors in endothelial cells. Importazole (IPZ, 20 µM) was used as a nuclear import inhibitor and leptomycin B (LMB, 20 nM) as a nuclear export inhibitor. Cells were treated with IPZ or LMB for 2 hours at 37°C prior to ATAs exposure (25 µg/mL) for an additional hour. Results are expressed as the nuclear-to-cytoplasmic spot ratio. *n* = 3 experiments per condition, *n* ≥ 100 cells per condition. Box plots indicate median and full data range. ns: non-significant; ***P < 0.001; Mann-Whitney U test. **(C)** Quantification of intranuclear IgG staining in fibroblasts exposed to ATAs (100 µg/mL) following pretreatment with the importin-α inhibitor ivermectin (10 µM). *n* = 3 experiments per condition, *n* ≥ 100 cells per condition. Box plots indicate median and full data range. ns: non-significant; **P < 0.01; Mann-Whitney U test. Importin-α inhibition impaired nuclear ATAs uptake. **(D)** Quantification of intracytoplasmic (top) and intranuclear (bottom) IgG staining in endothelial cells exposed to ATAs (100 µg/mL) following pretreatment with the importin-α inhibitor ivermectin (10 µM). *n* = 3 experiments per condition, *n* ≥ 100 cells per condition. Box plots indicate median and full data range. ns: non-significant; **P < 0.01; Mann-Whitney U test. Importin-α inhibition impaired nuclear ATAs uptake. **(E)** Representative confocal micrographs of fibroblasts exposed 1 hour to ATA^+^-IgG, ATA^+^-Fc or ATA^+^-Fab’. All fluorescence images were acquired using an Airyscan confocal microscope (∼140 nm lateral resolution).

**Figure S15.**
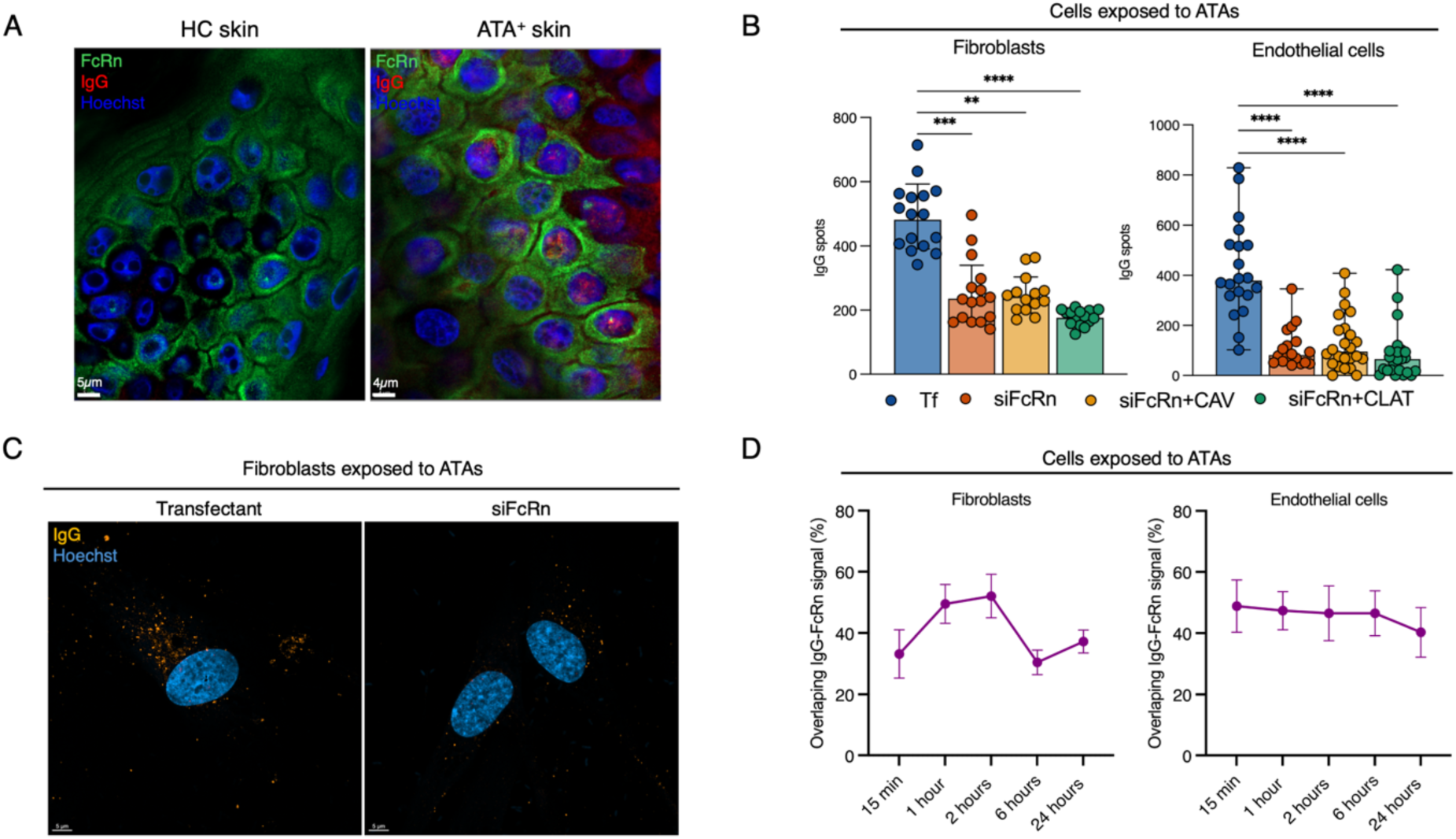
FcRn involvement. **(A)** Representative confocal microscopy of IgG and FcRn staining in HC and SSc skin samples. **(B)** Quantification of IgG staining in fibroblasts following exposure to ATAs (25 µg/mL) for 1 hour after pretreatment with FcRn siRNA, FcRn+caveolin (CAV) siRNA or FcRn+clathrin (CLAT) siRNA. *n* ≥ 100 cells analyzed per condition, *n* = 3 experiments per condition. Box plots show median, interquartile range, and full data range. *P < 0.05, **P < 0.01, ***P < 0.001 by Kruskal-Wallis test with Dunn’s post hoc correction for multiple comparison. **(C)** Representative confocal micrographs of IgG staining in fibroblasts following exposure to ATAs (25 µg/mL) for 1 hour after pretreatment with FcRn siRNA. **(D)** Time-lapse quantification of ATAs and FcRn distribution in cytoplasm and nucleus in fibroblasts (left) and endothelial cells (right) at 15 minutes, 1, 2, 6, and 24 hours after ATAs exposure. The y-axis represents the percentage of overlapping between IgG and FcRn signals. *n* ≥ 100 cells analyzed per condition, *n* = 3 experiments per condition.

**Figure S16.**
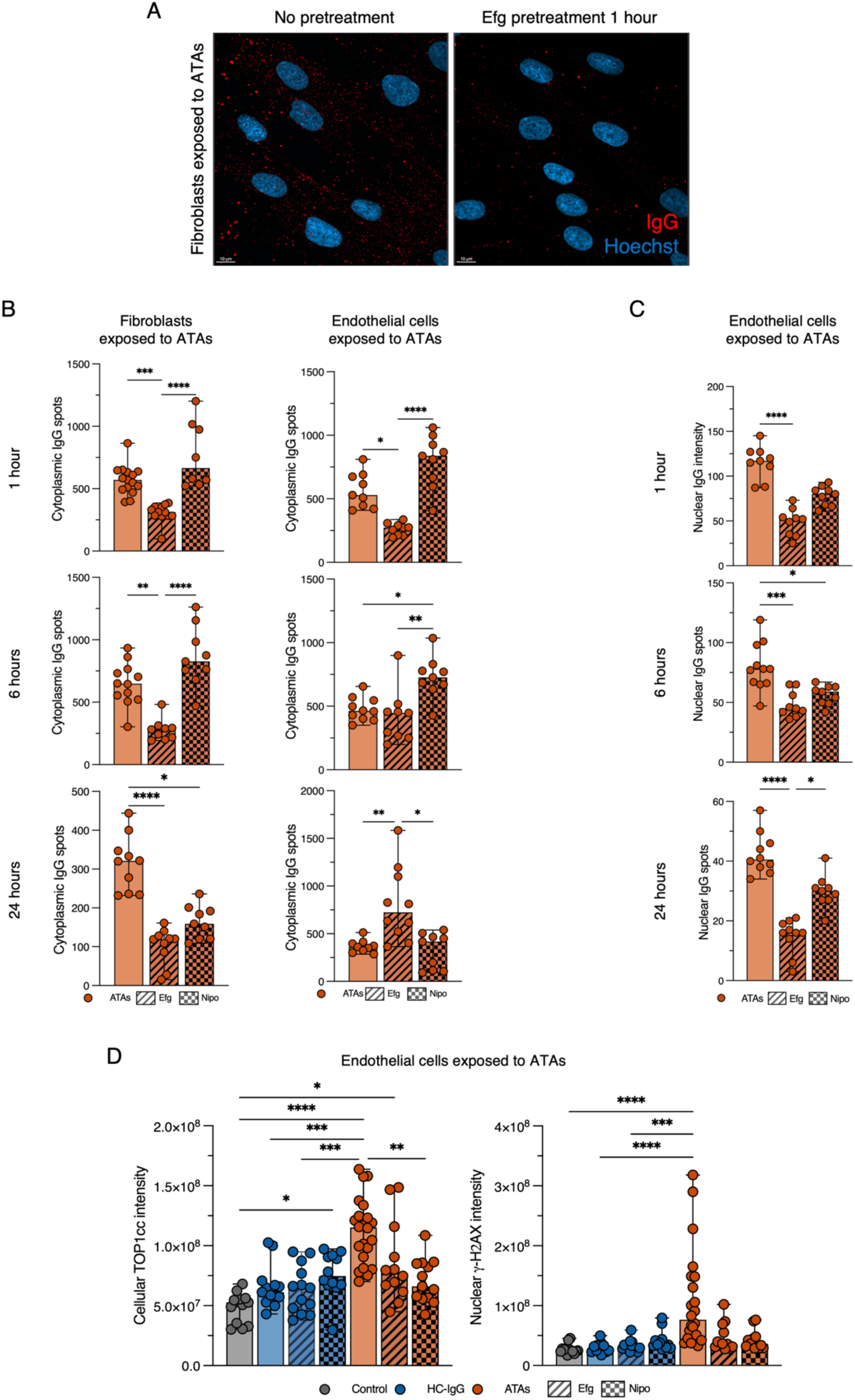
Anti-FcRn agents in cellular models. **(A)** Representative confocal micrographs of fibroblasts exposed for 1 hour to ATAs (25 µg/mL) without (left) or with efgartigimod (Efg, right) pretreatment for 1 hour (25 µg/mL). **(B)** Quantification of intracytoplasmic IgG deposits in fibroblasts (left) and endothelial cells (right) pretreated with anti-FcRn agents (Efg, 25 µg/mL or nipocalimab (Nipo), 10 µg/mL) following ATAs (25 µg/mL) exposure for 1, 6, and 24 hours. Efg pretreatment reduced cytoplasmic IgG uptake at all times expected at 24 hours in endothelial cells. Nipo increased cytoplasmic IgG signal at all time points, except at 24 hours, in endothelial cells. This likely reflects non-specific detection of Nipo itself by the secondary anti-human IgG antibody, given that Nipo is a monoclonal human IgG1 antibody. *n* = 3 independent experiments, *n* ≥ 100 cells analyzed per condition. Box plots indicate median and full data range. ns: non-significant; *P < 0.05; **P < 0.01; ***P < 0.001; ****P < 0.0001; Kruskal-Wallis test with Dunn’s post hoc correction for multiple comparisons. **(C)** Quantification of intranuclear IgG deposits in endothelial cells pretreated with anti-FcRn agents (Efg, 25 µg/mL or Nipo, 10 µg/mL), following ATAs exposure (25 µg/mL) for 1, 6, and 24 hours. Anti-FcRn pretreatment reduced nuclear ATAs uptake at all time points. *n* = 3 independent experiments, *n* ≥ 100 cells analyzed per condition. Box plots indicate median and full data range. ns: non-significant; *P < 0.05; **P < 0.01; ***P < 0.001; ****P < 0.0001; Kruskal-Wallis test with Dunn’s post hoc correction for multiple comparisons. **(D)** Quantification of cellular TOP1cc (left) and intranuclear γ-H2AX (right) staining in endothelial cells pretreated with anti-FcRn agents (Efg, 25 µg/mL or Nipo, 10 µg/mL), following 24 hours of ATAs (25 µg/mL) exposure. *n* = 3 independent experiments, *n* ≥ 100 cells analyzed per condition. Box plots indicate median and full data range. ns: non-significant; *P < 0.05; **P < 0.01; ***P < 0.001; ****P < 0.0001; Kruskal-Wallis test with Dunn’s post hoc correction for multiple comparisons.

**Figure S17.**
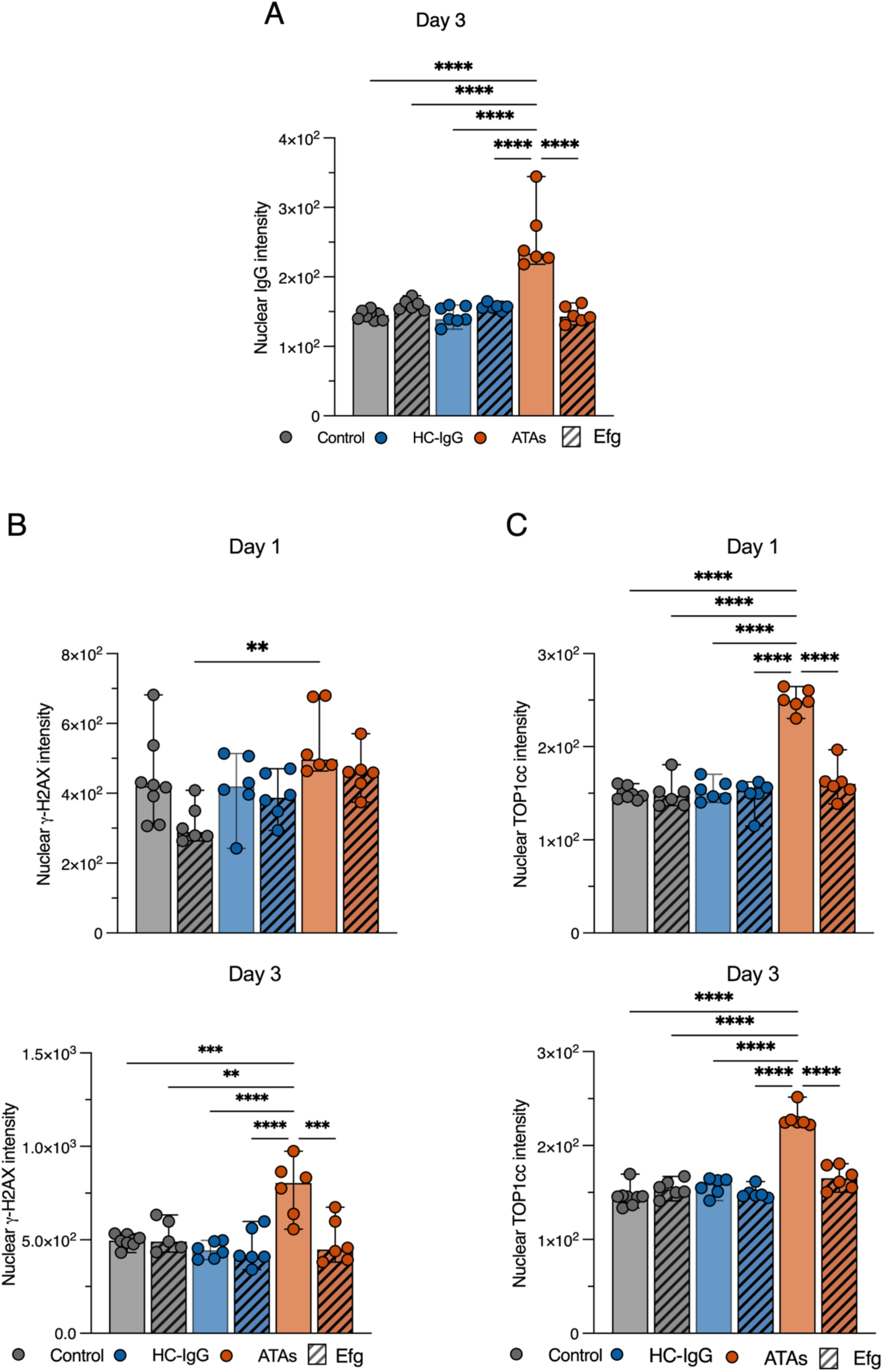
Anti-FcRn ameliorates ATAs impact in skin explants. (A) Quantification of intranuclear IgG staining in NativeSkin® explants exposed to medium culture (control, *n* = 4) or HC-IgG (450 µg, *n* = 3) or ATAs (300 µg, *n* = 3) ± Efgartigimod (Efg, 10 µg) for 3 days. Box plots indicate median and full data range. ****P < 0.0001; one-way ANOVA with Tukey’s post hoc correction for multiple comparisons. (B) Quantification of intranuclear γ-H2AX staining in NativeSkin® explants exposed to medium culture (control, *n* = 4) or HC-IgG (450 µg, *n* = 3) or ATAs (300 µg, *n* = 3) ± Efg (10 µg), for 1 (top) or 3 (bottom) days. Box plots indicate median and full data range. ****P < 0.0001; one-way ANOVA with Tukey’s post hoc correction for multiple comparisons. (C) Representative confocal micrographs (top) and quantification (bottom) of intracellular TOP1cc staining in NativeSkin® explants exposed to medium culture (control, *n* = 4) or HC-IgG (450 µg, *n* = 3) or ATAs (300 µg, *n* = 3) ± Efg (10 µg), for 1 (top) or 3 (bottom) days. *n* = 3 independent experiments. Box plots indicate median and full data range. ****P < 0.0001; one-way ANOVA with Tukey’s post hoc correction for multiple comparisons.

**Figure S18.**
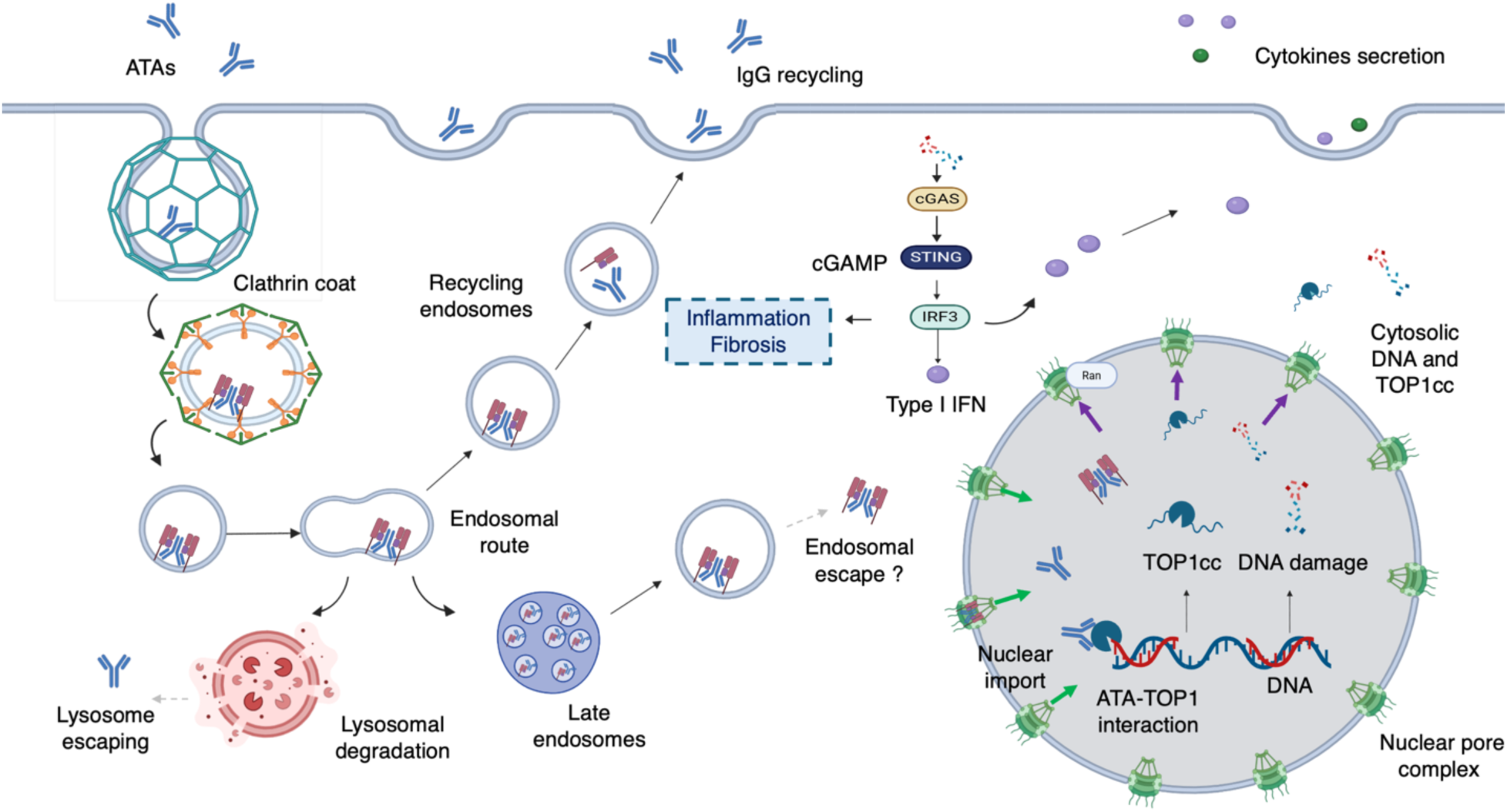
Pathogenicity of ANAs in CTDs: ATAs in SSc as a paradigm. ANAs enter living cells, accumulate in the nucleus, and engage their intracellular targets. In the case of ATAs, nuclear accumulation leads to direct inhibition of topoisomerase I catalytic activity, induction of DNA damage, and activation of type I interferon production via the cGAS–STING pathway, ultimately promoting collagen deposition and fibrosis. The neonatal Fc receptor (FcRn) is identified as the key determinant of ANAs intracellular trafficking and nuclear access.

**Figure S19.**
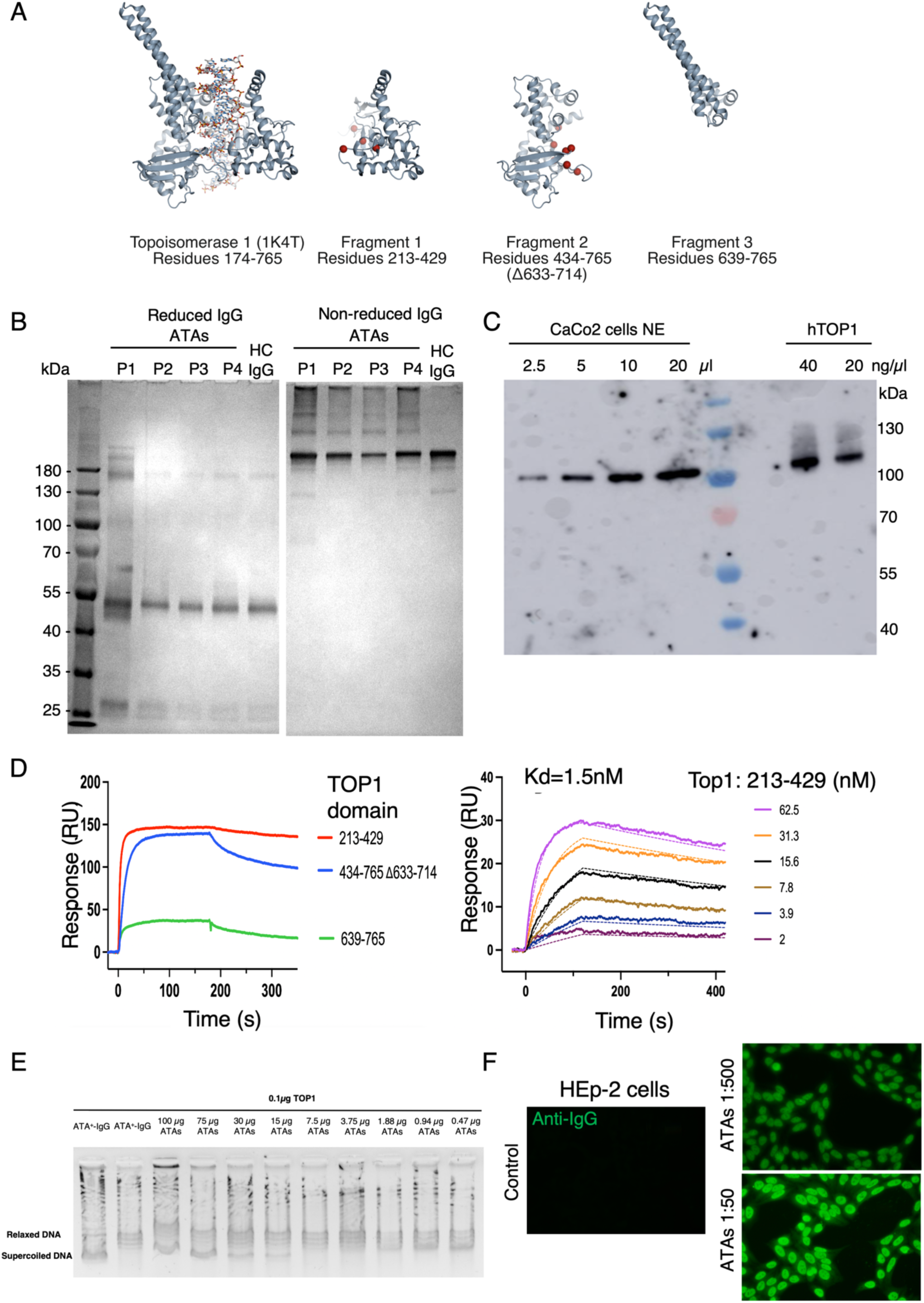
ATA purification. **(A)** Constructs for recombinant expression of TOP1. **(B)** Purification control of ATAs. ATAs (10 ng/µL) from four SSc donors and one HC IgG (10 ng/µL) loaded on SDS-PAGE in reduced (using beta-mercaptoethanol) or non-reduced form and visualized with silver staining. Size-based separations of the IgG were used to determine IgG integrity, showing bands of approximately 150 kDa for the non-reduced and 28 and 51 kDa for the reduced SDS-PAGE at each time point tested. The left lane is a molecular size marker. **(C)** Nuclear extract (NE) from CaCo2 cells and purified human topoisomerase 1 (hTOP1) were applied in varying volumes. An increasing amount of lysed CaCo2 cells produced a single band at 100 kDa, corresponding to human TOP1, when the gel was incubated with ATAs (10 ng/ml). **(D)** Representative SPR sensorgrams showing binding between ATA and the three domains of TOPI. Affinity measurements of ATA binding to the TopI_213-429 domain. **(E)** DNA relaxation assays in the presence of TOP1 with ATA^+^-IgG and commercial ATAs showing similar TOP1 inhibition capacity. **(F)** Indirect immunofluorescence on HEp-2 cells showing the DNA topoisomerase I (topo I)-like staining pattern, as defined by ICAP nomenclature (AC-29). Representative images are shown for the negative control (top), ATAs at 1:500 dilution (middle), and ATAs at 1:50 dilution (bottom).

## Movies S1-6

**Movie S1:** Classical live-cell microscopy of fibroblasts exposed to fluo-ATAs (acquisition started at *t* = 5 minutes after incubation with antibodies, 1-hour recording). Red: CellMask; green: fluo-ATAs; blue: Hoechst.

**Movie S2:** 3D high-resolution live-cell microscopy of fibroblast exposed to fluo-ATAs for 30 minutes. Nuclei were reconstructed based on the Hoechst signal. Blue: nucleus; green: cytoplasmic fluo-ATAs; orange: nuclear labelling of fluo-ATAs based on computerized nuclear segmentation.

**Movies S3-S5:** Cellular tracking examples of fluo-ATAs in fibroblasts showing nuclear translocation. Nuclei were reconstructed based on the Hoechst signal. For trajectory visualization, ATAs particles were detected using the Spots module (estimated diameters ranging from 0.15 to 0.8 µm) and tracked using the Imaris autoregressive motion algorithm (maximum frame-to-frame linking distance 5 µm). Particles were subsequently classified according to their position relative to the reconstructed nuclear surface and visualized as distinct color-coded populations inside or outside the nucleus Blue: nucleus; green: cytoplasmic fluo-ATAs; orange: nuclear labelling of fluo-ATAs based on computerized nuclear segmentation. S3/S4 are the same video without/with apparent tracking.

**Movie S6:** Video explaining the 3D cell reconstruction workflow using Z-stack acquisition of fibroblasts exposed to antibodies. Red: IgG; blue: Hoechst.

## Supplementary Tables S1 to S2

**Table S1.**
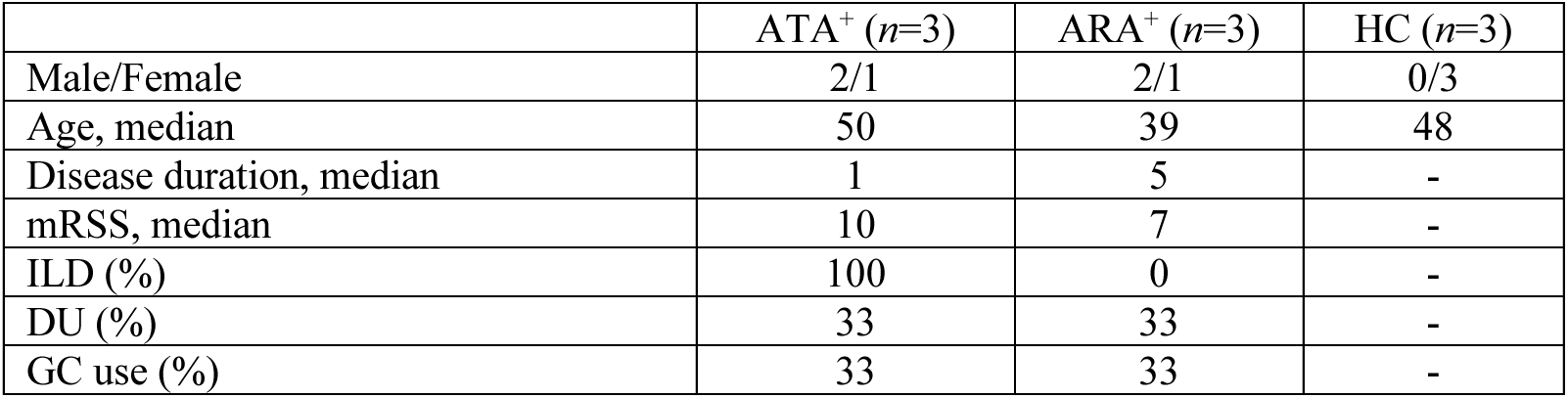
Individuals’ characteristics at the time of skin biopsy. ATA: anti-topoisomerase 1 antibodies; ARA: anti-RNA polymerase III antibodies; HC: healthy controls; mRSS: modified Rodnan Skin Score; ILD: interstitial lung disease; DU: digital ulcer; GC: glucocorticosteroids.

|  | ATA<sup>+</sup> (n=3) | ARA<sup>+</sup> (n=3) | HC (n=3) |
| --- | --- | --- | --- |
| Male/Female | 2/1 | 2/1 | 0/3 |
| Age, median | 50 | 39 | 48 |
| Disease duration, median | 1 | 5 | - |
| mRSS, median | 10 | 7 | - |
| ILD (%) | 100 | 0 | - |
| DU (%) | 33 | 33 | - |
| GC use (%) | 33 | 33 | - |

**Table S2.** Characteristics of the individuals included in the proteomics study. ATA: anti-topoisomerase 1 antibodies; ARA: anti-RNA polymerase III antibodies; HC: healthy controls; mRSS: modified Rodnan Skin Score; ILD: interstitial lung disease; DU: digital ulcer; GC: glucocorticosteroids; IS: immunosuppressant

|  | ATA<sup>+</sup> (n=10) | ACA<sup>+</sup> (n=10) | ATA<sup>-</sup>/ACA<sup>-</sup> (n=10) | HC (n=10) |
| --- | --- | --- | --- | --- |
| Female (%) | 70 | 90 | 80 | 80 |
| Age, median (IQR) | 51 (38-56) | 55 (50-61) | 51 (45-57) | 52 (45-56) |
| Disease duration, median (IQR) | 2 (1-6) | 5 (2-8) | 8 (3-13) | - |
| mRSS, median (IQR) | 13 (9-22) | 2 (2-4) | 8 (3-12) | - |
| ILD (%) | 90 | 30 | 70 | - |
| DU (%) | 70 | 40 | 30 | - |
| PAH (%) | 10 | 40 | 0 | - |
| GC use (%) | 70 | 0 | 90 | - |
| IS use (%) | 60 | 0 | 60 | - |

